# Bean extract-based gargle for efficient diagnosing COVID-19 at early-stage using rapid antigen tests : a clinical, prospective, diagnostic study

**DOI:** 10.1101/2021.08.13.21261463

**Authors:** Joseph Kwon, Euna Ko, Se-Young Cho, Young-Ho Lee, Sangmi Jun, Kyuhong Lee, Eunha Hwang, Bipin Vaidya, Jeong-Hwan Hwang, Joo-Hee Hwang, Namsu Kim, Mi-Kyung Song, Hye-Yeon Kim, Dai Ito, Yuxi Lin, Eunae Jo, Kyeong Eun Yang, Hee-Chung Chung, Soyoung Cha, Dong Im Kim, Yoon-Sun Yi, Sung-Ho Yun, Sun Cheol Park, Sangmin Lee, Jong-Soon Choi, Dal Sik Kim, Duwoon Kim

## Abstract

**Importance:** The antigen-based rapid diagnostic test (Ag-RDT), using saliva specimens, is fast, non-invasive and suitable for SARS-CoV-2 self-testing, unlike nasopharyngeal swab (NPS) testing.

**Objective:** To assess the diagnostic sensitivity of a novel Beanguard gargle™ (BG)-based virus detection method for early diagnosis of COVID-19.

**Design:** This clinical trial was conducted at Gunsan Medical Center, Namwon Medical Center, and Jeonbuk National University Hospital, between May 7 and July 7, 2021.

**Setting:** Paired NPS and BG-based saliva specimens collected from COVID-19 patients and healthy individuals were analyzed using NPS-RT-PCR, BG-RT-PCR, and BG-Ag-RDTs.

**Participants:** The study comprised 102 COVID-19-positive patients hospitalized after governmental screening process and 100 healthy individuals. Forty-five COVID-19 patients were sampled within 6 days of illness and 57 within 7–15 days; 27 were categorized as asymptomatic and 75, as symptomatic. Eight and 2 patients carried the SARS-CoV-2 Alpha and Delta variants, respectively.

**Intervention:** The diagnostic performances of BG-Ag-RDT, BG-RT-PCR, and NPS-RT-PCR for detecting SARS-CoV-2 were compared.

**Main outcomes:** The sensitivities of BG-Ag-RDT and BG-RT-PCR towards salivary viral detection were highly concordant, with no discrimination between symptomatic, asymptomatic, or SARS-CoV-2 variant cases.

**Results:** Among total participants (mean age, 43.7 years), 51% were women. BG-Ag-RDTs showed high sensitivity (97.8%, [95% CI, 88.4% to 99.6%]) and specificity (100%, [95% CI, 96.3% to 100%) in 45 patients within 6 days of illness and could detect all cases of SARS-CoV-2 Alpha and Delta variants. In 11 asymptomatic early-stage cases, both BG-Ag-RDTs and BG-RT-PCR showed excellent sensitivity and specificity of 100% (95% CI, 74.1% to 100% and 95% CI, 20.7% to 100%, respectively). The interaction between SARS-CoV-2 spike proteins and truncated canavalin, an active ingredient from bean extract (BE) and the ultrastructural features of SARS-CoV-2 particles coated with BE were observed. The detachment of the SARS-CoV-2 receptor-binding domain from hACE2 increased as the BE concentration increased, allowing the release of the virus from hACE2 for early diagnosis.

**Conclusions and Relevance:** Using BG-based saliva remarkably enhances the Ag-RDT diagnostic performance as an alternative to NPS and enables rapid and accurate COVID-19 self-testing and mass screening, supporting efficient COVID-19 management.

**Trial Registration:** KCT0006438

**Key Points:** *Question:* How can we collect SARS-CoV-2 from oral cavity to improve the sensitivity of antigen-based rapid diagnostic test (Ag-RDT)?

*Findings:* In this clinical study involving 102 hospitalized COVID-19 patients, the Ag-RDT test using Beanguard gargle™-based saliva specimens showed significantly enhanced sensitivity and specificity towards detection of SARS-CoV-2 along with Alpha and Delta variants in all patients tested within 6 days of illness.

*Meaning:* Our self-testing method represents an attractive alternative to nasopharyngeal swab RT-PCR for the early diagnosis of symptomatic and asymptomatic COVID-19 cases.

## INTRODUCTION

The prevalence of coronavirus disease-2019 (COVID-19) caused by severe acute respiratory syndrome coronavirus-2 (SARS-CoV-2) has significantly altered daily living and rapidly become a global health threat,^1^ with variants rapidly spreading across the world.^2^ Therefore, accurate identification and rapid diagnosis of SARS-CoV-2 is critical for lowering continued transmission and gaining control over the current pandemic.

RT-PCR-based quantification of SARS-CoV-2 RNA obtained from the upper respiratory tract via nasopharyngeal swabs (NPSs) has been universally adopted as the reference standard for viral detection.^3^ Although this method offers high sensitivity and specificity for SARS-CoV-2 detection, it requires skilled professionals and sophisticated instrumentation, as well as long period of time for detection.^4^ Therefore, a rapid and improved self-diagnostic strategy is required to facilitate early detection and prevent COVID-19 spread.

Antigen detection using lateral flow-based rapid diagnostic tests (known as Ag-RDTs) are widely used to provide on-site diagnosis and mass screening for the early detection of pathogens,^5^ prior to becoming a significant risk for community transmission.^6,7^ However, the performance of several commercial RDTs is highly variable and additional method is required to enhance their sensitivity for precise clinical diagnosis and subsequent medical action.^8^

The sensitivity of RDT can be improved by selecting appropriate specimens with alternative sampling techniques, which is critical to ensure accurate disease diagnosis without false test results. As an alternative to NPS, saliva-based specimens have attracted increasing attention for the diagnosis of respiratory infection.^9–11^ The non-invasive and self-collection methods associated with saliva specimens have the potential to increase population-based surveillance coverage without increasing the risk of exposure to nosocomial virus infections during the testing process. Several studies have reported that saliva, laden with virus, can serve as a transient medium for the transmission of SARS-CoV-2, which is broadly enriched on the epithelial cells lining the oral cavity and oral mucosae.^12–14^ Moreover, the U.S. Food and Drug Administration, and the Centres for Disease Control and Prevention have updated their guidelines to include saliva-based COVID-19 testing.^15,16^ Virus detection in saliva have been adopted by different techniques such as chemiluminescence immunoassay,^17^ electrochemical analysis,^18^ and fluorescence assay.^19^ Although saliva specimens have advantages, including self-collection of sample, it is difficult to obtain sufficient titers of virus attached to oral cavity and to apply directly saliva specimen itself to conventional RDTs without further treatment.

The current study aims to improve the efficiency of COVID-19 diagnostics using a novel saliva collection method with bean extract (BE)-based Beanguard gargle™ (BG, BIO3S, Inc., Republic of Korea). BG-RT-PCR and BG-Ag-RDT for SARS-CoV-2 exhibited outstanding analytical performance with the highest sensitivity of 100% and 97.8% compared to NPS-RT-PCR, respectively. Moreover, the BG-Ag-RDT met the minimum performance criteria for early diagnosis of COVID-19, as recommended by the World Health Organization (WHO) (≥ 80% sensitivity and ≥ 97% specificity, compared to a standard PCR method).^5^ These results suggest that the proposed BG-based saliva specimen collection method can serve as an efficient and promising strategy for self-diagnosis and on-site COVID-19 screening, thus, facilitating the identification, and subsequent isolation, of symptomatic and asymptomatic cases at early stage, as well as the detection of SARS-CoV-2 variants.

## METHODS

### Study design and participants

In this study, paired NPS and BG-based saliva specimens were collected from 102 patients with COVID-19 who were hospitalized at Gunsan Medical Center, Namwon Medical Center, and Jeonbuk National University Hospital, between May 7 and July 7 of 2021, and 100 healthy subjects as negative controls; statistically, we estimated a minimum sample size to obtain 95% sensitivity. A schematic study design and procedures are shown in the Supplement 2. The specimens were then analyzed with NPS-RT-PCR, BG-RT-PCR, and BG-Ag-RDTs. The baseline clinical and demographic data of enrolled patients, presented in Table 1 and Figure 1, did not show any significant differences. The date of patient’s clinical symptoms onset, hospitalization, and initial confirmation of COVID-19 were recorded by healthcare professionals (Supplement 1). Ethical approval for the study was granted by the Institutional Review Board of Jeonbuk National University Hospital (CUH 2021-04-036-002) and written informed consent statements were obtained from all participants. The inclusion and exclusion criteria are listed in Supplement 2.

**Table 1.** Characteristics of the participants.

|  | **SARS-CoV-2 positive (n=102)** | **Healthy subjects (n=100)** |
| --- | --- | --- |
| **Demographic composition of participants** |  |  |
| **Sex [no. (%)]** |  |  |
| **Female** | 37 (36.3) | 68 (68.0) |
| **Male** | 65 (63.7) | 32 (32.0) |
| **Age (years)** |  |  |
| **Average** | 45.7 | 40.4 |
| **Range** | 18–83 | 23–60 |
| **Patient with days of illness [no. (%)]** |  |  |
| **≤ 6 days** | 45 (44.1) | .. |
| **> 6 days** | 57 (55.9) | .. |
| **Symptomatic [no. (%)]** | 75 (73.5) | .. |
| **Asymptomatic [no. (%)]** | 27 (26.5) | .. |
| **SARS-CoV-2 variant [no. (%)]** |  |  |
| **Alpha** | 8 (7.8) | .. |
| **Delta** | 2 (2.0) | .. |
| **Days of illness (days)** |  |  |
| **Average** | 7.9 | .. |
| **Range** | 2–15 | .. |

**Figure 1.**
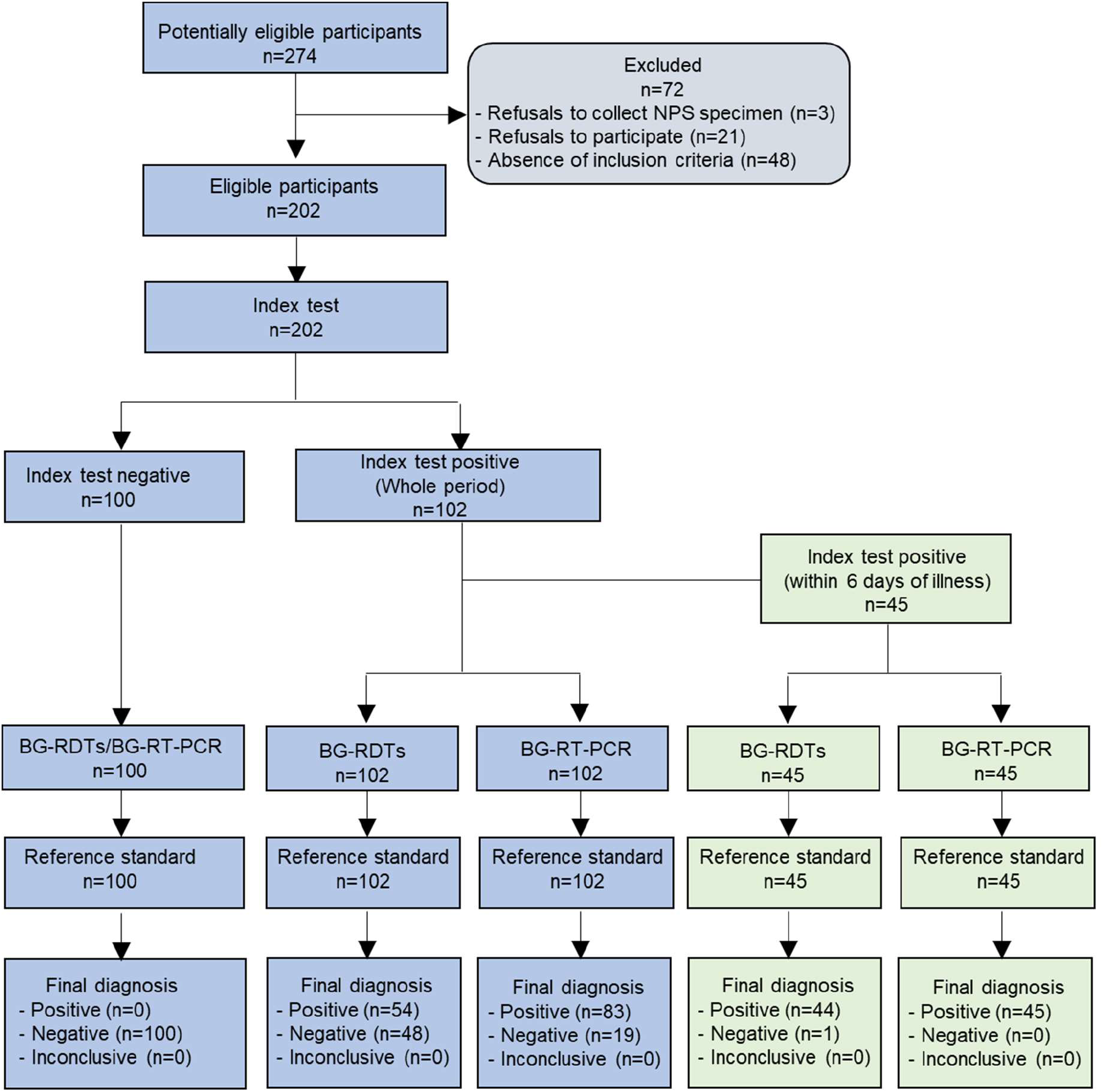
Enrolment and outcomes of participants in this clinical trial. Among total participants (n=202), 86 participants (42.6%) were 18 to 39 years of age, 106 participants (52.5%) were aged 40 to 69 years, and 10 participants (5.0%) were older than 70 years. Nasopharyngeal swabs-based RT-PCR (NPS-RT-PCR) was adopted as reference standard.

### Procedures

Medical professionals collected NPS specimens as a reference from participants and saliva specimens by asking participants to spit into a tube after swirling and gargling 5 mL of BG for 2 min. The RT-PCR diagnosis for SARS-CoV-2 in NPS and saliva specimens was performed using Allplex 2019-nCoV Real-time PCR (Seegene, Seoul, Korea), with RNA-dependent RNA polymerase (RdRP), envelope protein (E) and nucleocapsid protein (N) genes as the targets, and a STANDARD M nCoV Real-Time Detection kit (SD BIOSENSOR, Suwon, Korea), with RdRP and E genes as the targets, according to the manufacturer’s instructions. When all target genes are detected, the RT-PCR result for SARS-CoC-2 RNA is considered as positive. For BG-Ag-RDTs, we used two different COVID-19 Ag-RDT kits, namely, STANDARD Q COVID-19 Ag Saliva test (SD BIOSENSOR, Suwon, Korea) and Gmate® COVID-19 Ag Saliva (AG-020, Philosys Co., Ltd, Korea). BG-based saliva specimens (200 μL) were gently mixed with equal volumes of extraction buffer included in each COVID-19 antigen test kit. Subsequently, the treated specimen was added to the sample well of the test cassette, according to the manufacturer’s instructions. The test results obtained from Ag-RDTs were confirmed with the naked eye and the appearance of a colored band in both control and test line indicates positive results.

The interaction of BE with the receptor binding domain (RBD)-human version of angiotensin-converting enzyme 2 (hACE2) complex was evaluated by ELISA and the encapsulation of BE on HCoV-229E and SARS-CoV-2 surface was visualized by cryo-EM. Intermolecular interactions between truncated canavalin (TCan) purified from BE and spike proteins and molecular properties of TCan were examined by calorimetry and various biophysical approaches, respectively. Further details on calorimetry, molecular characterization, cryo-EM, ELISA, cell viability, reactive oxygen species (ROS) production, and statistical analysis are described in the Supplementary appendix.

### Statistical analysis

All statistical analyses were performed using GraphPad Prism v.7, and statistical multiple comparisons were performed using one-way ANOVA followed by post hoc Dunnett’s test. Data are expressed as mean ± SD. Significance was set at *p* < .05. We determined the significant difference between BG-RT-PCR and NPS-RT-PCR for SARS-CoV-2 using two-sided Wilcoxon signed-rank and two-sided Mann-Whitney rank-sum tests. We reported the diagnostic accuracy of sensitivity, specificity, positive percent agreement (PPA) and negative percent agreement (NPA) with 95% confidence intervals. Bland-Altman analysis was used to assess method agreement between BG-RT-PCR and NPS-RT-PCR. The average refers to the mean Ct values of BG-RT-PCR and NPS-RT-PCR, and the difference is obtained by subtracting the Ct values for BG-RT-PCR from those of NPS-RT-PCR.

Minimal sample size was calculated with 90 percent power at a significance level of 0.05 for the estimated sensitivity and specificity, according to the equation:

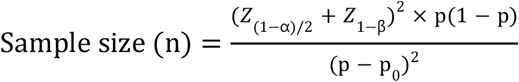

Where, *Z*_*(1-α)/2*_ and *Z*_*1-β*_ are Z-values for the corresponding level of confidence and the desired power, respectively. *p* is the estimated sensitivity or specificity, and *p*_*0*_ is the sensitivity or specificity of the reference. In this study, n = 50 for 95% sensitivity with 85% reference sensitivity, and n = 65 for 99% specificity with 95% reference specificity.

## RESULTS

### Trial population characteristics

The mean ages of the participants were 45.7 ± 16.00 years for patients with COVID-19 and 40.4 ± 11.18 years for healthy subjects. Of the 102 patients initially confirmed by COVID-19 government’s screening, 45 (44.1%) were sampled within 6 days of illness and 57 (55.9%) within 7 to 15 days of illness. Additionally, 27 (26.5%) patients were categorized as asymptomatic group, while 75 (73.5%) were assigned to the COVID-19 symptomatic group characterized by symptoms, including cough (48.0%), sore throat (41.3%), fever (40.0%), headache (18.7%), myalgia (18.7%), chest pain (6.7%), chills (6.7%), fatigue (6.7%), diarrhea and nausea (5.3%), dry mouth (5.3%), and loss of taste or smell (4.0%). In addition, 8 (7.8%) and 2 (2.0%) of the 102 patients carried the SARS-CoV-2 B.1.1.7 (Alpha) and B.1.617.2 (Delta) variants, respectively.

### Application of BG-RT-PCR for SARS-CoV-2 detection

Of the total 202 clinical specimens, 102 (50.5%) were positive via NPS-RT-PCR for SARS-CoV-2, with the cycle threshold (Ct) ranging from 11.1 to 39.3, and 100 (49.5%) were negative (Figure 1 and eTable 1 in Supplement 2). For 45 samples taken from individuals within 6 days of illness, both sensitivity and specificity of BG-RT-PCR against NPS-RT-PCR were 100% (95% confidence interval [CI], 92·1–100) and 100% (95% CI, 96·3–100), respectively (Table 2 and eTable 2 in Supplement 2). In 11 asymptomatic cases within 6 days of initial confirmation of COVID-19, both sensitivity and specificity of BG-RT-PCR against NPS-RT-PCR were also 100% (95% CI, 74.1– 100 and 95% CI, 20.7–100, respectively). All specimens tested positive for the SARS-CoV-2 Alpha (B.1.1.7) and Delta (B.1.617.2) variants were detectable using BG-RT-PCR within 6 days of illness (Table 3 and eTable 3 in Supplement 2). In all COVID-19 positive patients in our clinical trial, the overall sensitivity of BG-RT-PCR were 81.4% (95% CI, 72.7–87.7) (eTable 1 in Supplement 2).

**Table 2.** Evaluation of the diagnostic performance of BG-RT-PCR and BG-Ag-RDTs for negative controls (n=100) and COVID-19 patients within 6 days of illness (n=45)

|  |  | Sensitivity (%) | Specificity (%) | PPA (%) | NPA (%) |
| --- | --- | --- | --- | --- | --- |
|  |  | (95% CI) | (95% CI) | (95% CI) | (95% CI) |
| BG-RT-PCR compared with NPS-RT-PCR |  |  |  |  |  |
| BG-RT-PCR (n) | Participants (145) | 100 | 100 | 100 | 100 |
|  |  | (92.1–100) | (96.3–100) | (92.1–100) | (96.3–100) |
|  | Asymptomatic (11) | 100 | 100 | 100 | 100 |
|  |  | (74.1–100) | (20.7–100) | (74.1–100) | (20.7–100) |
| BG-Ag-RDT compared to BG-RT-PCR |  |  |  |  |  |
| BG-Ag-RDT<sup>a</sup> (n) | Participants (145) | 97.8 | 100 | 100 | 99.0 |
|  |  | (88.4–99.6) | (96.3–100) | (92.0–100) | (94.6–99.8) |
|  | Asymptomatic (11) | 100 | 100 | 100 | 100 |
|  |  | (74.1–100) | (20.7–100) | (74.1–100) | (20.7–100) |
| BG-Ag-RDT compared to NPS-RT-PCR |  |  |  |  |  |
| BG-Ag-RDT<sup>a</sup> (n) | Participants (145) | 97.8 | 100 | 100 | 99.0 |
|  |  | (88.4–99.6) | (96.3–100) | (92.0–100) | (94.6–99.8) |
|  | Asymptomatic (11) | 100 | 100 | 100 | 100 |
|  |  | (74.1–100) | (20.7–100) | (74.1–100) | (20.7–100) |

PPA, positive predictive agreement; NPA, negative predictive agreement.
<sup>a</sup>Two saliva-based Ag-RDTs were assessed: 1) STANDARD™ Q COVID-19 Ag Saliva test, and 2) Gmate® COVID-19 Ag Saliva. There is no difference between the test results of two Ag-RDTs tested for negative controls and COVID-19 patients within 6 days of illness.

**Table 3.**
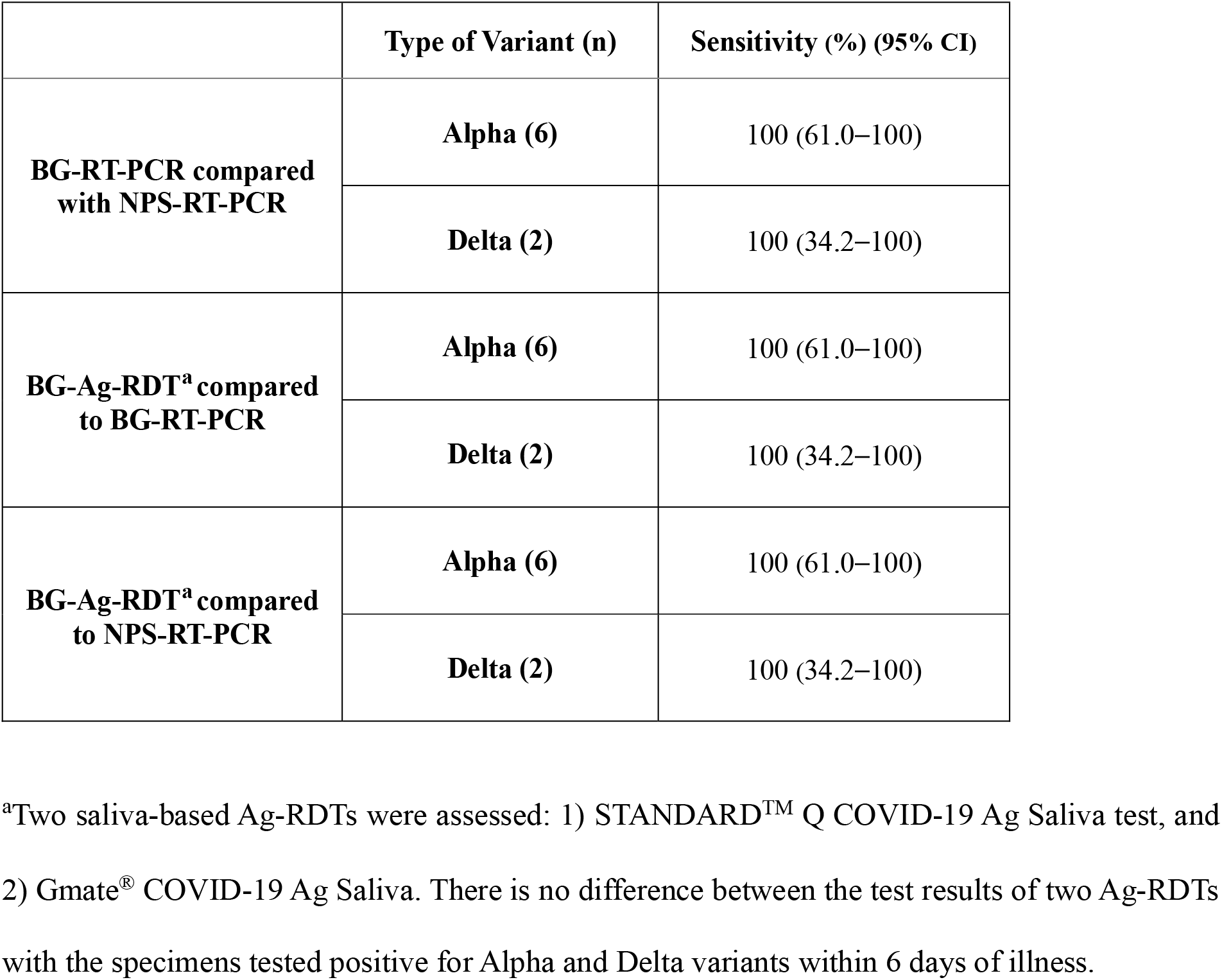
Evaluation of the sensitivity of BG-RT-PCR and BG-Ag-RDTs for patients of SARS-CoV-2 Alpha and Delta variants within 6 days of illness.

Comparing the Ct values between BG-RT-PCR and NPS-RT-PCR, samples from cases within 6 days of symptom onset or initial confirmation of COVID-19, the mean and median Ct values of BG-RT-PCR were 16.1 (95% CI, 14.9-17.3) and 15.8 [interquartile range (IQR), 13.0-18.3], respectively, whereas those of NPS-RT-PCR were 18.3 (95% CI, 17.0-19.6) and 17.8 [IQR, 15.2-20.4], respectively. In asymptomatic cases within 6 days of initial confirmation, the mean and median Ct values of BG-RT-PCR were 15.9 (95% CI, 13.4-18.4) and 15.8 [IQR, 13.2-18.5], respectively, whereas those of NPS-RT-PCR were 18.7 (95% CI, 15.8-21.6) and 17.9 [IQR, 14.7-22.4], respectively, indicating that BG-RT-PCR and NPS-RT-PCR are equivalent (Figure 2A and B, and eFigure 1 in Supplement 2). Notably, the scatter plots also showed that the Ct values of BG-RT-PCR were positively correlated with that of NPS-RT-PCR, and their mean differences of Ct values for all cases and those within 6 days of illness were 2.1 and 2.2, respectively (eFigure 1 in Supplement 2).

**Figure 2.**
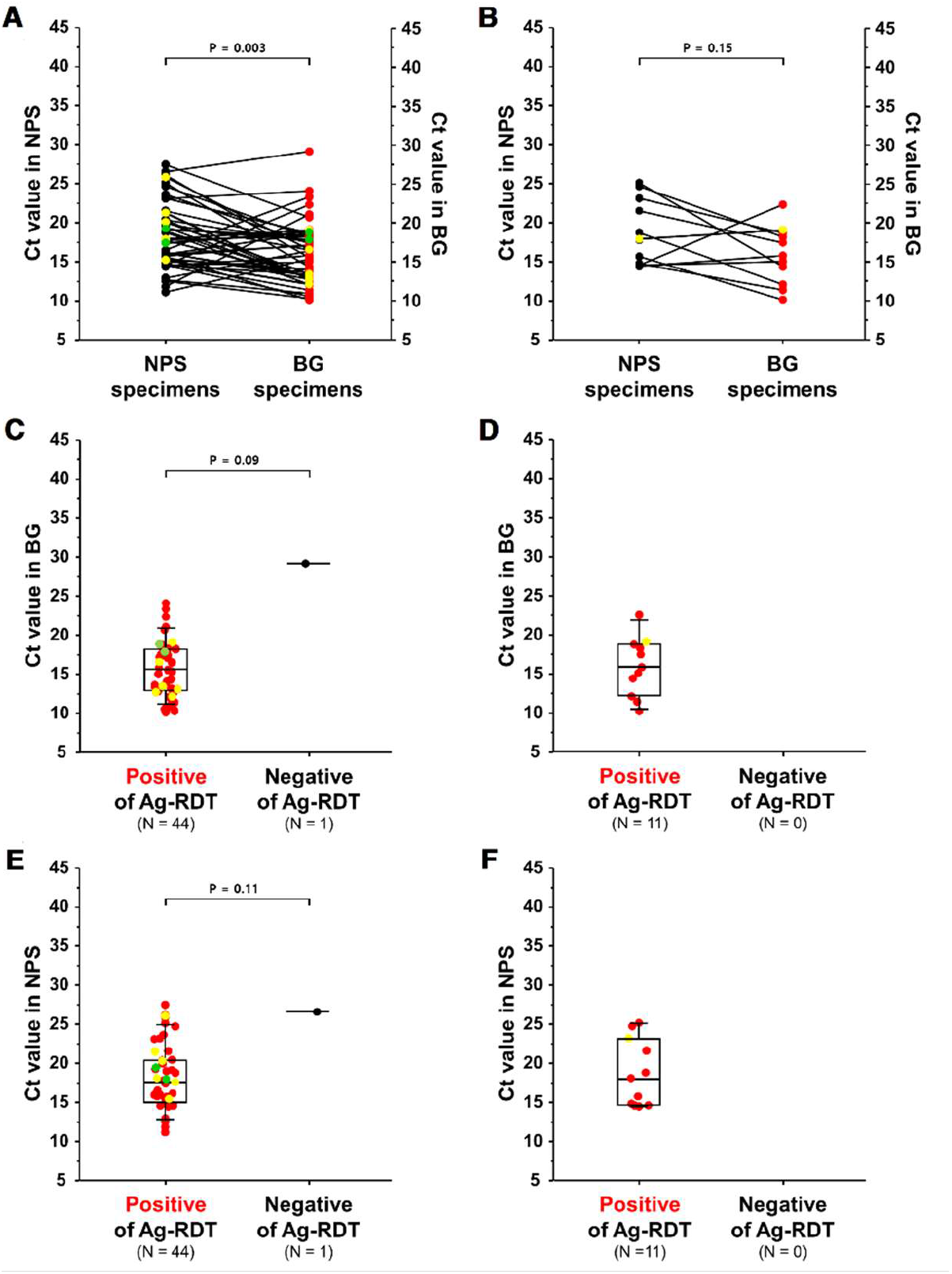
Comparison of Ct values from Beanguard gargle™-based RT-PCR (BG-RT-PCR) with NPS-based RT-PCR (NPS-RT-PCR) analysis. (A) all patients and (B) asymptomatic patients within 6 days of initial COVID-19 confirmation. Among the positive COVID-19 cases within 6 days of illness, the relationship between BG-based antigen rapid detection test results and Ct values of BG-RT-PCR with the RdRP gene for all patients (C) and asymptomatic cases (D). (E, F) The relationship between BG-Ag-RDTs and the Ct values of NPS-RT-PCR within 6 days of COVID-19 patients and asymptomatic cases, respectively. Yellow and green circles represent data point of Alpha and Delta variants, respectively.

### SARS-CoV-2 Ag-RDTs using BG-based saliva specimens

The sensitivity and specificity of two commercial Ag-RDTs using BG-based saliva within 6 days of illness were 97.8% (95% CI, 88.4–99.6) and 100% (95% CI, 96.3–100), respectively (Table 2 and eTable 2 in Supplement 2). Notably, the sensitivity and specificity of BG-Ag-RDT for asymptomatic cases within 6 days from initial confirmation were also 100% (95% CI, 74.1–100) and 100% (95% CI, 20.7–100), respectively. BG-Ag-RDTs could detect all cases of SARS-CoV-2 Alpha and Delta variants within 6 days of illness (Table 3 and eTable 3 in Supplement 2). Additionally, the sensitivity of BG-Ag-RDT for positive samples with Ct ≤ 30 within 6 days of illness was 97.8% (95% CI, 88.4–99.6; eTable 4 in Supplement 2).

The BG-Ag-RDT results were also displayed with the Ct values of BG-RT-PCR and NPS-RT-PCR (Figure 2C– F). In cases within 6 days of symptom onset or initial confirmation of COVID-19, the mean and median Ct values of BG-RT-PCR for positive Ag-RDT samples were 15.8 (95% CI, 14.7–16.9) and 15.7 [IQR, 12.9–18.3], respectively (Figure 2C). For the asymptomatic cases within 6 days from the initial confirmation, the mean and median Ct values of BG-RT-PCR were 15.9 (95% CI, 13.4–18.4) and 15.8[IQR, 13.2–18.5], respectively (Figure 2D). Additionally, the mean and median Ct values of NPS-RT-PCR for the positive Ag-RDT samples within 6 days of illness were 18.1 (95% CI, 16.8–19.4) and 17.6 [IQR, 15.0–20.4], respectively (Figure 2E). For the asymptomatic cases (initial confirmation within 6 days of illness), the mean and median Ct values of NPS-RT-PCR for positive BG-Ag-RDT samples were 18.7 (95% CI, 15.8–21.6) and 17.9 [IQR, 14.7–22.4], respectively (Figure 2F).

### Interaction of active ingredient with SARS-CoV-2

To examine the interaction of HCoV-229E and SARS-CoV-2 with BE contained in the gargle, the ultrastructure of whole HCoV-229E and SARS-CoV-2 before and after treatment with BE was observed by cryo-EM (Figure 3A–E). The viral particles were spherical with club-shaped spikes embedded in the envelope. After BE treatment, BE covered the surface of the coronavirus particle, providing the evidence that BE could effectively interact with SARS-CoV-2. Moreover, the results of transmission electron microscopy (TEM) using the negative staining method were consistent with those of cryo-EM (eFigure 2 in Supplement 2). To confirm that BE interferes with the interaction of hACE2 receptors and recombinant SARS-CoV-2 RBD-Fc-tagged proteins using indirect ELISA, the RBD–hACE2 receptor complexes attached to ELISA wells were washed two times with BE at concentrations ranging from 0 to 140 ppm (Figure 3F and G). ELISA results showed that BE effectively inhibited the binding of RBD and hACE2 in a dose-dependent manner up to 70 ppm.

**Figure 3.**
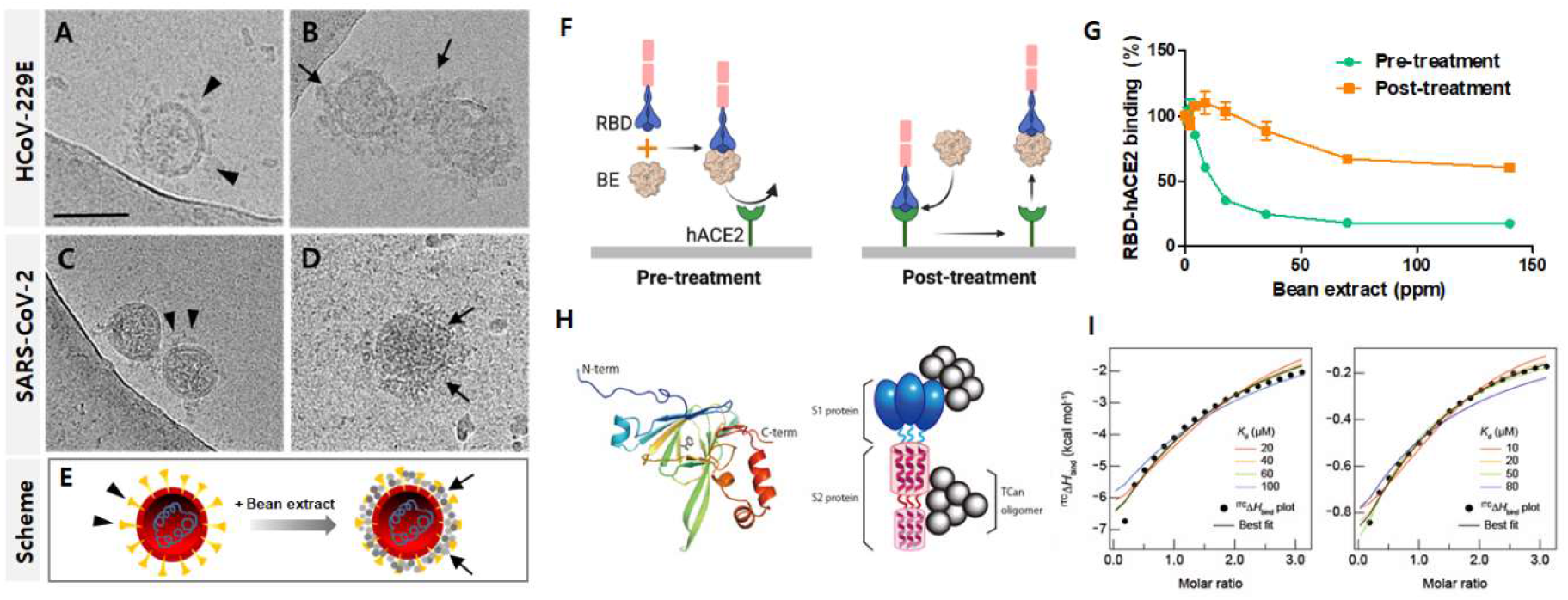
Intermolecular interaction of bean extract (BE) and truncated canavalin (TCan) with SARS-CoV-2. Cryo-EM images revealed the surface of HCoV-229E and SARS-CoV-2 surrounded by BE. Before (A) and after (B) exposure of BE to HCoV-229E and before (C) and after (D) exposure of BE to SARS-CoV-2 are presented with a schematic diagram, showing that BE is attached to the virus particle (E). The arrowheads and arrows indicate a spike and BE, respectively. Scale bar, 100 nm. The schematic diagram shows binding inhibition of the recombinant SARS-CoV-2 RBD-Fc-tagged protein to human version of angiotensin-converting enzyme 2 (hACE2) by pre- and post-treatment of BE (F). The illustrations were created using BioRender. The inhibition of RBD binding to hACE2 receptor was quantified by the difference in absorbance of labeled RBD between the test and control (G). The three-dimensional structure of TCan generated using a crystal structure of canavalin (PDB ID: 6v7j) (H, left panel). Binding of TCan oligomers (gray spheres) to S1 (blue ellipses) and S2 proteins (red helical structures) is schematically represented (H, right panel). Binding isotherms of TCan for S1 (I, left panel) and S2 (I, right panel) obtained from isothermal titration calorimetry are shown. Colored fit curves in G were obtained with a fixed dissociation constant (*K*_d_), and black curves show the best fit.

Next, we identified the active ingredient, truncated canavalin (TCan), from BE (eFigure 3 and 4 in Supplement 2), and showed that TCan has a stable three-dimensional structure and exists as an oligomer (Figure 3H and eFigure 5 in Supplement 2). Details regarding identification and molecular characterization of TCan are provided in Supplementary appendix. Furthermore, the binding capability of TCan to the two SARS-CoV-2 spike proteins, S1 and S2, was directly demonstrated using isothermal titration calorimetry (ITC) (figure 3I). TCan spontaneously bound to S1 and S2 (Figure 3I) by releasing heat, i.e., in an exothermic reaction, which showed an energetically favorable negative value of enthalpy change (^ITC^Δ*H*_bind_). The dissociation constant (*K*_d_) of TCan for S1 and S2 ranged between ∼20 – ∼100 μM and ∼10 – ∼80 μM, respectively. Additional details regarding ITC are provided in the Supplement 2 (eFigure 6-8).

### *In vivo* toxicity and *in vitro* cytotoxicity of BE

To evaluate the toxicity of BE, the toxic effects of BE from sword beans (*Canavalia gladiata*) and concanavalin A (Con A) on cell viability, lung and liver injuries, and inflammatory cells were compared. Con A, one of the components in sword bean, is well known for its mitogenic effect on splenocytes; it also activates the immune system, recruits lymphocytes, and elicits cytokine production.^20^ Results showed that BE neither induced toxic effects (based on analysis of survival rate, liver toxicity, and lung injury), nor exhibited cytotoxic effects or induce ROS generation (eFigure 9-16 in Supplement 2).

## DISCUSSION

Due to COVID-19 pandemic and the rapid spread of multiple SARS-CoV-2 variants, the demand for diagnostic tests of SARS-CoV-2 infection is increasing. However, the conventional NPS-based RT-PCR method, adopted as a reference standard, requires skilled health workers and personnel protective equipment for the workers during sampling. There is a risk of cross-infection due to the spread of contaminated aerosol during NPS specimen collection and handling. Moreover, NPS is not the first preference of patients due to the discomfort associated with insertion and removal of the swab. Hence, saliva as specimen that can be self-collected could be a suitable alternative to NPS. Even though previous studies reported application of saliva specimen for SARS-CoV-2 diagnosis, the sensitivity of saliva as specimen was considerably lower than that of NPS.^21,22^ Therefore, we focused on the use of a newly designed virus collection method, Beanguard gargle™, for user-friendliness and convenience of virus collection from oral cavity and increasing the efficacy of viral detection by collecting virus with BE.

In this study, BG-based saliva collection method remarkably increases sensitivity and specificity of Ag-RDT and RT-PCR for SARS-CoV-2 detection. Although the performance of saline gargle was evaluated for the detection of different respiratory viruses and SARS-CoV-2 using only RT-PCR, the application of saline gargle was not reported for Ag-RDTs.^23–25^ We show that BG-based saliva collection method can be compatible with Ag-RDTs and allow for rapid diagnosis of COVID-19 at early stage of infection without discriminating between symptomatic and asymptomatic cases.

Compared with the NPS-RT-PCR results, we found the performance of the BG-RT-PCR within 6 days of illness to be 100% sensitive and specific for SARS-CoV-2, including Alpha and Delta variant, suggesting the high sensitivity of BG-RT-PCR with wide adoptability to the virus variants. Furthermore, the sensitivity of the BG-Ag-RDT within 6 days of illness was enhanced up to 97.8% compared with the previously reported 65.0% sensitivity against salivary RT-PCR.^26^ All BG specimens of SARS-CoV-2, including Alpha and Delta variants, were also detected by BG-Ag-RDT, implying that the use of BG-Ag-RDT will facilitate rapid and accurate diagnosis of SARS-CoV-2 variants at early stage of COVID-19. Moreover, the diagnostic performance of the proposed BG-Ag-RDTs remarkably fulfilled WHO’s recommendation for the use of Ag-RDTs in early diagnosis of COVID-19 within 5–7 days of symptoms onset, which recommends ≥ 80% sensitivity and ≥ 97% specificity compared with that of the PCR assay.^5^

## LIMITATIONS

One limitation of this study is the exclusion of three participants from NPS sampling due to the inability to confirm the test. Furthermore, due to the small sample size of Alpha and Delta variants available for this study, the generalization of our test results for the variants could be limited.

## CONCLUSIONS

In conclusion, based on the interaction between BE and SARS-CoV-2, the BG-Ag-RDT offers an effective method with high sensitivity for self-diagnosis, on-site monitoring, and rapid testing of COVID-19 at early stages of infection. Moreover, this novel strategy will facilitate the execution of effective clinical responses by overcoming the long durations required for NPS-RT-PCR results and low sensitivity of Ag-RDTs.

## Data Availability

The complete de-identified participant data set will be available upon request to for researchers whose proposed use of the data has been approved, for any purpose. Data will be available with the publication. If needed, requests will require the ethics committee approval of the Jeonbuk National University Hospital in Jeonju (Republic of Korea). Anonymized data are fully available on reasonable request from the corresponding author after approval by the hospital ethics committee.

## ARTICLE INFORMATION

### Author Contributions

Drs. Kwon, D.S. Kim, and D. Kim had full access to all the data in the study and take responsibility for the integrity of the data and the accuracy of the data analysis.

*Concept and design*: D.S. Kim

*Acquisition, analysis, or interpretation of data*: Kwon, Jo. H. Hwang, N. Kim, Ko, Cho, Y. Lee, Jun, K. Lee, E. Hwang, Vaidya, Song, Ito, Lin, Jo, Yang, Chung, D. I. Kim, Yi, Park.

*Drafting of the manuscript*: Kwon, Ko, Cho, Y. Lee, Jun, K. Lee, E. Hwang, Vaidya, Je. H. Hwang, Jo. H. Hwang, N. Kim, D. S. Kim, D. Kim.

*Critical revision of the manuscript for important intellectual content*: All authors.

*Statistical analysis*: Je. H. Hwang, Jo. H. Hwang, N. Kim, Ko, Cho.

*Obtained funding*: Y. Lee, Choi, D. S. Kim, D. Kim.

*Administrative, technical, or material support*: E. Hwang, Je. H. Hwang, Jo. H. Hwang, N. Kim, H. Kim, Cha, Park, S. Lee

*Supervision*: D. S. Kim.

### Conflict of Interest Disclosures

Dr. D. S. Kim received grant support for this clinical study from BIO3S, Inc. Drs. D. Kim, Kwon, and Vaidya were involved in developing Beanguard gargle™. All disclosures are unrelated to the present clinical study. The remaining authors declare no competing interests.

### Funding/Support

This research work was supported by BIO3S, Inc. (2021-04-036-002, to Dr. D.S.K), Korea Basic Science Institute (Development Bio-active materials for Biological Disaster, C18310, to Dr. J.S.C) (Establishment of an integrated analysis system to overcome aging diseases caused by protein aggregation, C130000, to Dr. Y.H.L.) and Jeonbuk National University Hospital (CUH2016-0016, to Dr. D.S.K).

### Role of Funder/Sponsor

The funder (BIO3S, Inc. and Korea Basic Science Institute) was involved in the development of Beanguard gargle™, but had no role in the study design, data collection, analysis, and interpretation, or writing of the manuscript.

### Data Sharing Statement

The complete de-identified participant data set will be available upon request to for researchers whose proposed use of the data has been approved, for any purpose. Data will be available with publication. If needed, requests will require the ethics committee approval of the Jeonbuk National University Hospital in Jeonju (Republic of Korea). Anonymised data are fully available on reasonable request from the corresponding author after approval by the hospital ethics committee.

### Additional Contributions

We thank all the participants in this trial, the staff members at the trial locations, the members of the data and safety monitoring board, all the investigators at the clinical sites, Namwon, Gunsan Medical Center, and Jeonbuk National University Hospital.

## Notes

### Clinical Trial

KCT0006438

### Author Declarations

Ethical approval for the study was granted by the Institutional Review Board of Jeonbuk National University Hospital (CUH 2021-04-036-002) and written informed consent statements were obtained from all participants.

