## Supplement 1 for "Bean extract-based gargle for efficient diagnosing COVID-19 at early-stage using rapid antigen tests : a clinical, prospective, diagnostic study"

### Supplementary Appendix 1. Raw Data from 102 Patients with COVID-19

| Column | Description |
| --- | --- |
| 1 | Serial Number |
| 2 | Clinical Trial Patient Number |
| 3 | Age range (years) |
| 4 | Sex (Man=1, Woman=2) |
| 5 | Hospitaliztion Period (day) (from hospitalization date to clinical trial date) |
| 6 | Hospitaliztion Period of Symptomatic case (day) (from symtoms onset date to clinical trial date) |
| 7 | Hospitaliztion Period of Asymptomatic case (day) (from initial confirmed date to clinical trial date) |
| 8 | Presence or absence of symptoms at diagnosis (presence=1, absence=2) |
| 9 | Clinical Symtoms at diagnosis |
| 10 | Routes of transmission/infection (close contact=1, unknown=2) |
| 11 | Presence of SARS-CoV-2 Variant (Alpha=1, Delta=2) |
| 12 | NPS_PCR_Ct value, E gene |
| 13 | NPS_PCR_Ct value, RdRP gene |
| 14 | NPS_PCR_Ct value, N gene |
| 15 | NPS_PCR_Result (Positive=1, Negative=2) |
| 16 | Saliva_SARS-CoV-2 Antigen RDT [STANDARD Q COVID-19 Ag Saliva test (SD BIOSENSOR)] (Positive=1, Negative=2) |
| 17 | Saliva_SARS-CoV-2 Antigen RDT [Gmate® COVID-19 Ag Saliva (Philosys)] (Positive=1, Negative=2) |
| 18 | Saliva_PCR_Result (Positive=1, Negative=2, Inconclusive=3) |
| 19 | Saliva_PCR_Ct value, RdRP gene |
| 20 | Saliva_PCR_Ct value, E gene |

### Supplementary Appendix 1. Raw Data from 102 Patients with COVID-19

| 1 | 2 | 3 | 4 | 5 | 6 | 7 | 8 | 9 | 10 | 11 | 12 | 13 | 14 | 15 | 16 | 17 | 18 | 19 | 20 |
|---|---|---|---|---|---|---|---|---|---|---|---|---|---|---|---|---|---|---|---|
| 1 | J001 | 43~47 | 2 | 15 | 15 | 15 | 1 | fever, sore throat | 2 | | 30.92 | 31.79 | 33.78 | 1 | 2 | 2 | 3 | 34.35 | negative |
| 2 | J003 | 33~37 | 1 | 5 | 8 | 8 | 1 | cough | 1 | | 31.02 | 31.49 | 30.70 | 1 | 2 | 2 | 3 | 34.32 | negative |
| 3 | J005 | 36~40 | 1 | 5 | 7 | 7 | 2 | | 1 | | 28.73 | 29.17 | 28.69 | 1 | 2 | 2 | 1 | 26.77 | 26.06 |
| 4 | J006 | 77~81 | 1 | 11 | >11 | 11 | 1 | cough | 1 | | 35.35 | 35.01 | 33.94 | 1 | 2 | 2 | 1 | 33.23 | 31.72 |
| 5 | J008 | 68~72 | 1 | 11 | 13 | 13 | 1 | fatigue, muscle ache | 1 | | 21.84 | 21.54 | 21.75 | 1 | 2 | 2 | 1 | 22.04 | 22.35 |
| 6 | J009 | 45~49 | 1 | 7 | | 7 | 2 | | 1 | 1 | 27.54 | 28.88 | 26.74 | 1 | 1 | 1 | 1 | 19.82 | 19.53 |
| 7 | J011 | 36~40 | 1 | 6 | 6 | 6 | 1 | fever, sputum, sore throat | 2 | 1 | 16.12 | 17.78 | 15.38 | 1 | 1 | 1 | 1 | 13.05 | 13.29 |
| 8 | J012 | 44~48 | 1 | 6 | 6 | 6 | 1 | cough | 2 | 1 | 19.11 | 20.11 | 17.35 | 1 | 1 | 1 | 1 | 13.43 | 13.05 |
| 9 | J015 | 63~67 | 1 | 9 | 11 | 11 | 1 | fever, sore throat, cold sweat, chest tightness | 2 | | 30.24 | 31.00 | 32.40 | 1 | 2 | 2 | 3 | negative | 34.00 |
| 10 | J016 | 61~65 | 2 | 9 | 11 | 11 | 1 | fever | 1 | | 19.65 | 19.74 | 21.38 | 1 | 1 | 2 | 1 | 16.01 | 16.31 |
| 11 | J018 | 54~58 | 2 | 6 | 14 | 14 | 2 | | 1 | | 31.05 | 31.43 | 30.76 | 1 | 2 | 2 | 1 | 31.44 | 30.92 |
| 12 | J019 | 73~77 | 2 | 12 | | 13 | 2 | cough, headache | 2 | | 33.14 | 32.72 | 31.34 | 1 | 2 | 2 | 1 | 18.35 | 18.54 |
| 13 | J020 | 32~36 | 1 | 5 | 5 | 5 | 2 | | 1 | 1 | 17.69 | 17.91 | 15.65 | 1 | 1 | 1 | 1 | 19.08 | 18.97 |
| 14 | J021 | 48~52 | 1 | 3 | 6 | 6 | 1 | muscle ache, chills, headache | 1 | 1 | 20.57 | 21.30 | 18.53 | 1 | 1 | 1 | 1 | 12.69 | 12.72 |
| 15 | J022 | 61~65 | 2 | 4 | 4 | 4 | 1 | fever, cough | 1 | | 20.64 | 20.43 | 19.62 | 1 | 1 | 1 | 1 | 17.59 | 17.84 |
| 16 | J023 | 33~37 | 1 | 3 | 3 | 3 | 1 | fever, sore throat, muscle ache | 2 | | 10.50 | 12.52 | 9.97 | 1 | 1 | 1 | 1 | 10.32 | 10.44 |
| 17 | J024 | 53~57 | 1 | 3 | 5 | 5 | 1 | fever, sore throat, muscle ache, cough | 2 | | 11.93 | 14.58 | 10.90 | 1 | 1 | 1 | 1 | 10.56 | 10.74 |
| 18 | J025 | 51~55 | 1 | 3 | | 4 | 2 | | 1 | | 24.43 | 21.57 | 21.21 | 1 | 1 | 1 | 1 | 17.46 | 17.64 |
| 19 | J026 | 18~22 | 2 | 2 | | 3 | 2 | | 1 | | 16.76 | 18.78 | 16.33 | 1 | 1 | 1 | 1 | 12.07 | 12.07 |
| 20 | J027 | 53~57 | 1 | 4 | 4 | 4 | 1 | fever, chest tightness, headache, foreign body sensation of throat | 1 | 1 | 11.80 | 15.22 | 11.04 | 1 | 1 | 1 | 1 | 12.12 | 12.29 |
| 21 | J028 | 58~62 | 1 | 8 | >8 | 8 | 1 | dry mouth, fatigue | 1 | | 32.56 | 31.92 | 33.08 | 1 | 2 | 2 | 1 | 21.31 | 20.89 |
| 22 | J029 | 57~61 | 1 | 10 | 12 | 12 | 1 | sore throat, fever, sputum, throat tightness | 2 | | 31.13 | 31.07 | 33.96 | 1 | 2 | 2 | 1 | 28.04 | 27.46 |
| 23 | J031 | 81~85 | 1 | 9 | | 9 | 2 | | 1 | | 37.87 | 36.69 | 37.67 | 1 | 2 | 2 | 1 | 29.19 | 28.24 |

### Supplementary Appendix 1. Raw Data from 102 Patients with COVID-19

| 1 | 2 | 3 | 4 | 5 | 6 | 7 | 8 | 9 | 10 | 11 | 12 | 13 | 14 | 15 | 16 | 17 | 18 | 19 | 20 |
|---|---|---|---|---|---|---|---|---|---|---|---|---|---|---|---|---|---|---|---|
| **24** | J032 | 32~36 | 1 | 6 | 6 | 6 | 1 | fever, flu sensation | 1 | 1 | 24.41 | 25.89 | 22.21 | 1 | 1 | 1 | 1 | 16.57 | 16.47 |
| **25** | J033 | 32~36 | 1 | 11 | 12 | 12 | 1 | fever, cough | 1 | 1 | 21.50 | 22.89 | 21.07 | 1 | 2 | 2 | 1 | 22.31 | 21.84 |
| **26** | J036 | 69~73 | 1 | 10 | 11 | 11 | 1 | sore throat | 2 | | 34.38 | 34.95 | 33.19 | 1 | 2 | 2 | 3 | 35.96 | negative |
| **27** | J037 | 62~66 | 2 | 10 | 11 | 11 | 1 | dry mouth, fatigue | 1 | | 30.56 | 32.47 | 29.71 | 1 | 2 | 2 | 2 | negative | negative |
| **28** | J038 | 60~64 | 1 | 11 | | 11 | 2 | | 1 | | 38.65 | 39.3 | 38.73 | 1 | 2 | 2 | 2 | negative | negative |
| **29** | J040 | 58~62 | 1 | 3 | | 4 | 2 | | 1 | | 12.94 | 14.55 | 18.58 | 1 | 1 | 1 | 1 | 15.03 | 14.98 |
| **30** | J041 | 19~23 | 1 | 3 | | 4 | 2 | | 1 | | 16.26 | 15.7 | 19.59 | 1 | 1 | 1 | 1 | 10.13 | 10.15 |
| **31** | J042 | 49~53 | 1 | 2 | | 3 | 2 | | 2 | | 23.61 | 23.18 | 24.12 | 1 | 1 | 1 | 1 | 18.71 | 18.44 |
| **32** | J043 | 36~40 | 1 | 2 | 2 | 2 | 1 | fever, sore throat, muscle ache | 2 | | 15.78 | 15.95 | 17.27 | 1 | 1 | 1 | 1 | 17.14 | 15.61 |
| **33** | J044 | 56~60 | 1 | 2 | 4 | 4 | 1 | cough, rhinorrhea | 2 | | 24.15 | 23.66 | 24.22 | 1 | 1 | 1 | 1 | 17.43 | 17.22 |
| **34** | J045 | 25~29 | 1 | 2 | 3 | 3 | 1 | sore throat | 1 | | 15.94 | 15.52 | 19.08 | 1 | 1 | 1 | 1 | 13.63 | 13.2 |
| **35** | J047 | 49~53 | 2 | 7 | | 7 | 2 | | 1 | | 33.61 | 34.81 | 32.29 | 1 | 2 | 2 | 2 | negative | negative |
| **36** | J048 | 34~38 | 1 | 2 | 3 | 3 | 1 | sputum | 1 | | 13.25 | 14.54 | 16.6 | 1 | 1 | 1 | 1 | 13.41 | 12.73 |
| **37** | J049 | 30~34 | 1 | 3 | >10 | 10 | 1 | decreased taste sense, anosmia | 1 | | 35.33 | 35.46 | 36.64 | 1 | 2 | 2 | 1 | 31.6 | 30.66 |
| **38** | J050 | 74~78 | 2 | 4 | >10 | 10 | 2 | | 2 | | 33.49 | 32.53 | 35.67 | 1 | 2 | 2 | 1 | 31.52 | 30.56 |
| **39** | J051 | 51~55 | 2 | 5 | 12 | 12 | 1 | chills, sputum | 1 | | 33.75 | 33.47 | 36.12 | 1 | 2 | 2 | 1 | 29.12 | 28.24 |
| **40** | J052 | 70~74 | 2 | 5 | 15 | 15 | 1 | cough | 2 | | 35.81 | 35.4 | 35.39 | 1 | 2 | 2 | 2 | negative | negative |
| **41** | J054 | 27~31 | 1 | 2 | 5 | 5 | 1 | cough, fever, cold sweating | 1 | | 16.09 | 16.18 | 18.78 | 1 | 1 | 1 | 1 | 12.74 | 12.04 |
| **42** | J055 | 52~56 | 2 | 3 | 3 | 3 | 2 | | 1 | | 15.22 | 14.78 | 17.7 | 1 | 1 | 1 | 1 | 11.39 | 11.33 |
| **43** | J056 | 35~39 | 1 | 3 | 3 | 3 | 1 | cough, sputum, chills | 2 | | 11.88 | 12.68 | 15.84 | 1 | 1 | 1 | 1 | 10.95 | 10.8 |
| **44** | J057 | 19~23 | 1 | 3 | 3 | 3 | 2 | | 1 | | 25.36 | 25.16 | 28.19 | 1 | 1 | 1 | 1 | 14.37 | 14.37 |
| **45** | J058 | 42~46 | 2 | 2 | 4 | 4 | 1 | cough, sputum | 1 | | 16.2 | 15.96 | 18.85 | 1 | 1 | 1 | 1 | 18.65 | 18.15 |
| **46** | J060 | 50~54 | 2 | 14 | >14 | 14 | 2 | | 2 | | 34.95 | 36.13 | 34.72 | 1 | 2 | 2 | 1 | 35.34 | 34.86 |
| **47** | J061 | 30~34 | 1 | 2 | 3 | 3 | 1 | cough, muscle ache, sputum, rhinorrhea | 1 | | 15.68 | 16.13 | 19.2 | 1 | 1 | 1 | 1 | 13.2 | 13.6 |
| **48** | J062 | 43~47 | 1 | 2 | | 2 | 2 | | 1 | | 12.65 | 14.43 | 16.69 | 1 | 1 | 1 | 1 | 15.81 | 15.05 |
| **49** | J064 | 81~85 | 1 | 5 | 12 | 12 | 2 | | 2 | | 27.55 | 28.21 | 29.25 | 1 | 1 | 1 | 1 | 24.16 | 24.44 |

### Supplementary Appendix 1. Raw Data from 102 Patients with COVID-19

| 1 | 2 | 3 | 4 | 5 | 6 | 7 | 8 | 9 | 10 | 11 | 12 | 13 | 14 | 15 | 16 | 17 | 18 | 19 | 20 |
|---|---|---|---|---|---|---|---|---|---|---|---|---|---|---|---|---|---|---|---|
| **50** | J065 | 52~56 | 2 | 9 | 11 | 11 | 1 | fever, cough | 2 | | 33.08 | 33.73 | 31.98 | 1 | 2 | 2 | 3 | negative | 35.54 |
| **51** | J068 | 37~41 | 2 | 2 | | 2 | 2 | | 2 | | 13.22 | 14.57 | 18.17 | 1 | 1 | 1 | 1 | 22.37 | 21.62 |
| **52** | J069 | 29~33 | 1 | 2 | 2 | 2 | 1 | fatigue, dizness | 1 | | 10.28 | 11.14 | 14.63 | 1 | 1 | 1 | 1 | 15.36 | 14.65 |
| **53** | J070 | 74~78 | 2 | 5 | 8 | 8 | 1 | fever and chills | 2 | | 36.74 | 35.49 | 37.58 | 1 | 2 | 2 | 3 | negative | 35.02 |
| **54** | J071 | 28~32 | 1 | 6 | 6 | 6 | 1 | cough, fever | 2 | | 25.33 | 26.20 | 25.07 | 1 | 1 | 1 | 1 | 15.55 | 15.55 |
| **55** | J072 | 40~44 | 1 | 2 | 2 | 2 | 1 | sore throat, rhinorrhea | 2 | | 15.48 | 15.30 | 18.6 | 1 | 1 | 1 | 1 | 14.03 | 13.2 |
| **56** | J073 | 37~41 | 1 | 2 | 2 | 2 | 1 | headache | 1 | | 19.97 | 19.48 | 22.45 | 1 | 1 | 1 | 1 | 12.83 | 12.35 |
| **57** | J074 | 34~38 | 1 | 2 | 2 | 2 | 1 | cough | 1 | | 16.2 | 16.51 | 19.09 | 1 | 1 | 1 | 1 | 23.34 | 21.82 |
| **58** | J075 | 35~39 | 1 | 2 | 2 | 2 | 1 | cough, muscle ache | 1 | | 15.75 | 15.74 | 18.53 | 1 | 1 | 1 | 1 | 17.21 | 15.74 |
| **59** | J081 | 50~54 | 1 | 4 | 8 | 8 | 1 | cough | 2 | | 21.03 | 20.54 | 24.21 | 1 | 1 | 1 | 1 | 23.38 | 23 |
| **60** | J086 | 29~33 | 2 | 7 | 8 | 8 | 1 | rhinorrhea, headache | 1 | | 35.56 | 38.91 | 33.52 | 1 | 2 | 2 | 3 | negative | 34.49 |
| **61** | J093 | 18~22 | 1 | 6 | 6 | 6 | 1 | headache, sore throat, cough | 1 | | 18.16 | 19.12 | 18.61 | 1 | 1 | 1 | 1 | 18.36 | 16.99 |
| **62** | J094 | 70~74 | 1 | 6 | 6 | 6 | 2 | | 1 | | 24.00 | 24.72 | 23.71 | 1 | 1 | 1 | 1 | 18.21 | 17.00 |
| **63** | J096 | 42~46 | 2 | 6 | 6 | 6 | 1 | chills, cough | 1 | | 16.64 | 17.81 | 16.74 | 1 | 1 | 1 | 1 | 18.28 | 17.07 |
| **64** | J097 | 46~50 | 2 | 6 | 9 | 9 | 1 | headache, chills, fever | 1 | | 20.17 | 21.01 | 20.41 | 1 | 1 | 1 | 1 | 19.80 | 18.63 |
| **65** | J101 | 41~45 | 2 | 3 | 8 | 8 | 1 | cough, sputum, rhinorrhea | 1 | | 18.74 | 18.81 | 22.24 | 1 | 1 | 1 | 1 | 27.38 | 25.54 |
| **66** | J105 | 22~26 | 2 | 7 | 10 | 10 | 1 | sore throat, cough, nasal stuffness | 1 | | 31.35 | 33.08 | 31.21 | 1 | 2 | 2 | 2 | negative | negative |
| **67** | J107 | 57~61 | 1 | 9 | >10 | 10 | 2 | | 1 | | 28.79 | 30.09 | 28.73 | 1 | 2 | 2 | 1 | 31.51 | 29.40 |
| **68** | J116 | 24~28 | 2 | 3 | 4 | 4 | 1 | chills, cough, headache | 1 | | 10.64 | 11.84 | 16.87 | 1 | 1 | 1 | 1 | 21.11 | 20.68 |
| **69** | J117 | 21~25 | 2 | 3 | 6 | 6 | 1 | sore throat, abdominal pain, diarrhea | 1 | | 20.61 | 20.37 | 24.58 | 1 | 1 | 1 | 1 | 12.74 | 12.79 |
| **70** | J121 | 34~38 | 1 | 7 | 9 | 9 | 1 | cough, muscle ache | 1 | | 18.48 | 19.54 | 18.65 | 1 | 1 | 1 | 1 | 24.50 | 24.05 |
| **71** | J122 | 41~45 | 2 | 5 | 5 | 5 | 1 | dizness, sputum, fever, nasal stuffness | 2 | | 26.74 | 27.51 | 30.08 | 1 | 1 | 1 | 1 | 20.72 | 19.70 |
| **72** | J123 | 27~31 | 1 | 6 | | 7 | 2 | 4/12, AstraZeneca first shot | 1 | | 34.47 | 34.01 | 36.96 | 1 | 2 | 2 | 3 | negative | 34.67 |

### Supplementary Appendix 1. Raw Data from 102 Patients with COVID-19

| 1 | 2 | 3 | 4 | 5 | 6 | 7 | 8 | 9 | 10 | 11 | 12 | 13 | 14 | 15 | 16 | 17 | 18 | 19 | 20 |
|---|---|---|---|---|---|---|---|---|---|---|---|---|---|---|---|---|---|---|---|
| **73** | J126 | 55~59 | 1 | 4 | 6 | 6 | 1 | chills, cough, sore throat | 1 | | 20.06 | 19.95 | 22.49 | 1 | 1 | 1 | 1 | 16.53 | 15.71 |
| **74** | J127 | 50~54 | 1 | 5 | 6 | 6 | 1 | cough | 1 | | 19.29 | 18.98 | 22.25 | 1 | 1 | 1 | 1 | 12.99 | 12.39 |
| **75** | J129 | 21~25 | 2 | 9 | 12 | 12 | 1 | sore throat, abdominal pain, diarrhea | 1 | | 29.72 | 29.58 | 32.44 | 1 | 2 | 2 | 1 | 26.65 | 25.62 |
| **76** | J130 | 55~59 | 2 | 7 | 10 | 10 | 1 | chills, muscle ache | 1 | | 34.88 | 35.04 | 32.97 | 1 | 2 | 2 | 2 | negative | negative |
| **77** | J131 | 31~35 | 2 | 6 | 7 | 7 | 1 | headache, muscle ache | 1 | | 27.17 | 27.18 | 26.31 | 1 | 1 | 1 | 1 | 16.13 | 15.62 |
| **78** | J144 | 46~50 | 1 | 9 | 10 | 10 | 1 | cough, muscle ache, fever | 1 | | 29.95 | 30.70 | 31.90 | 1 | 2 | 2 | 1 | 32.10 | 30.09 |
| **79** | J145 | 55~59 | 1 | 7 | 13 | 13 | 1 | sputum, sore throat | 1 | | 24.73 | 34.37 | 28.64 | 1 | 1 | 1 | 1 | 24.51 | 22.89 |
| **80** | J146 | 67~71 | 1 | 11 | 12 | 12 | 1 | muscle ache | 1 | | 24.71 | 24.56 | 27.66 | 1 | 2 | 2 | 1 | 29.52 | 27.01 |
| **81** | J147 | 44~48 | 1 | 11 | 12 | 12 | 1 | fever, entry from Cambodia | 2 | | 35.82 | 38.11 | 35.94 | 1 | 2 | 2 | 1 | 34.44 | 32.41 |
| **82** | J148 | 40~44 | 1 | 13 | 14 | 14 | 1 | fever, sputum, 5/27, AstraZeneca first shot | 1 | | 28.85 | 29.17 | 32.65 | 1 | 1 | 1 | 1 | 17.40 | 16.06 |
| **83** | J149 | 25~29 | 2 | 12 | 12 | 12 | 1 | cough, sputum, nasal stufness, diarrhea, muscle ache, fever | 1 | | 31.66 | 31.25 | 33.91 | 1 | 2 | 2 | 2 | negative | negative |
| **84** | J150 | 50~54 | 2 | 6 | 6 | 6 | 1 | cough, sputum, fever | 1 | | 22.71 | 23.11 | 24.70 | 1 | 1 | 1 | 1 | 24.10 | 21.82 |
| **85** | J151 | 46~50 | 2 | 5 | 9 | 9 | 2 | | 1 | | 30.90 | 31.30 | 33.10 | 1 | 2 | 2 | 1 | 27.34 | 25.61 |
| **86** | J152 | 46~50 | 2 | 11 | 13 | 13 | 1 | headache, cough | 1 | | 31.25 | 31.79 | 34.32 | 1 | 2 | 2 | 1 | 27.11 | 25.20 |
| **87** | J153 | 20~24 | 1 | 13 | 15 | 15 | 1 | fever | 1 | | 31.78 | 32.64 | 31.67 | 1 | 2 | 2 | 2 | negative | negative |
| **88** | J154 | 56~60 | 1 | 12 | 15 | 15 | 1 | common cold sense | 2 | | 34.65 | 37.05 | 36.33 | 1 | 2 | 2 | 2 | negative | negative |
| **89** | J155 | 44~48 | 2 | 14 | 14 | 14 | 1 | fever | 1 | | 27.67 | 28.67 | 27.53 | 1 | 2 | 2 | 1 | 30.20 | 28.40 |
| **90** | J157 | 38~42 | 1 | 7 | 12 | 12 | 1 | chills, common cold sense | 1 | | 28.92 | 29.12 | 31.42 | 1 | 2 | 2 | 1 | 24.79 | 23.17 |
| **91** | J158 | 59~63 | 1 | 3 | 13 | 13 | 2 | | 1 | | 34.66 | 34.20 | 32.29 | 1 | 2 | 2 | 3 | negative | 33.28 |
| **92** | J159 | 20~24 | 1 | 3 | 10 | 10 | 1 | fever, cough, diarrhea, body weght loss (5kg) | 1 | | 33.68 | 34.87 | 32.95 | 1 | 2 | 2 | 3 | negative | 33.23 |

### Supplementary Appendix 1. Raw Data from 102 Patients with COVID-19

| 1 | 2 | 3 | 4 | 5 | 6 | 7 | 8 | 9 | 10 | 11 | 12 | 13 | 14 | 15 | 16 | 17 | 18 | 19 | 20 |
|---|---|---|---|---|---|---|---|---|---|---|---|---|---|---|---|---|---|---|---|
| **93** | J160 | 62~66 | 2 | 2 | 9 | 9 | 1 | chills, muscle ache, anosmia, headache | 2 | | 27.18 | 27.79 | 30.50 | 1 | 2 | 2 | 1 | 27.30 | 26.19 |
| **94** | J161 | 44~48 | 1 | 2 | 11 | 11 | 1 | nausea, anosmia | 1 | | 30.82 | 31.94 | 30.61 | 1 | 2 | 2 | 1 | 33.84 | 33.28 |
| **95** | J162 | 37~41 | 1 | 4 | 6 | 6 | 1 | muscle ache, fever, cold sweating, entry from Russia | 2 | 2 | 18.84 | 19.25 | 19.37 | 1 | 1 | 1 | 1 | 17.85 | 17.52 |
| **96** | J165 | 35~39 | 1 | 6 | 12 | 12 | 1 | fever, cough, headache | 1 | | 21.79 | 22.62 | 21.87 | 1 | 2 | 2 | 1 | 23.85 | 23.36 |
| **97** | J166 | 44~48 | 1 | 6 | 12 | 12 | 2 | | 1 | | 22.95 | 24.28 | 22.47 | 1 | 2 | 2 | 1 | 30.24 | 29.53 |
| **98** | J167 | 23~27 | 2 | 3 | 6 | 6 | 1 | chills, cough | 2 | | 25.39 | 26.59 | 24.99 | 1 | 2 | 2 | 1 | 29.18 | 28.42 |
| **99** | J168 | 16~20 | 1 | 6 | 9 | 9 | 2 | | 1 | | 19.15 | 18.92 | 22.07 | 1 | 2 | 2 | 1 | 26.01 | 25.14 |
| **100** | J171 | 16~20 | 1 | 7 | 8 | 8 | 1 | fever, rhinorrhea | 1 | | 22.62 | 23.98 | 23.06 | 1 | 2 | 2 | 1 | 27.46 | 26.73 |
| **101** | J173 | 43~47 | 2 | 4 | 5 | 5 | 1 | headache, sore throat, fever sensation | 1 | 2 | 16.93 | 17.41 | 17.21 | 1 | 1 | 1 | 1 | 18.82 | 18.64 |
| **102** | J174 | 22~26 | 2 | 3 | 3 | 3 | 1 | headache, eye pain, fever, cough | 1 | 2 | 12.59 | 12.94 | 17.74 | 1 | 1 | 1 | 1 | 16.42 | 16.35 |

### Supplementary Appendix 2. Raw Data from 100 Healthy Individuals as Negative Control

| No. | Clinical Trial Healthy Control Number | Sex (Man=1, Woman=2) | Age range (years) | NPS_PCR _Ct value E gene | NPS_PCR _Ct value RdRP gene | NPS_PCR _Ct value N gene | NPS_PCR _Result (Positive=1, Negative=2) | Antigen RDT [STANDARD Q COVID-19 Ag Saliva test (SD BIOSENSOR)] (Positive=1, Negative=2) | Antigen RDT [Gmate® COVID-19 Ag Saliva (Philosys)] (Positive=1, Negative=2) | Saliva_PCR _Result (Positive=1, Negative=2, Inconclusive=3) | Saliva_PCR _Ct value RdRP gene | Saliva_PCR _Ct value E gene |
|---|---|---|---|---|---|---|---|---|---|---|---|---|
| 1 | N001 | 2 | 55~59 | negative | negative | negative | 2 | 2 | 2 | 2 | negative | negative |
| 2 | N002 | 1 | 37~41 | negative | negative | negative | 2 | 2 | 2 | 2 | negative | negative |
| 3 | N003 | 2 | 44~48 | negative | negative | negative | 2 | 2 | 2 | 2 | negative | negative |
| 4 | N004 | 2 | 23~27 | negative | negative | negative | 2 | 2 | 2 | 2 | negative | negative |
| 5 | N005 | 2 | 45~49 | negative | negative | negative | 2 | 2 | 2 | 2 | negative | negative |
| 6 | N006 | 1 | 54~58 | negative | negative | negative | 2 | 2 | 2 | 2 | negative | negative |
| 7 | N007 | 2 | 54~58 | negative | negative | negative | 2 | 2 | 2 | 2 | negative | negative |
| 8 | N008 | 2 | 39~43 | negative | negative | negative | 2 | 2 | 2 | 2 | negative | negative |
| 9 | N009 | 2 | 23~27 | negative | negative | negative | 2 | 2 | 2 | 2 | negative | negative |
| 10 | N010 | 2 | 50~54 | negative | negative | negative | 2 | 2 | 2 | 2 | negative | negative |
| 11 | N011 | 2 | 51~55 | negative | negative | negative | 2 | 2 | 2 | 2 | negative | negative |
| 12 | N012 | 2 | 22~26 | negative | negative | negative | 2 | 2 | 2 | 2 | negative | negative |
| 13 | N013 | 2 | 48~52 | negative | negative | negative | 2 | 2 | 2 | 2 | negative | negative |
| 14 | N014 | 1 | 42~46 | negative | negative | negative | 2 | 2 | 2 | 2 | negative | negative |
| 15 | N015 | 2 | 29~33 | negative | negative | negative | 2 | 2 | 2 | 2 | negative | negative |
| 16 | N016 | 2 | 28~32 | negative | negative | negative | 2 | 2 | 2 | 2 | negative | negative |
| 17 | N017 | 1 | 49~53 | negative | negative | negative | 2 | 2 | 2 | 2 | negative | negative |
| 18 | N018 | 2 | 49~53 | negative | negative | negative | 2 | 2 | 2 | 2 | negative | negative |
| 19 | N019 | 2 | 46~50 | negative | negative | negative | 2 | 2 | 2 | 2 | negative | negative |
| 20 | N020 | 2 | 48~52 | negative | negative | negative | 2 | 2 | 2 | 2 | negative | negative |
| 21 | N021 | 2 | 23~27 | negative | negative | negative | 2 | 2 | 2 | 2 | negative | negative |
| 22 | N022 | 2 | 24~28 | negative | negative | negative | 2 | 2 | 2 | 2 | negative | negative |
| 23 | N023 | 2 | 41~45 | negative | negative | negative | 2 | 2 | 2 | 2 | negative | negative |

### Supplementary Appendix 2. Raw Data from 100 Healthy Individuals as Negative Control

| No. | Clinical Trial Healthy Control Number | Sex (Man=1, Woman=2) | Age range (years) | NPS_PCR _Ct value E gene | NPS_PCR _Ct value RdRP gene | NPS_PCR _Ct value N gene | NPS_PCR _Result (Positive=1, Negative=2) | Antigen RDT [STANDARD Q COVID-19 Ag Saliva test (SD BIOSENSOR)] (Positive=1, Negative=2) | Antigen RDT [Gmate® COVID-19 Ag Saliva (Philosys)] (Positive=1, Negative=2) | Saliva_PCR _Result (Positive=1, Negative=2, Inconclusive=3) | Saliva_PCR _Ct value RdRP gene | Saliva_PCR _Ct value E gene |
|---|---|---|---|---|---|---|---|---|---|---|---|---|
| 24 | N024 | 2 | 46~50 | negative | negative | negative | 2 | 2 | 2 | 2 | negative | negative |
| 25 | N025 | 1 | 46~50 | negative | negative | negative | 2 | 2 | 2 | 2 | negative | negative |
| 26 | N026 | 2 | 41~45 | negative | negative | negative | 2 | 2 | 2 | 2 | negative | negative |
| 27 | N027 | 2 | 23~27 | negative | negative | negative | 2 | 2 | 2 | 2 | negative | negative |
| 28 | N028 | 2 | 47~51 | negative | negative | negative | 2 | 2 | 2 | 2 | negative | negative |
| 29 | N029 | 2 | 22~26 | negative | negative | negative | 2 | 2 | 2 | 2 | negative | negative |
| 30 | N030 | 2 | 45~49 | negative | negative | negative | 2 | 2 | 2 | 2 | negative | negative |
| 31 | N031 | 2 | 38~42 | negative | negative | negative | 2 | 2 | 2 | 2 | negative | negative |
| 32 | N032 | 2 | 45~49 | negative | negative | negative | 2 | 2 | 2 | 2 | negative | negative |
| 33 | N033 | 2 | 54~58 | negative | negative | negative | 2 | 2 | 2 | 2 | negative | negative |
| 34 | N034 | 1 | 51~55 | negative | negative | negative | 2 | 2 | 2 | 2 | negative | negative |
| 35 | N035 | 2 | 54~58 | negative | negative | negative | 2 | 2 | 2 | 2 | negative | negative |
| 36 | N036 | 2 | 50~54 | negative | negative | negative | 2 | 2 | 2 | 2 | negative | negative |
| 37 | N037 | 2 | 48~52 | negative | negative | negative | 2 | 2 | 2 | 2 | negative | negative |
| 38 | N038 | 2 | 39~43 | negative | negative | negative | 2 | 2 | 2 | 2 | negative | negative |
| 39 | N039 | 2 | 41~45 | negative | negative | negative | 2 | 2 | 2 | 2 | negative | negative |
| 40 | N040 | 2 | 38~42 | negative | negative | negative | 2 | 2 | 2 | 2 | negative | negative |
| 41 | N041 | 2 | 41~45 | negative | negative | negative | 2 | 2 | 2 | 2 | negative | negative |
| 42 | N042 | 2 | 47~51 | negative | negative | negative | 2 | 2 | 2 | 2 | negative | negative |
| 43 | N043 | 2 | 41~45 | negative | negative | negative | 2 | 2 | 2 | 2 | negative | negative |
| 44 | N044 | 1 | 42~46 | negative | negative | negative | 2 | 2 | 2 | 2 | negative | negative |
| 45 | N045 | 1 | 34~38 | negative | negative | negative | 2 | 2 | 2 | 2 | negative | negative |
| 46 | N046 | 2 | 23~27 | negative | negative | negative | 2 | 2 | 2 | 2 | negative | negative |

### Supplementary Appendix 2. Raw Data from 100 Healthy Individuals as Negative Control

| No. | Clinical Trial Healthy Control Number | Sex (Man=1, Woman=2) | Age range (years) | NPS_PCR _Ct value E gene | NPS_PCR _Ct value RdRP gene | NPS_PCR _Ct value N gene | NPS_PCR _Result (Positive=1, Negative=2) | Antigen RDT [STANDARD Q COVID-19 Ag Saliva test (SD BIOSENSOR)] (Positive=1, Negative=2) | Antigen RDT [Gmate® COVID-19 Ag Saliva (Philosys)] (Positive=1, Negative=2) | Saliva_PCR _Result (Positive=1, Negative=2, Inconclusive=3) | Saliva_PCR _Ct value RdRP gene | Saliva_PCR _Ct value E gene |
|---|---|---|---|---|---|---|---|---|---|---|---|---|
| 47 | N047 | 2 | 22~26 | negative | negative | negative | 2 | 2 | 2 | 2 | negative | negative |
| 48 | N048 | 1 | 27~31 | negative | negative | negative | 2 | 2 | 2 | 2 | negative | negative |
| 49 | N049 | 2 | 22~26 | negative | negative | negative | 2 | 2 | 2 | 2 | negative | negative |
| 50 | N050 | 1 | 24~28 | negative | negative | negative | 2 | 2 | 2 | 2 | negative | negative |
| 51 | N051 | 1 | 24~28 | negative | negative | negative | 2 | 2 | 2 | 2 | negative | negative |
| 52 | N052 | 2 | 30~34 | negative | negative | negative | 2 | 2 | 2 | 2 | negative | negative |
| 53 | N053 | 2 | 23~27 | negative | negative | negative | 2 | 2 | 2 | 2 | negative | negative |
| 54 | N054 | 2 | 26~30 | negative | negative | negative | 2 | 2 | 2 | 2 | negative | negative |
| 55 | N055 | 1 | 53~57 | negative | negative | negative | 2 | 2 | 2 | 2 | negative | negative |
| 56 | N056 | 1 | 28~32 | negative | negative | negative | 2 | 2 | 2 | 2 | negative | negative |
| 57 | N057 | 1 | 29~33 | negative | negative | negative | 2 | 2 | 2 | 2 | negative | negative |
| 58 | N058 | 1 | 29~33 | negative | negative | negative | 2 | 2 | 2 | 2 | negative | negative |
| 59 | N059 | 1 | 56~60 | negative | negative | negative | 2 | 2 | 2 | 2 | negative | negative |
| 60 | N060 | 2 | 57~61 | negative | negative | negative | 2 | 2 | 2 | 2 | negative | negative |
| 61 | N061 | 2 | 39~43 | negative | negative | negative | 2 | 2 | 2 | 2 | negative | negative |
| 62 | N062 | 2 | 28~32 | negative | negative | negative | 2 | 2 | 2 | 2 | negative | negative |
| 63 | N063 | 1 | 37~41 | negative | negative | negative | 2 | 2 | 2 | 2 | negative | negative |
| 64 | N064 | 1 | 27~31 | negative | negative | negative | 2 | 2 | 2 | 2 | negative | negative |
| 65 | N065 | 1 | 36~40 | negative | negative | negative | 2 | 2 | 2 | 2 | negative | negative |
| 66 | N066 | 1 | 30~34 | negative | negative | negative | 2 | 2 | 2 | 2 | negative | negative |
| 67 | N067 | 2 | 25~29 | negative | negative | negative | 2 | 2 | 2 | 2 | negative | negative |
| 68 | N068 | 1 | 29~33 | negative | negative | negative | 2 | 2 | 2 | 2 | negative | negative |
| 69 | N069 | 2 | 53~57 | negative | negative | negative | 2 | 2 | 2 | 2 | negative | negative |

### Supplementary Appendix 2. Raw Data from 100 Healthy Individuals as Negative Control

| No. | Clinical Trial Healthy Control Number | Sex (Man=1, Woman=2) | Age range (years) | NPS_PCR _Ct value E gene | NPS_PCR _Ct value RdRP gene | NPS_PCR _Ct value N gene | NPS_PCR _Result (Positive=1, Negative=2) | Antigen RDT [STANDARD Q COVID-19 Ag Saliva test (SD BIOSENSOR)] (Positive=1, Negative=2) | Antigen RDT [Gmate® COVID-19 Ag Saliva (Philosys)] (Positive=1, Negative=2) | Saliva_PCR _Result (Positive=1, Negative=2, Inconclusive=3) | Saliva_PCR _Ct value RdRP gene | Saliva_PCR _Ct value E gene |
|---|---|---|---|---|---|---|---|---|---|---|---|---|
| 70 | N070 | 2 | 31~35 | negative | negative | negative | 2 | 2 | 2 | 2 | negative | negative |
| 71 | N071 | 1 | 37~41 | negative | negative | negative | 2 | 2 | 2 | 2 | negative | negative |
| 72 | N072 | 2 | 38~42 | negative | negative | negative | 2 | 2 | 2 | 2 | negative | negative |
| 73 | N073 | 2 | 22~26 | negative | negative | negative | 2 | 2 | 2 | 2 | negative | negative |
| 74 | N074 | 2 | 36~40 | negative | negative | negative | 2 | 2 | 2 | 2 | negative | negative |
| 75 | N075 | 1 | 52~56 | negative | negative | negative | 2 | 2 | 2 | 2 | negative | negative |
| 76 | N076 | 2 | 43~47 | negative | negative | negative | 2 | 2 | 2 | 2 | negative | negative |
| 77 | N077 | 2 | 43~47 | negative | negative | negative | 2 | 2 | 2 | 2 | negative | negative |
| 78 | N078 | 2 | 31~35 | negative | negative | negative | 2 | 2 | 2 | 2 | negative | negative |
| 79 | N079 | 1 | 56~60 | negative | negative | negative | 2 | 2 | 2 | 2 | negative | negative |
| 80 | N080 | 2 | 29~33 | negative | negative | negative | 2 | 2 | 2 | 2 | negative | negative |
| 81 | N081 | 2 | 46~50 | negative | negative | negative | 2 | 2 | 2 | 2 | negative | negative |
| 82 | N082 | 2 | 46~50 | negative | negative | negative | 2 | 2 | 2 | 2 | negative | negative |
| 83 | N083 | 1 | 48~52 | negative | negative | negative | 2 | 2 | 2 | 2 | negative | negative |
| 84 | N084 | 2 | 37~41 | negative | negative | negative | 2 | 2 | 2 | 2 | negative | negative |
| 85 | N085 | 2 | 36~40 | negative | negative | negative | 2 | 2 | 2 | 2 | negative | negative |
| 86 | N086 | 2 | 25~29 | negative | negative | negative | 2 | 2 | 2 | 2 | negative | negative |
| 87 | N087 | 2 | 21~25 | negative | negative | negative | 2 | 2 | 2 | 2 | negative | negative |
| 88 | N088 | 2 | 34~38 | negative | negative | negative | 2 | 2 | 2 | 2 | negative | negative |
| 89 | N089 | 2 | 24~28 | negative | negative | negative | 2 | 2 | 2 | 2 | negative | negative |
| 90 | N090 | 1 | 42~46 | negative | negative | negative | 2 | 2 | 2 | 2 | negative | negative |
| 91 | N091 | 2 | 26~30 | negative | negative | negative | 2 | 2 | 2 | 2 | negative | negative |
| 92 | N092 | 2 | 24~28 | negative | negative | negative | 2 | 2 | 2 | 2 | negative | negative |

### Supplementary Appendix 2. Raw Data from 100 Healthy Individuals as Negative Control

| No. | Clinical Trial Healthy Control Number | Sex (Man=1, Woman=2) | Age range (years) | NPS_PCR _Ct value E gene | NPS_PCR _Ct value RdRP gene | NPS_PCR _Ct value N gene | NPS_PCR _Result (Positive=1, Negative=2) | Antigen RDT [STANDARD Q COVID-19 Ag Saliva test (SD BIOSENSOR)] (Positive=1, Negative=2) | Antigen RDT [Gmate® COVID-19 Ag Saliva (Philosys)] (Positive=1, Negative=2) | Saliva_PCR _Result (Positive=1, Negative=2, Inconclusive=3) | Saliva_PCR _Ct value RdRP gene | Saliva_PCR _Ct value E gene |
|---|---|---|---|---|---|---|---|---|---|---|---|---|
| 93 | N093 | 1 | 26~30 | negative | negative | negative | 2 | 2 | 2 | 2 | negative | negative |
| 94 | N094 | 1 | 58~62 | negative | negative | negative | 2 | 2 | 2 | 2 | negative | negative |
| 95 | N095 | 2 | 58~62 | negative | negative | negative | 2 | 2 | 2 | 2 | negative | negative |
| 96 | N096 | 2 | 49~53 | negative | negative | negative | 2 | 2 | 2 | 2 | negative | negative |
| 97 | N097 | 1 | 56~60 | negative | negative | negative | 2 | 2 | 2 | 2 | negative | negative |
| 98 | N098 | 1 | 38~42 | negative | negative | negative | 2 | 2 | 2 | 2 | negative | negative |
| 99 | N099 | 1 | 40~44 | negative | negative | negative | 2 | 2 | 2 | 2 | negative | negative |
| 100 | N100 | 2 | 56~60 | negative | negative | negative | 2 | 2 | 2 | 2 | negative | negative |
