## Supplement 2 for "Bean extract-based gargle for efficient diagnosing COVID-19 at early-stage using rapid antigen tests : a clinical, prospective, diagnostic study"

This appendix has been provided by the authors to give readers additional information about their work.

Supplement to: Joseph Kwon, Euna Ko, Se-Young Cho, et al. Bean extract-based gargle as promising tool for efficient diagnosis of COVID-19 at early-stage using rapid antigen tests: a clinical, prospective, diagnostic sensitivity study

### TABLE OF CONTENTS

**Study design** 4

**Inclusion and exclusion criteria of study participants** 5

**Supplementary Methods** 6

Chemicals and reagents 6
Cryo-electron microscopy 6
Enzyme-linked immunosorbent assay (ELISA) to test RBD binding inhibition to hACE2 receptors 6
Preparation and Purification of TCan 6
On-line Top-down Analysis of TCan using UPLC-ESI-QTOF (top-down) 6
Protein analysis using Nano UPLC-QTOF for TCan analysis (bottom-up) 7
N-Terminal sequencing of TCan 7
Preparation of SARS-CoV-2 spike protein 7
Circular dichroism spectroscopy 7
Dynamic and light scattering measurement 8
Intrinsic and extrinsic fluorescence spectroscopy 8
Differential scanning calorimetry 8
Isothermal titration calorimetry 8
Size exclusion chromatography-combined multi-angle light scattering measurement 9
Cell culture, viruses, and treatment 9
Determination of cell viability 9
Measurement of reactive oxygen species (ROS) levels 9
Primary splenocyte preparation and culture 9
Animals 10
Experimental design for *in vivo* study 10
Intratracheal and intravenous instillation 10
Bronchoalveolar lavage fluid preparation 10
Measurement of cytokine levels 10
Histological analysis 11

**Supplementary Study Results and Discussion** 12

**Supplementary Figures** 17

**eFigure 1.** Comparison of the Ct values for BG-RT-PCR with those for NPS-RT-PCR 17
**eFigure 2.** Transmission electron micrographs (TEM) of HCoV-229E and SARS-CoV-2 with bean extract (BE) treatment 18
**eFigure 3.** The profile of the TCan from size exclusion chromatography 19
**eFigure 4**. Multiple-charge electrospray mass spectra of TCan 20
**eFigure 5.** Molecular characterization of TCan using various biophysical approaches 21
**eFigure 6.** Structural characterization of TCan using spectroscopies 22
**eFigure 7.** Intramolecular characterization of TCan 23
**eFigure 8.** Irreversibility of the unfolding reaction of TCan after heat scanning 24
**eFigure 9.** Changes in the expression levels of inflammatory cytokines 25
**eFigure 10.** Effect of Con A and BE instillation on survival rate of C57BL/6 mice 26
**eFigure 11.** Changes in the levels of hepatic biomarker (AST, ALT, TBIL, and GGT) in response to Con A and BE instillation in mice 27
**eFigure 12.** Changes in lung and spleen weight-to-body weight ratio after Con A and BE instillation in mice 28
**eFigure 13.** Changes of inflammatory cell numbers in bronchoalveolar lavage fluid (BALF) of mice following Con A and BE instillation 29
**eFigure 14.** Changes in the expression of inflammatory cytokines 30

**eFigure 15.** *In vitro* cytotoxicity of Con A and BE ··· **31**
**eFigure 16.** Reactive oxygen species (ROS) level in cells after Con A and BE treatment ··· **32**

#### Supplementary Tables ··· 33

**eTable 1.** Evaluation of the diagnostic performance of BG-RT-PCR and BG-Ag-RDTs for patients with COVID-19 throughout the infection duration (n=102) ··· **33**
**eTable 2.** Results for the BG-RT-PCR and BG-Ag-RDTs from negative controls (n=100) and COVID-19 patients within 6 days of illness (n=45) ··· **34**
**eTable 3.** Evaluation of the diagnostic performance of BG-RT-PCR and BG-Ag-RDTs for patients of SARS-CoV-2 variants within 6 days of illness ··· **35**
**eTable 4.** Sensitivity of BG-Ag-RDTs depending on Ct value of positive BG-RT-PCR samples ··· **36**
**eTable 5.** Summary of thermodynamic parameters for the structural stability of TCan by circular dichroism spectroscopy ··· **37**
**eTable 6.** Summary of thermodynamic parameters for the structural stability of TCan by differential scanning calorimetry ··· **37**

#### Supplementary References ··· 38

#### Study design

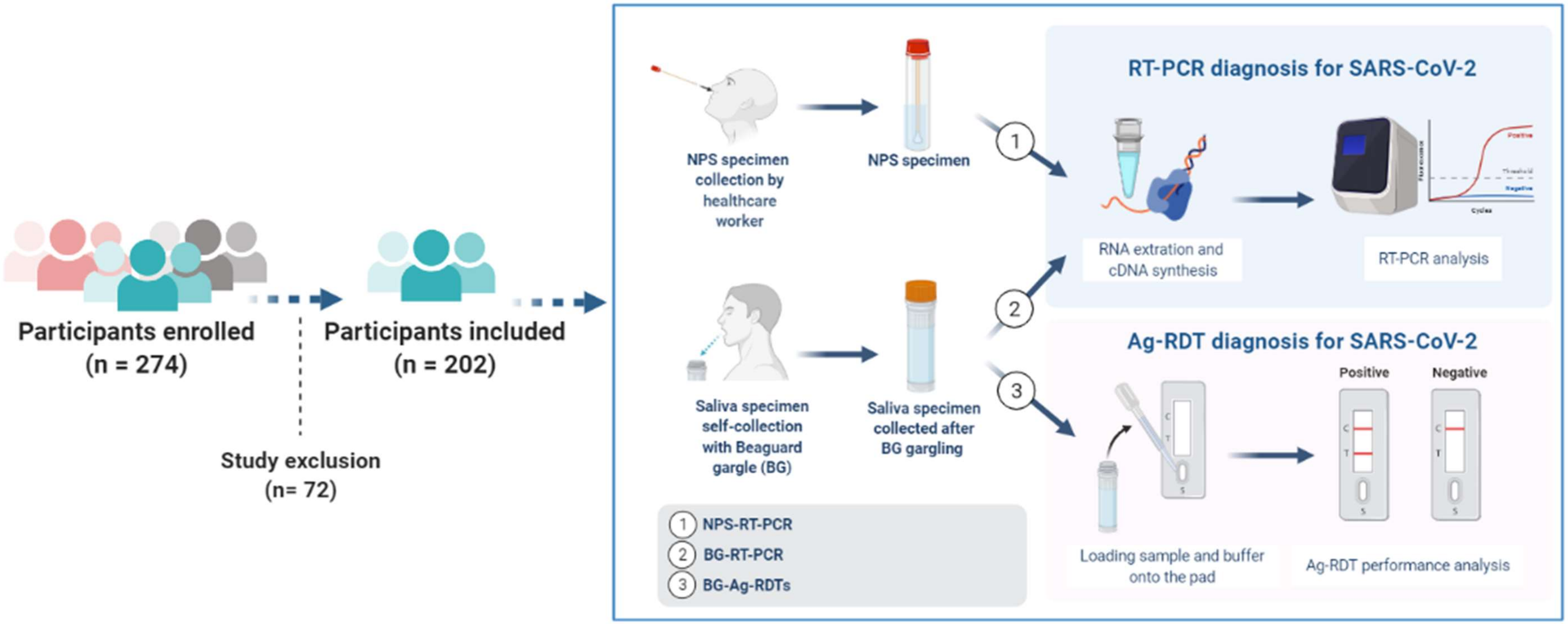

#### Inclusion and exclusion criteria of study participants

*Inclusion criteria*

(a) Adults aged ≥ 18 years

(i) Patients hospitalized with COVID-19 confirmation in the screening center.

(ii) Healthy subjects with no previous contact with confirmed or suspected cases in the two weeks prior to the trial.

(b) Agree to participate in this trial and provide written informed consent.

(c) Permit the investigator to access the patient's medical records relevant to study procedures.

(d) Provide paired NPS and BG-based saliva specimen. Medical professionals collected NPS specimens from participants by spitting into a tube after swirling and gargling 5 mL of BG for 2 min.

*Exclusion criteria*

(a) Healthy subjects with a body temperature higher than 37.0 ºC

(b) Healthy subjects with respiratory symptoms

(c) Not willing to provide sufficient BG-based saliva specimen.

(d) Not willing to collect NPS specimen.

#### Supplementary Methods

##### Chemicals and reagents

Beanguard™ gargle (BG) was supplied by BIO3S, Inc. (Gwangju, Republic of Korea). Roswell Park Memorial Institute (RPMI) 1640, Dulbecco's modified Eagle's medium (DMEM), fetal bovine serum (FBS), trypsin-EDTA, phosphate-buffered saline (PBS), and streptomycin/penicillin were purchased from Gibco (New York, USA); 2′,7′-dichloro-fluorescein diacetate (DCF-DA) was purchased from Thermo Fisher Scientific (California, USA); 1-anilinonaphthalene-8-sulfonate ammonium salt (ANS) was purchased from Sigma-Aldrich (St. Louis, MO, USA). Heat inactivated SARS-CoV-2 culture fluid (Isolate: Italy-INMI1) virus particles were obtained from Zeptometrix (Buffalo, NY). The SARS-CoV-2 spike (S) protein S1 was obtained from ACROBiosystems (Delaware, USA). Recombinant S2 was expressed and purified as described below. The concentrations of S1 and S2 were determined based on the UV absorbance at 280 nm with molar extinction coefficients of 91,775 and 42,455/ M cm, respectively, and were calculated using the ExPASy ProtParam tool.[1]

##### Cryo-electron microscopy (cryo-EM)

The purified whole virus, SARS-CoV-2, and a mixture of SARS-CoV-2 and bean extract (BE) samples were inactivated with 2% paraformaldehyde overnight at 4 °C. Four microliters of each sample was applied to R1.2/1.3 Quantifoil holey carbon EM grids (200 mesh), which were glow-discharged for 60 s at 20 mA. The grids were then plunge-frozen in liquid ethane using a Vitrobot Mark IV (ThermoFisher Scientific) with 5 s of blotting in 100% humidity at 4 °C. Cryo-EM images were acquired using a Titan Krios (ThermoFisher Scientific), operated at 300 kV, and equipped with a Falcon III direct detector.

##### Enzyme-linked immunosorbent assay (ELISA) to test inhibition of RBD binding to hACE2 receptors

To assess the dissociation activity of the receptor binding domain (RBD)–human version of angiotensin-converting enzyme 2 (hACE2) complex by BE, COVID-19 Neutralizing Antibody ELISA Kit (Abnova, Taiwan) was used according to the manufacturer's instructions, with slight modifications. Briefly, the hACE2 protein was coated on 96-well ELISA plates at 4 °C overnight and unbound proteins were removed with washing buffer, followed by blocking for 1 h at 37 °C. For pre-treatment, RBD and BE solutions were incubated for 2 h at 37 °C before reaction with hACE2-coated plate. For post-treatment, COVID-19 Spike RBD Rabbit Fc-Tag protein was added to the plates and incubated for 2 h at 37 °C, followed by washing two times with each successive 2-fold dilution of BE solution. To detect bound RBD-Fc proteins in the plate, it was washed with a wash buffer provided in the ELISA kit, and horseradish peroxidase (HRP)-conjugated goat anti-rabbit antibody was added to each well and the plate was incubated for 1 h at 37 °C. The plate was then washed with wash buffer and subsequently treated with 3,3′,5,5′-tetramethylbenzidine (TMB). Color development was terminated using the stop solution in the ELISA kit. Absorbance at 450 nm was measured using a microplate reader (Bio Tek Co., USA). The percentage of RBD–hACE2 binding was calculated by measuring the differences in the amount of labeled RBD between the test and control samples.

##### Preparation and purification of TCan

For the preparation of truncated canavalin (TCan), sword bean (*Canavalia gladiata*) was proteolyzed by *Bacillus subtilis* at 33 °C for 4 days under the condition of dissolved oxygen (20–40% saturation) and pH 7.0. After proteolysis, the product was centrifuged (12,000 × g) at 4 °C for 15 min, and supernatant was collected and lyophilized. The final purification was performed by size exclusion chromatography (SEC) using a WTC-050S5 column (Wyatt Technology, Santa Barbara, CA) that was equilibrated with PBS (140 mM NaCl, 2.7 mM KCl, 6.5 mM $Na_2HPO$, 1.5 mM $KH_2PO$, pH 7.4). Pure protein was frozen in liquid nitrogen and stored at −80 °C. The SEC profile of TCan is shown in Figure S3.

##### On-line top-down analysis of TCan using UPLC-ESI-QTOF (top-down)

On-line top-down MS was carried out on a waters ACQUITY UPLC I-Class system coupled to Synapt G2-S QTOF mass spectrometer (Waters Corp., USA). Briefly, 2 μL of purified TCan was injected into a 2.1 × 50 mm ACQUITY UPLC BEH300 C4 column (particle size 1.7 μm). The gradient was delivered at 400 μL/min, starting at 5% buffer B (0.1% formic acid in acetonitrile), then increased to 95% B at 6.7 min, and 5% B at 9.0 min. The mass spectrometer was operated in the ESI-Positive $MS^E$ continuum data acquisition mode with the following conditions: capillary voltage 3.0 kV, cone voltage 50 V, desolvation gas 800 L/h, source temperature 120 °C, and desolvation temperature 300 °C. Data were collected at 1 spectra/second from 500–4,000 m/z. A maximum entropy algorithm (MaxEnt1) was used for mass deconvolution. Deconvolution was performed with a range of

1,500–2,600 Da around the expected mass, a target resolution of 0.5 Da, with 100 iterations of MaxEnt1. The electrospray spectra of TCan is shown in Supplementary Figure S4.

**Protein analysis using Nano UPLC-QTOF for TCan analysis (bottom-up)**

The purified TCan was digested using the "Tube-Gel digestion" protocol as described previously.[2] LC-MS/MS was conducted according to a previous procedure.[3] The tryptic peptide mixtures were dissolved with 0·5% trifluoroacetic acid prior to further analysis. A 5 μL dissolved sample was added onto a 100 μm × 2 cm nanoviper trap column and 15 cm × 75 μm nanoviper analysis column (Thermo Fisher Scientific) at a flow rate of 300 nL/min and were eluted with a gradient of 5%–40% acetonitrile over 95 min. All MS and MS/MS spectra captured by the Q Exative Plus mass spectrometer (Thermo Fisher Scientific) were acquired in the data-dependent top 12 mode. The MS/MS data were analyzed using MASCOT 2·7, with a parameter corresponding to a false discovery rate (FDR) of 1%. One missed cleavage was allowed for identifying the peptides from TCan.

**N-Terminal sequencing of TCan**

For N-terminal amino acid sequencing, the protein samples were transferred from an SDS–PAGE gel to PVDF membrane and the transferred protein band on the membrane was analyzed using the LC 492 Protein Sequencing System (Applied Biosystems Instruments, USA).

**Preparation of SARS-CoV-2 spike protein**

The gene encoding S2 of the SARS-CoV-2 spike protein (accession number QKU5385.1, amino acid residues 686–1213) was codon-optimized for protein expression in *Nicotiana benthamiana*, synthesized by GenScript Biotechnology Corp. (Nanjing, China) and cloned into a pCAMBIA1300 vector with 8 histidine tag, ER retention signal HDEL, and Cor1 sequence. *N. benthamiana* leaves (fresh weight) expressing recombinant S2 were harvested and homogenized using a blender in the presence of an extraction buffer (25 mM Tris-HCl [pH 8.0], 300 mM NaCl, 10 mM imidazole, 5% glycerol, 1% Sarkosyl, 10 mM $H_2O_2$, and 1 mM PMSF). To remove debris, the extracts were centrifuged at 20,000 g for 40 min, and supernatants were filtered through Miracloth (EMD Millipore Corp., Billerica, MA, USA). The resulting supernatant was loaded onto a 100-mL Ni-IDA agarose column (Bio-Rad, Seoul, Republic of Korea). The column was washed with a washing buffer consisting of 25 mM Tris-HCl (pH 8.0), 300 mM NaCl, and 10 mM imidazole. S2 was eluted with the same buffer containing 30–300 mM imidazole. The proteins were further purified using gel-filtration chromatography (Superdex 200 Increase 10/300 GL, GE Healthcare, USA), pre-equilibrated with phosphate-buffered saline (PBS), and stored at -80 °C.

**Circular dichroism spectroscopy**

Circular dichroism (CD) measurements were performed on a JASCO J-1500 spectrophotometer (JASCO, Japan) using a quartz cuvette with 0·1-cm path length. Samples were prepared using 0.25 mg/mL of TCan in 20 mM sodium phosphate buffer (pH 7·5). Far-UV CD spectra from 195 to 250 nm were obtained at a scanning rate of 200 nm/min. CD signals were expressed as mean residue ellipticity [θ] (degrees $cm^2$/dmol).[4] For the thermal scans, far-UV CD spectra were acquired every 5 °C from 5 to 100 °C at a heating rate of 1 °C $min^{-1}$. Then, monitoring CD signals at 204 nm was carried out by decreasing the temperature from 100 to 5 °C at a rate of 10 °C/ /min. The temperature was controlled using an RW-0525G low-temperature bath circulator (Lab Companion, Republic of Korea).

The melting temperature (${}^{CD}T_m$), enthalpy change (${}^{CD}\Delta H$), and heat capacity change ($\Delta C_p$) for the heat denaturation of folded TCan were obtained by regression analysis as follows:[5]

$$\theta = \frac{(\mathrm{a}-\mathrm{c}) + (\mathrm{b}-\mathrm{d})T}{1 + \exp\left(-\dfrac{{}^{CD}\Delta H({}^{CD}T_m)\left(1 - \dfrac{T}{{}^{CD}T_m}\right) - \Delta C_p\left({}^{CD}T_m - T + T\ln\dfrac{T}{{}^{CD}T_m}\right)}{RT}\right)} + (c + dT)$$

where $\theta$ is the signal intensity monitored by CD. The pre- and post-unfolding baselines are described by a + b$T$ and c + d$T$, respectively. $T$ is the temperature in Kelvin, and $R$ indicates the gas constant. ${}^{CD}T_m$ is the midpoint temperature of the thermal denaturation of TCan, $\Delta C_p$ is the heat capacity change between pre- and post-unfolded TCan, and ${}^{CD}\Delta H$ (${}^{CD}T_m$) is the enthalpy change at ${}^{CD}T_m$. CD spectra were analyzed using the BeStSel algorithm[6] to estimate the content of the secondary structure. The concentration of TCan was determined using the UV absorbance and a molar extinction coefficient of 8,940/M cm at 280 nm.

**Dynamic and light scattering measurement**

Dynamic light scattering (DLS) and static light scattering measurements of TCan in 20 mM sodium phosphate buffer (pH 7.5) were performed using a DynaPro Plate Reader II (Wyatt Technology Corp., USA) and a Cary Eclipse fluorescence spectrophotometer (Agilent Technologies, USA) at 25 °C, respectively. For DLS measurements, sample solutions containing 11·7 μM TCan before and after heat treatment were subjected to centrifugation at 10,000 rpm for 5 min, and the supernatants were loaded into a 384-well plate. The average hydrodynamic radius ($R_h$) of the 20 measurements was obtained. Experimental data were processed and analyzed using the DYNAMICS software (Wyatt Technology Corp., ver. 7.0). The emission spectrum of light scattering of approximately 12 μM TCan was recorded from 300 to 400 nm with an excitation wavelength of 350 nm. The slit widths of the excitation and emission wavelengths were 10 and 5 nm, respectively. Scattering of 20 mM sodium phosphate buffer (pH 7.5) was also performed as a control using the same method.

**Intrinsic and extrinsic fluorescence spectroscopy**

All fluorescence measurements were carried out in the absence and presence of 6.0 M Urea using a Cary Eclipse fluorescence spectrophotometer (Agilent Technologies, USA) and approximately 11.7 μM TCan in 20 mM sodium phosphate buffer (pH 7·5) at 25 °C. Intrinsic fluorescence spectra were obtained by exciting phenylalanine and tyrosine/tryptophan at 240 and 280 nm, respectively. An excitation slit of 10 nm and an emission slit of 20 nm were used. ANS (50 μM) was utilized as an extrinsic fluorophore, and its emission spectrum was recorded from 370 to 600 nm with an excitation wavelength of 370 nm. The concentration of ANS was determined using UV absorbance at 350 nm and a molar extinction coefficient of 4,950/M cm. The emission spectra were monitored from 370 nm to 600 nm. The excitation and emission slit widths for intrinsic and extrinsic fluorescence were both set to 20 nm. Fluorescence measurements of 20 mM sodium phosphate buffer (pH 7·5) were carried out as a control using the same method.

**Differential scanning calorimetry**

Differential scanning calorimetry (DSC) experiments of 17.6 μM TCan in 20 mM sodium phosphate buffer (pH 7·5) were carried out using a PEAQ-DSC instrument (Malvern Panalytical, UK). The TCan solution was heated from 20 °C to 100 °C at a heating rate of 1 °C/min. After subtraction of the buffer baseline and normalization with protein concentrations, fitting analyses were performed based on the non-two-state transition using PEAQ-DSC analysis software (Malvern Panalytical, UK). The fit equation used was based on the Levenberg-Marquardt nonlinear least-squares method.[7]

$$C_p(T) = \frac{K(T)^{\mathrm{DSC}}\Delta H_{\mathrm{VH}}{}^{\mathrm{DSC}}\Delta H_{\mathrm{cal}}}{(1+K(T))^2 RT^2} + \cdots$$

$$K(T) = \exp[\frac{-{}^{\mathrm{DSC}}\Delta H_{\mathrm{VH}}}{RT}\left(1 - \frac{T}{{}^{\mathrm{DSC}}T_{\mathrm{m}}}\right)]$$

where $T_m$ is the thermal midpoint of a transition, as described above. The change in calorimetric enthalpy (${}^{DSC}\Delta H_{cal}$) was determined using the DSC peak area, and the enthalpy change was based on the van't Hoff method (${}^{DSC}\Delta H_{VH}$) and was estimated by exploiting the shape of the DSC peak, that is, the slope of the transition curve between folded and unfolded proteins. $C_p(T)$ was calculated at any temperature $T$ in Kelvin. $K(T)$ indicates the equilibrium constant as a function of the temperature.

**Isothermal titration calorimetry**

Isothermal titration calorimetry (ITC) was performed using a MicroCal PEAQ-ITC (Malvern Panalytical, UK) at 37 °C. All samples, TCan, S1, and S2, were prepared in 20 mM sodium phosphate buffer (pH 7·5) containing 100 mM NaCl. TCan (287.6 μM) in an ITC syringe was titrated to solutions containing either S1 or S2 at 17 μM in an ITC cell with continuous stirring at 750 rpm. The total number of titrations was 19 with 150-s intervals between titration. The initial delay and reference power were set to 60 s and 10 μcal/s, respectively. The dilution heat of TCan was also measured using the same method and the ITC parameters. Binding isotherms after the subtraction of dilution heat and the baseline correction were fitted to a one-set of sites binding model:[8]

$$Q = \frac{\mathrm{n}[\mathrm{S\ protein}]_\mathrm{t}{}^{\mathrm{ITC}}\Delta H_\mathrm{bind}V_0}{2}[1 + \frac{[\mathrm{TCan}]_\mathrm{t}}{\mathrm{n}[\mathrm{S\ protein}]_\mathrm{t}} + \frac{K_\mathrm{d}}{n[\mathrm{S\ protein}]_\mathrm{t}} - \sqrt{\left(1 + \frac{[\mathrm{TCan}]_\mathrm{t}}{\mathrm{n}[\mathrm{S\ protein}]_\mathrm{t}} + \frac{K_\mathrm{d}}{\mathrm{n}K[\mathrm{S\ protein}]_\mathrm{t}}\right)^2 - \frac{4[\mathrm{TCan}]_\mathrm{t}}{\mathrm{n}[\mathrm{S\ protein}]_\mathrm{t}}}]$$

where $Q$ is the total heat constant, and n indicates the number of TCan molecules which bind to one of the S proteins (*i.e.*, S1 and S2). ${}^{ITC}\Delta H_{bind}$ is the change in enthalpy for the intermolecular interactions. $V_0$ is an active cell volume which considers the volume of the ITC cell and the volume increased by titration. The total concentrations of S proteins and TCan are displayed as $[\mathrm{S\ protein}]_t$ and $[\mathrm{TCan}]_t$, respectively, at any given time ($t$). The dissociation constant is denoted as $K_d$. In binding isotherms of TCan-S1 and TCan-S2, a best fit was obtained without fixation of variable, and fit by fixing a value of $K_d$ was also performed.

**Size exclusion chromatography-combined multi-angle light scattering measurement**

Size exclusion chromatography with inline multi-angle light scattering (SEC-MALS) measurements were conducted using an instrument from Wyatt Technology (USA). Freshly purified 94·4 µM TCan solution was loaded onto a WTC-050S5 column (Wyatt Technology Corp., USA) with PBS running buffer. The flow rate was set to 0·5 mL/min. MALS and UV spectra were recorded using a DAWN HELEOS-II (Wyatt Technology Corp., USA), and Optilab T-rEX (Wyatt Technology Corp., USA) was used to determine the differential refractive index. SEC-MALS results were analyzed using Astra 6·1 software (Wyatt Technologies Corp., USA). A differential refractive index increment (dn/dc) value of 0.185 was used.

**Cell culture, viruses, and treatment**

To compare the cytotoxicity of BE on different cell types, two human hepatocellular carcinoma cell lines (HepG2 and Huh7 cells) and two lung origin cell lines (adenocarcinomic human alveolar basal epithelial cells, A549, and alveolar macrophages, MH-S), were purchased from the American Type Culture Collection (ATCC, Manassas, VA, USA). A549 cells were cultured in RPMI 1640 containing 5% FBS and 1% streptomycin/penicillin at 37 °C in an atmosphere containing 5% $CO_2$. MH-S cells were cultured in RPMI 1640 containing 10% FBS and 1% streptomycin/penicillin at 37 °C in an atmosphere containing 5% $CO_2$. HepG2 and Huh7 cells were cultured in DMEM containing 10% FBS and 1% streptomycin/penicillin at 37 °C in an atmosphere containing 5% $CO_2$. Using sterile 24-well culture plates, cells were seeded at a concentration of $2 \times 10^5$ in 500 µL of each medium and treated with Con A or BE at 5, 10, 25, 50, 100, 200, and 400 ppm.

**Determination of cell viability**

After 6 h of BE treatment, A549, HepG2, MH-S, and Huh7 cells were harvested using 0.25% trypsin-EDTA and counted using a NucleoCounter (NC-250, ChemoMetec, Gydevang, Denmark), according to the manufacturer's protocol. Cell viability was calculated as the ratio of live cells to total cells. Inhibitory concentration (IC) was also calculated from nonlinear regression of concentration-response inhibition curves using GraphPad Prism v·7 (GraphPad Software, San Diego, CA).

**Measurement of reactive oxygen species (ROS) levels**

A549, HepG2, MH-S, and Huh7 cells were seeded and treated with BE, as previously described. The treated cells were incubated for 30 min at 37 °C in culture medium containing 3.3 µmol/L DCF-DA for ROS detection, and the cells were washed with PBS. DCF-DA intensity in the cells was immediately measured using flow cytometry (CytoFLEX, Beckman Coulter, Brea, CA, USA). ROS production in the cells is represented as a percentage of DCF-DA intensity relative to the naïve control, which was defined as 100%.

**Primary splenocyte preparation and culture**

Primary splenocytes were isolated from BALB/c and C57BL/6 mice. Single cell suspensions were obtained by mincing the spleen and gently pressing the fragments through a 45 µm nylon cell strainer (BD Falcon, Bedford, MA, USA). The suspension was mixed with 1× RBC lysis buffer (eBioscience Inc., San Diego, CA, USA) for 5 min at room temperature. For primary culture, the spleen cells were resuspended in RPMI 1640 containing 5% FBS, 100 U/mL penicillin, and 100 µg/mL streptomycin. Using sterile 24-well culture plates, cells were seeded at a density of $3 \times 10^6$ cells in 500 µL of medium. Spleen cells were cultured for 24 or 48 h in the presence of 2.5 µg/mL Con A in a humidified atmosphere of 5% $CO_2$ and 95% air. Cellular morphology was observed under a light microscope (20×; Leica Microsystems, Wetzlar, Germany). Cells were harvested and centrifuged at 2,000 ×

*g* for 5 min; thereafter, the supernatants were collected and stored at −70 °C until measurement of inflammatory cytokine levels by ELISA.

**Animals**

Male C57BL/6 mice (Orient Bio Ltd, Seongnam, Korea) were housed under 12 h light/dark cycles in temperature-controlled (22 ± 30 °C) rooms. Throughout the experiments, they had *ad libitum* access to standard laboratory chow and tap water. The experiments were performed in accordance with the protocols approved by the Institutional Animal Care and Use Committee of the Korea Institute of Toxicology (No. 2001–0007, 2005–0163, and 2008–0244).

**Experimental design for *in vivo* study**

The following three studies were conducted to compare the toxic effects of BE:
Study 1) Effect of high-dose BE acute toxicity on survival rate
Study 2) Comparison of BE hepatotoxicity
Study 3) Comparison of BE respiratory toxicity to determine the possibility of intratracheal instillation as a mode of administration to measure the antiviral efficacy of BE

Study 1: To examine and compare the effect of BE on survival rate, seven groups were created (n=3 per group), *i.e.*, vehicle control group, Con A treatment groups (40, 80, and 160 mg/kg), and BE treatment groups (75, 150, and 200 mg/kg). BE was diluted in saline to create equivalent doses, which were intratracheally administered using a modified automatic video instillator (Doobae System, Seoul, South Korea). Following Con A and BE administration, the survival rate was monitored for 24 h.
Study 2: To examine and compare the hepatotoxicity of Con A and BE, mice were administered Con A and BE intravenously (5 and 15 mg/kg) and intratracheally (10, 20, and 40 mg/kg). Several parameters were then evaluated, including serum alanine aminotransferase (ALT), aspartate aminotransferase (AST), total bilirubin (TBIL), and γ-glutamyl transferase (GGT) levels in the blood, as well as histological changes in the liver.
Study 3: To examine and compare the toxicity of Con A and BE on the lungs, mice were intratracheally administered different concentrations of Con A or BE, *i.e.*, 1, 2.5, 5, and 10 mg/kg. Organ weight, cellular changes, and protein levels of inflammatory cytokines in bronchoalveolar lavage fluid (BALF), as well as histological changes in the lung and spleen tissues were assessed 24 h after the single administration of Con A or BE.

**Intratracheal and intravenous instillation**

For intratracheal instillation, mice were anesthetized by inhalation. Prior to instillation, isoflurane was delivered into the induction chamber using a small animal porTable anesthesia system (L-PAS-02, LMSKOREA, Inc., Seongnam, South Korea) equipped with an isoflurane vaporizer. The mice were then exposed to 2·5% isoflurane delivered in $O_2$ (2 L/min) within the induction chamber until a sleep-like state was reached. Mice receiving isoflurane anesthesia were removed from the induction chamber, and instillation was performed immediately. For intravenous instillation, mice were placed in a murine restraining device. The tail vein was dilated by soaking the tail in warm (e.g., at 30–35 °C) water for 1 to 2 min. After swabbing with 70% alcohol, the tail veins were injected with the designated concentration. Mice were subsequently transferred to their cages.

**Bronchoalveolar lavage fluid preparation**

Twenty-four hours after Con A and BE instillation, mice were anesthetized and the left lung was ligated, while the right lungs were gently lavaged three times via a tracheal tube with a total volume of 0.7 mL PBS. The total number of cells in the collected bronchoalveolar lavage fluid (BALF) was counted using a NucleoCounter. For differential cell counts, BALF cell smears were prepared using Cytospin (Thermo Fisher Scientific) and stained with the Diff-Quik solution (Dade Diagnostics, Aguada, Puerto Rico), followed by counting of different cell types (n = 200/slide). BALF was immediately centrifuged at 2,000 × *g* for 5 min, and the collected supernatant was stored at −70 °C until measurement of cytokine levels by ELISA.

**Measurement of cytokine levels**

Interleukin (IL)-4, IL-5, IL-13, IL-17, IL-1β, IFN-γ, and tumor necrosis factor alpha (TNF-α) levels in BALF and cell supernatants were quantified by ELISA using commercial kits (Thermo Fisher Scientific), according to the manufacturer's protocols. The sensitivity for IL-4, IL-5, IL-13, IL-17, IL-1β, IFN-γ, and TNF-α assays were 0.32, 3.3, 2.8, 1.2, 5.3, and 3.7 pg/mL, respectively. The intra and inter assay coefficients of variation for IL-4, IL-5, IL-13, IL-17, IL-1β, IFN-γ and TNF-α were 7.1% 7.9%, 5%, 4.7%, <5.0% and 6.5%, and 10%, 4.6%, 5%, 5.7%,

< 10% and 5.7%, respectively.

**Histological analysis**

Twenty-four hours after BE instillation, the mice were euthanized for histological analysis. Lung and liver tissues were removed and fixed in 10% (v/v) neutral-buffered formalin, dehydrated, embedded in paraffin, and cut into 4 µm sections. They were then deparaffinized with xylene and stained with hematoxylin and eosin (Sigma-Aldrich). Stained sections were analyzed using a light microscope (Axio Imager M1; Carl Zeiss, Oberkochen, Germany). The degree of inflammation was scored on a scale of 0–4.

#### Supplementary Study Results and Discussion

##### Characterization of TCan

In this work, fermented sword bean (*Canavalia gladiata*) was used and the active ingredient TCan was successfully produced in culture medium during the cultivation. Interestingly, N-terminal sequence of the canavalin was determined by Edman degradation as $^{242}$LSSQDKPFN$^{251}$, indicating that truncation occurs at N-terminal regions of canavalin of *C. gladiata*. To know the mass value of the whole protein, the multiple-charge electrospray mass spectrum (Figure S4) of TCan protein was deconvoluted to calculate protein molecular weight by MaxEnt1 software. The mass value of the TCan protein was observed to be 21,176 Da, which was predicted to be truncation of not only the N-terminal but also the C-terminal part, lysin (190). The peptides of the protein were identified using Q Exative Plus mass spectrometer. A protein identification search revealed that the peptides present in the predicted TCan matched with those of canavalin (*C. gladiata*, P10562·1), providing 97% sequence coverage for the predicted TCan amino acid sequence using LC-MS/MS bottom-up analysis. In future, it is necessary to verify the biochemical characterization of TCan by designing a recombinant protein.

##### Structural characterization of TCan

To understand the molecular properties of TCan, we first performed a structural investigation using circular dichroism (CD) (Figure S5A) and fluorescence spectroscopic techniques (Figure S5 B to D). Far-UV CD spectroscopy has been extensively used to obtain information on the secondary structures of proteins based on the intrinsic pattern of a spectrum.[9] Intrinsic and extrinsic fluorescence have been shown to be capable of characterizing tertiary structures around fluorophores based on the wavelength at the maximum intensity of fluorescence emission and emission intensity.[10]

Far-UV spectra at 25 and 37 °C showed a similar pattern, with a minimum at ~215 nm and a maximum at ~200 nm, suggesting a mixture of helical and β structures in a folded state at both temperatures (Figure S5A and S7A). Further analyses of the secondary structures indicated that TCan consists of several types of secondary structures, and β structures show a higher proportion than helical structures by ~20% (Figure S5D). These results are in line with the three-dimensional (3D) structure of TCan determined by X-ray crystallography (Figure S6E and F):[9,11] TCan is mainly composed of two antiparallel β-sheets containing 13 β-strands (β1–β13) and four helices (α1–α4) (Figure S7A and D).

Next, three-dimensionally folded structures of TCan were examined using 1-anilino-8-naphthalenesulfonate (ANS) as an extrinsic fluorophore and phenylalanine (Phe) and tyrosine (Tyr) as intrinsic fluorophores.[10] ANS is a well-known probe for clarifying partially unfolded structures such as a molten-globule structure, which has the same secondary structure as a native structure without a solid 3D conformation. Thus, binding of ANS to a partially unfolded structure increases its fluorescence emission, that is, a red shift. Although a subtle change was observed, ANS spectra of TCan in the absence and presence of 6 M urea, a chemical denaturant, were similar to the spectrum of the buffer, indicating that ANS did not bind to TCan in either conditions and that TCan has a 3D structure (Figure S6B). The Phe fluorescence spectrum in the absence of urea showed a weak intensity at ~310 nm (Figure S6C), suggesting that the six Phes were not exposed to polar solvents and buried in cores, as indicated in the 3D structure (Figure S6E). The similar spectra at 0 and 6 M urea implied that physicochemical environments around Phes are not remarkably different in the folded and urea-induced unfolded states. Similarly, the Tyr fluorescence spectrum without urea exhibited a weak maximum emission at ~310 nm (Figure S6D)—similar to that with 6 M urea—demonstrating that five Tyrs are buried in the cores (Figure S6F) and are located in a similar environment, regardless of urea.

To explore the internal dynamic structures and core regions, we examined X-ray *B*-factors and the hydrophobicity of each residue in TCan (Figure S7). Higher values of the *B*-factor have been interpreted as higher flexibility of atoms and/or residues, and residues with higher hydrophobicity are often buried in the core regions of folded proteins.[9,12] Most *B*-factors ranged between 40 and 60 Å$^2$, and β-structures showed a tendency of lower *B*-factors than those of helical structures (Figure S7A). Random coil-like structures, including both terminal parts and loops between structured secondary structures, exhibited high *B*-factors. The loop region between the two β-sheets also showed a high number of *B*-factors. In contrast, N- ($\sim\beta_1$-$\sim\alpha_1$) and C- terminal parts ($\sim\beta_{12}$-$\sim\beta_{13}$) and the loop region between the two β-sheets showed low hydropathy scores, whereas the two central regions, $\sim\beta_4$-$\sim\beta_7$ and $\sim\beta_8$-$\sim\beta_{11}$, showed high hydropathy scores (Figure S7B and D). Interestingly, these two regions individually consisted of a β-sheet and displayed low *B*-factors, revealing a hydrophobic core of TCan (Figure S7D).

Considering these results together, we can conclude that TCan, a truncated region of canavalin, has a secondary and tertiary structure similar to the native structure of the globular protein. Hydrophobic cores of the two β-sheets with low flexibility are capped by two helical regions ($\sim\alpha_1$ in the N-terminal part and $\sim\alpha_2$ $\sim\alpha_4$ in the C-terminal part) and most likely contribute to holding a stereo-structure.

**Characterization of conformational stability of TCan**

To utilize TCan as a biomaterial and diagnostic agent, it is highly important to confirm conformational stability. Therefore, we examined the thermal stability of TCan using spectroscopy and calorimetry. Far-UV CD spectra did not change significantly in the temperature range between 10 and ~70 °C; however, at temperatures over ~70 °C, there were marked changes in the spectra (Figure S5A). The disappearance of a positive CD signal at ~200 nm and a simultaneous increase in the magnitude of the negative CD intensity at ~205 nm suggested the disruption of secondary structures, which is often accompanied by global or sub-global unfolding of 3D structures. Tracing of the CD intensity at 204 nm with an increase in temperature manifested a clear transition of heat-induced unfolding of TCan (Figure S5B). The CD intensity decreased from ~70 °C due to thermal denaturation and was saturated at ~80 °C, which indicated that all TCan thermally unfolded. A sharp transition was detected, indicating cooperative unfolding due to the high packing density interior of TCan, which comes from its high hydrophobicity and low flexibility. This revealed that TCan has a 3D structure as a sTable globular protein.

In accordance with the CD results, differential scanning calorimetry (DSC) displayed a positive DSC peak in the thermogram, revealing thermal unfolding of TCan by uptaking heat for denaturation (Figure S6C). An endothermic DSC peak appeared from ~70 °C and was at the level of the baseline from ~70 °C. Temperature-dependent changes in the secondary structure suggested that decreases in the content of β-structures and increases in random coil conformations were followed by thermal unfolding of TCan (Figure S5D). We believe that the hydrophobic cores of β-sheets essential for structural stability were disrupted due to heat stress.

With the assumption of a reversible reaction, we further carried out quantitative analyses of the conformational stability of TCan based on two-state unfolding. The melting temperature ($T_m$), at which unfolded and folded populations of TCan were identical, and the change in enthalpy ($\Delta H$) for unfolding were obtained using CD (${}^{CD}\Delta H$) and DSC (${}^{DSC}\Delta H$) (Tables S5 and S6). Tm values calculated from CD (${}^{CD}T_m$) and DSC (${}^{DSC}T_m$) were ~75 and ~76 °C, respectively, revealing a relatively high $T_m$ as a number of proteins showed $T_m$ lower than 70 °C.[13] CD analyses with the best fit also produced an endothermic ${}^{CD}\Delta H$ of ~100 kcal/mol and a change in heat capacity ($\Delta C_p$) of ~2 kcal/mol (Table S5), which falls into the range of values of globular proteins.[14] Hydration of the hydrophobic cores would cause a positive $\Delta C_p$ value. $\Delta H$ obtained using the van't Hoff method and DSC thermogram (${}^{DSC}\Delta H_{VH}$) was ~160 kcal/mol (Table S6), which was higher than that from CD; however, it was still in the range of $\Delta H$ for globular proteins.

The reversibility of the thermal unfolding of TCan was tested using several approaches (Figure S8). The far-UV CD spectrum after heat scanning was not superimposed, and a spectrum indicating an unfolded structure remained (Figure S8A). The ANS spectrum of heat-treated TCan manifested an increased fluorescence emission with a red shift, indicative of the exposure of hydrophobic regions of TCan with partial unfolding (Figure S8B). The intrinsic fluorescence spectra of Phe (Figure S8C) and Tyr (Figure S8D) of TCan subjected to heat treatment did not exhibit appreciable changes. The distribution of the hydrodynamic radius ($R_h$) of TCan after heat incubation was wider than that before heat scanning (Figure S8E). The average Rh was 3.76 nm; however, new molecular species with higher and lower molecular weights appeared to form. A decrease in the light scattering intensity also signified different structural states of TCan depending on the thermal scan (Figure S8F). These results indicate that thermally unfolded TCan cannot refold to its original structure, that is, irreversible unfolding of TCan.

In summary, a systematic investigation revealed the conformational stability of TCan with detailed thermodynamic properties. Although thermal unfolding of TCan is irreversible, it has been demonstrated that TCan with well-packed hydrophobic cores with low flexibility is a fully sTable protein for biological and diagnostic applications.

**Investigation on oligomeric states of TCan**

The association states of proteins are important for understanding the underlying mechanism of the stability and intermolecular recognition of proteins for functional, biological and diagnostic applications. In this regard, we focused on the colloidal and association states of TCan. Light scattering has been used to examine the association and colloidal states of proteins. The scattering intensity of TCan is larger than that of the buffer (Figure S5E), suggesting that TCan may exist as a multimer. Dynamic light scattering (DLS) revealed a monodisperse $R_h$ with an average Rh of 3.95 nm (Figure S5F). Based on the relationship between the number of residues, protein volume, and Rh,[15] it was calculated that approximately six TCan monomers consist of oligomers. Size exclusion chromatography with inline multi-angle light scattering (SEC-MALS) also elucidated the oligomeric states of TCan (Figure S5G). Considering that the molecular weight of the TCan monomer is 21,100, the molecular weight of ~140,000 obtained using SEC-MALS indicated that approximately seven TCan monomers interact

intermolecularly to form an oligomer. Furthermore, consistent with DLS and MALS analyses, DSC also showed the oligomeric states of TCan (Table S6). We used the fact that the ratio of ${}^{DSC}\Delta H_{VH}$ / ${}^{DSC}\Delta H_{cal}$ can be a criterion for the oligomeric state of proteins.[16] As the ratio of ${}^{DSC}\Delta H_{VH}$ (158.0 ± 0.7) / ${}^{DSC}\Delta H_{cal}$ (19.3 ± 0.1) was 8.3 ± 0.1, it was conceived that a TCan oligomer accommodates approximately eight monomers.
Taken together, TCan forms an oligomer containing ~6 to ~8 monomers under physiological conditions. We rationalized that TCan efficiently uses spontaneous oligomerization by escaping misfolding and aggregation which inactivates TCan. Furthermore, because intermolecular associations such as dimerization and a higher order of oligomerization often strengthen conformational stability,[17] oligomer formation contributes to the relatively high stability of TCan against outer stresses in addition to internal stabilizing factors, as mentioned above. Oligomerization of TCan may also be beneficial to protect itself from external perturbations, such as proteolysis, and be functional.

**Intermolecular interactions between TCan and spike proteins**
The binding capability of TCan for the two SARS-CoV-2 spike proteins, S1 and S2, was investigated using isothermal titration calorimetry (ITC). ITC is one of the most powerful and accurate methodologies for studying intermolecular interactions without labeling or immobilization.[18–20] It can elegantly clarify both weak and strong intermolecular interactions by detecting even a small amount of reaction heat.
Titration of TCan to either S1 (Figure 2I, left panel) or S2 (Figure 2I, right panel) produced energetically favorable exothermic heat, revealing interprotein interactions between TCan and spike proteins. The reaction heat of TCan-S1 was greater than that of TCan-S2, and, thus, the magnitude of the negative enthalpy change for binding (${}^{ITC}\Delta H_{bind}$) of TCan-S1 (~-6—~-7 kcal/mol) seemed to be larger than the ${}^{ITC}\Delta H_{bind}$ of TCan-S2 (~-0·8 kcal/mol). The two binding systems did not show saturation at a molar ratio of 3, indicating that intermolecular interactions are not strong. Although binding isotherms were not saturated, we attempted to evaluate the apparent dissociation constant ($K_d$). The apparent $K_d$ values for the TCan-S1 and TCan-S2 interactions were in the range of ~20—~100 μM and ~10—~80 μM, respectively. These findings suggest that TCan moderately binds to S1 and S2 with similar affinity. Given the similar interprotein affinity, TCan may bind and recognize a spike protein in a different thermodynamic way: the TCan-S1 interaction is more enthalpically driven, whereas entropy is a predominant player in the TCan-S2 interaction. It should be noted that ITC results, in addition to information on oligomeric states (Figure 2I and Table S6), collaborate with the Cryo-EM and TEM image (Figure 2A–E and S3) which demonstrated binding of several TCan, that is, TCan oligomers, to the surfaces of HCoV-229E and SARS-CoV-2 through S1 and S2.

**Mitogenic response induced by Con A and BE in mice splenocytes**
Con A is well-known for inducing the mitogenic response in splenocytes that activate the immune system, recruit lymphocytes, and elicit cytokine production.[21] To compare the mitogenic effects induced by Con A and BE, we investigated T helper (Th) cytokine (IFN-γ for Th1, IL-17 for Th17, and IL-4, IL-5, and IL-13 for Th2 cells) production by treating splenocytes with Con A or BE at a concentration of 2.5 μg/mL (Figure S9). The results showed that compared to BE, Con A induced significantly higher production of all Th cytokines. Although production of IFN-γ, IL-17, IL-5 and IL-13 was increased in BE-treated splenocytes over the duration of the experiment, the production was significantly lower than that in the Con A-treated group.

**Effects of Con A and BE on survival rate of mice**
To examine and compare the toxic effects of Con A and BE in a living system, we primarily investigated the effects of Con A and BE on mortality after intratracheal instillation in C57BL/6 mice (Figure S10). The survival rate was monitored every hour in C57BL/6 mice divided into seven groups: vehicle control (VC), Con A-treated at the concentration of 40, 80 and 160 mg/kg body weight (BW) (Con A 40, Con A 80, Con A 160 groups), and BE-treated at the concentration of 75, 160 and 200 mg/kg BW (BE 75, BE 160, BE 200 groups) for 24 h. The VC and BE-treated groups exhibited a survival rate of 100%, while the Con A-treated groups exhibited a decrease in survival. The survival rate for the mice in the Con A-treated groups began to decrease from the 17$^{th}$ h of treatment, and no survival of mice in the Con A 160 group was observed by the 24$^{th}$ h of treatment. Meanwhile, 60% survival was observed in mice in the Con A 40 and Con A 80 groups after 24 h of treatment. These results indicate that BE was not toxic to mice, which was consistent with the *in vitro* cytotoxicity results.

**Effects of Con A and BE on liver toxicity of mice**

Con A can activate T cells to secrete cytokines that cause liver injury.[22] To compare the effects of Con A and BE on liver injury, the levels of serum AST, ALT, TBIL, and GGT, which are the biomarkers of liver function, in mice were evaluated at 24 h after Con A and BE administration (Figure S11). The result showed that the mode of administration impacted hepatic biomarker levels. Although, administration by intratracheal instillation of both Con A and BE-treated mice did not significantly affect the level of hepatic biomarkers, the high concentration of Con A (15 mg/kg BW) administered intravenously remarkably increased the expression of ($p$ value < .05) all biomarkers.

**Effects of Con A and BE on lung injury of mice**

Animal experiments were conducted to compare lung toxicity by direct respiratory exposure to Con A and BE. The following parameters were analyzed to assess lung injury: (i) lung and spleen organ weight; (ii) histological analysis of lung and spleen tissue; and (iii) differential cell counts and cytokine expression in the BALF.

Increasing concentrations of intratracheal Con A instillation gradually increased % lung weight per BW compared with the VC. However, BE did not impact % lung weight compared to VC. In fact, increased lung weight indicates a significant increase in pulmonary vascular permeability and inflammatory cell infiltration into damaged lung regions (Figure S12). This result may be related to the increase in various types of inflammatory cells, including macrophages, neutrophils, and lymphocytes, in the BALF of ConA-instilled groups. In contrast to the lung weight, neither Con A nor BE altered the spleen weight up to doses of 5 mg/kg BW. A slight decrease in the spleen weight of mice was observed at the highest level (10 mg/kg BW) of Con A instillation. Spleen is a source of inflammatory cells that rapidly accumulate at sites of tissue injury. Decreased spleen weight indicates that inflammatory cells in the spleen are rapidly deployed to sites of lung injury.

Additionally, changes in the abundance of different inflammatory cell types were assessed in the BALF after Con A or BE instillation in mice to determine the pulmonary inflammatory response. The results revealed a significant increase in the total number of cells, as well as in the counts of macrophages, eosinophils, and neutrophils in the BALF of Con A-instilled mice as compared to the respective VC group (Figure S13). However, an increase in cell numbers was not observed with increasing concentration of Con A (1–10 mg/kg BW) instillation. In contrast, the abundance of different cell types did not differ in BE-instilled mice compared to the VC. Moreover, lymphocytes were not observed in the BALF of Con A- or BE-instilled mice. Meanwhile, the spleen weight was also not increased by Con A or BE instillation at the tested concentration levels. An increase in spleen weight could indicate the mitotic transformation of lymphocytes from small to large cells to counteract lectin administration, as previously observed in case of plant lectin toxicity.[23] In contrast, lung weight was increased in Con A-instilled mice, which may have been caused in response to water accumulation in the lungs after activated pulmonary neutrophil-induced pulmonary permeability. Alternatively, BE may control neutrophil activation in the lung and reduce ROS production, thereby, reducing pulmonary damage.[24]

The pathological changes in organ and inflammatory cells induced in response to lectins resulted in increased expression of inflammatory cytokines (Figure S14). Proinflammatory Th1, Th2, and Th17 cytokines play important roles in triggering lung inflammatory responses.[25–27] Therefore, different cytokines, including IL-1β, IFNγ, and TNFα secreted by Th1, IL-4, IL-5 and IL-13 secreted by Th2, and IL-17 secreted by Th17 cells were measured in the BALF of Con A- or BE-instilled mice. Proinflammatory cytokine levels in BALF were highly increased in the Con A-instilled group (compared to the VC). Th2 cytokine (IL-4, IL-5, and IL-13) levels in the BALF were significantly increased in the Con A-instilled group (compared to the VC); however, the levels of these cytokines appeared to decrease in the Con A-instilled group in a dose-dependent manner. These results are similar to those observed for cellular changes in BAL cells and are more likely to be related to a decrease in eosinophils. However, no change was observed in cytokine levels within the BALF of the BE-instilled group. Consistently, Con A treatment has been reported to induce proinflammatory cytokine expression in human peripheral blood mononuclear cells.[28] Meanwhile, griffithsin, which exhibits effects similar to those of BE, does not induce the production of such cytokines.[28] Moreover, a range of cytokines were expressed following Con A treatment with the highest expression being of IFNγ followed by that of IL-17, IL-5, IL-13, TNF-a, IL-4, and IL-1β. A previous study showed that IFNγ can induce ROS generation and ferroptosis in tumor cells.[29] Thus, Con A treatment may induce ferroptosis by upregulating IFNγ. Moreover, cytokine storms reportedly induce the dysfunction of multiple organs, including the liver, and ultimately can cause death.[30] Therefore, high expression of proinflammatory cytokines and pathological changes within organs, such as the liver and lung, following Con A treatment could explain the high mortality observed in these mice. The results further demonstrated that Con A can mimic the conditions observed in autoimmune hepatitis by inducing acute liver injury as indicated by an increase in the levels of hepatic biomarkers and several proinflammatory cytokines.[31] Thus, unlike the regular modes of programmed cell death, such as apoptosis and pyroptosis, the observation of cytotoxicity, ROS generation and inflammatory cytokine production, may indicate the occurrence of ferroptosis in Con A-treated cells.[32] Indeed, further analysis is required to verify the ferroptosis-related pathological changes in response to Con A treatment.

**Effect of Con A and BE on cytotoxicity and ROS generation**

Relative cell viability was gradually reduced with increasing concentrations of Con A or BE in all cell lines in a dose-dependent manner (Figure S15). However, BE induced a lower rate of cytotoxicity. Concordantly, previous studies have shown that Con A is highly cytotoxic, while griffithsin exhibits low cytotoxicity, comparable to that of BE.[28] The $IC_{20}$ of Con A in HepG2, MH-S, A549, and Huh7 cells treated for 6 h was determined to be 22.44, 35.95, 38.05, and 50.15 μg/mL, respectively. The $IC_{20}$ of BE for HepG2, MH-S, A549, and Huh7 cells treated for 6 h was determined to be 86·95, 167.69, 110.04, and 232.01 μg/mL, respectively. The higher $IC_{20}$ of BE indicated the lower cytotoxicity of BE (compared to Con A). In addition, different cells lines exhibited different sensitivities to Con A and BE, which may be attributed to the differences in protein expression during exposure to Con A.[33] Similar to the cytotoxic effect of Con A, a lower tolerance of HepG2 to the cytotoxic potency of doxorubicin has also been observed previously.[33]

Reactive oxygen species (ROS) levels in cells in response to Con A and BE treatment were measured using DCFH-DA staining.[34] As observed for the cytotoxic effects, ROS production was observed in a concentration-dependent manner in all cell types, including A549, MH-S, HepG2, and Huh7 cells (Figure S16). A comparably high and low level of ROS was observed in A549 and Huh7 cells, respectively, in response to Con A treatment. Similarly, a previous study reported that Huh7 cells have a high tolerance to hypoxic conditions.[33] Consistent with the cytotoxicity assay results, BE treatment resulted in remarkably lower ROS production than Con A.

#### Supplementary Figures

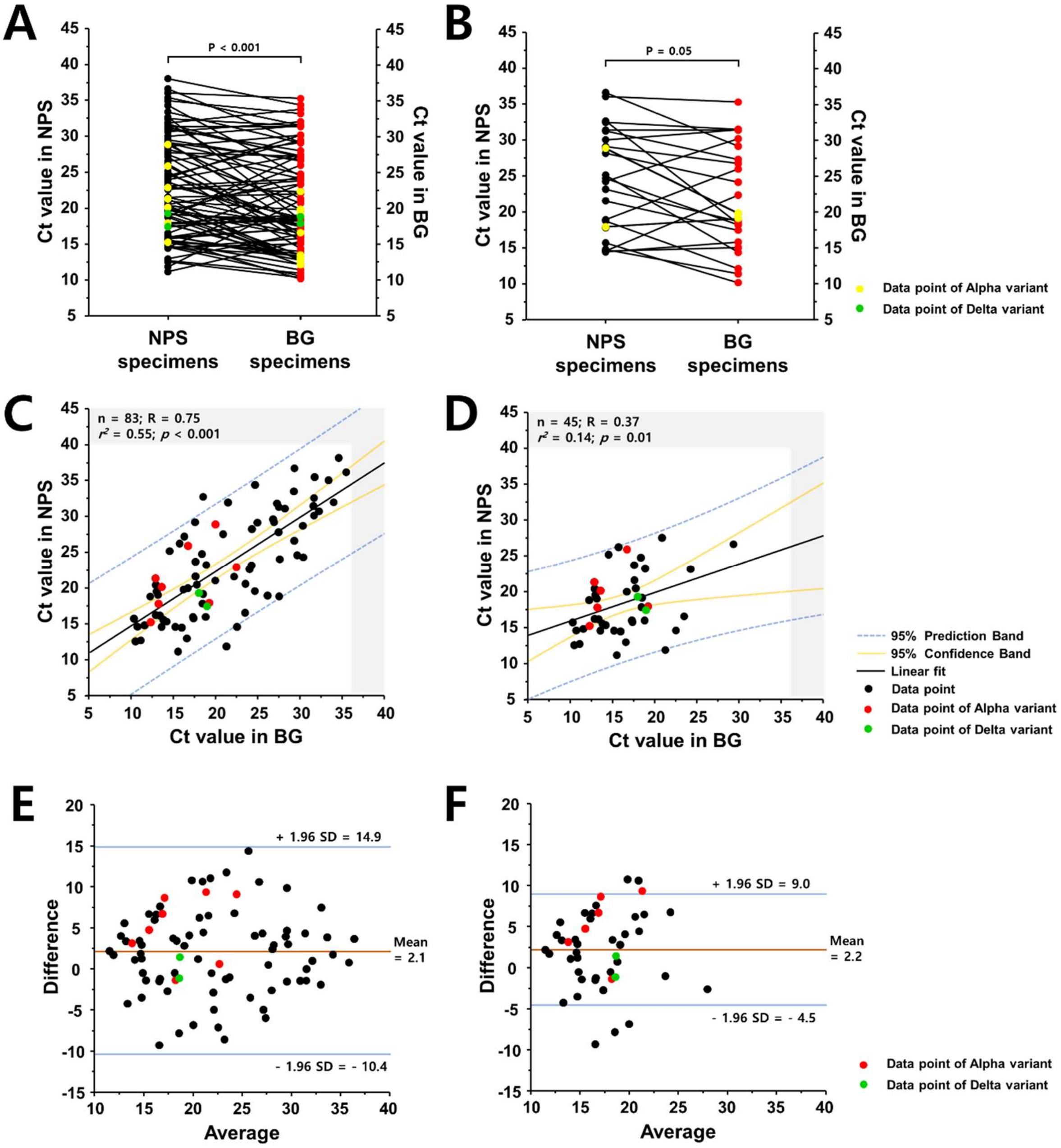


**eFigure 1**. **Comparison of the Ct values for BG-RT-PCR with those for NPS-RT-PCR.** In the case of all patients (A), the mean and median Ct values, which ranged from 10.1 to 35.3, for Beanguard gargle$^{TM}$ (BG) specimens were 20.8 (95% CI, 19.3–22.3) and 18.8 [IQR, 15.5 to 26.9], respectively, whereas the mean and median Ct values for nasopharyngeal swab (NPS) specimens, which ranged from 11.1 to 39.3, were 22.9 (95% CI, 21.4–24.5) and 21.5 (IQR, 17.0–29.0), respectively. (B) The mean and median Ct values for BG specimens for asymptomatic cases were 22.0 (95% CI, 18.8–25.2) and 19.8 [IQR, 16.6–28.3], respectively, whereas Ct values for NPS specimens were 24.6 (95% CI, 21.4–27.8) and 24.7 [IQR, 18.3–30.7], respectively. Correlation of the Ct values for BG-RT-PCR and NPS-RT-PCR with the RdRP gene over the entire period (C) and within 6 days of symptom onset or initial confirmation of COVID-19 (D). Bland-Altman plots for the relationship of Ct values between BG-RT-PCR and NPS-RT-PCR over the entire period (E) and within 6 days of infection (F).

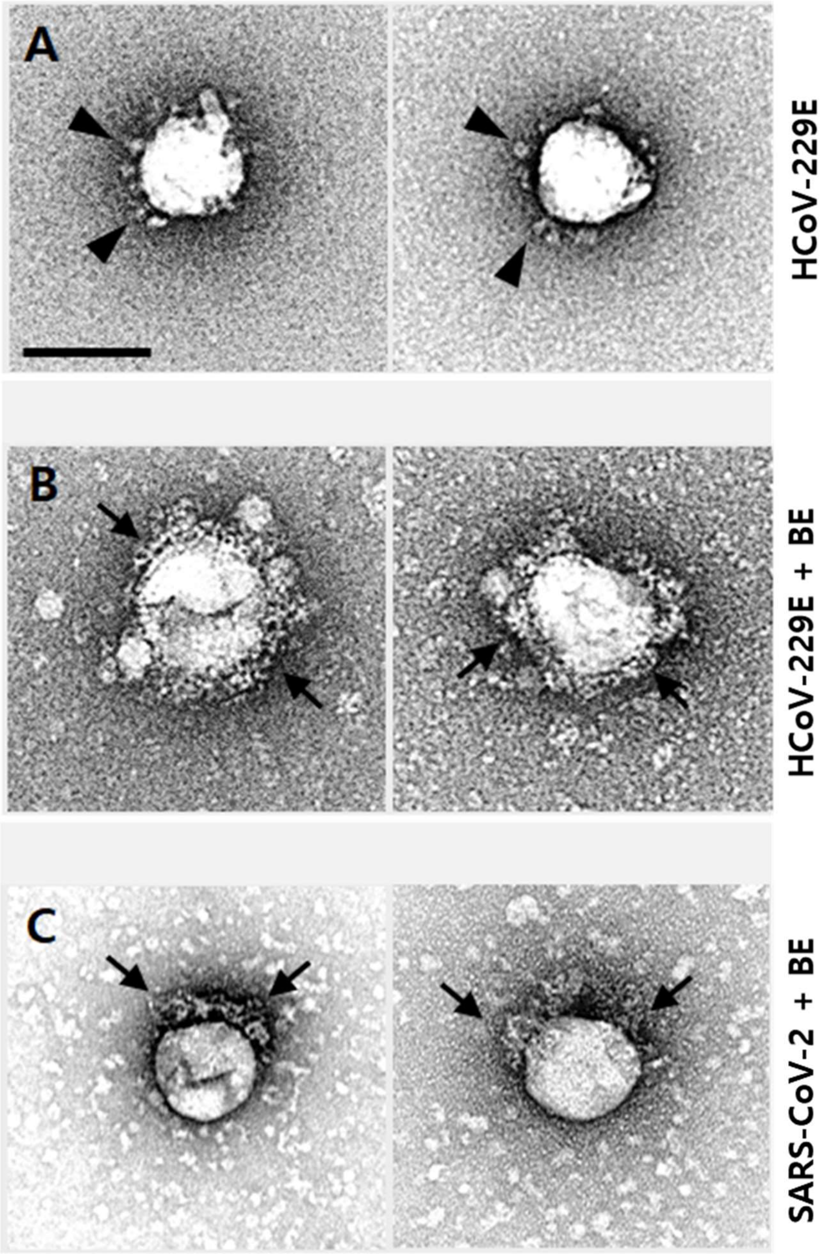


**eFigure 2. Transmission electron micrographs (TEM) of HCoV-229E and SARS-CoV-2 with bean extract (BE) treatment.** TEM images of (A) HCoV-229E before BE treatment, and (B) after treatment and (C) SARS-CoV-2 with BE treatment were obtained. The arrowheads and arrows indicate spikes and BE, respectively. Scale bar = 100 nm.

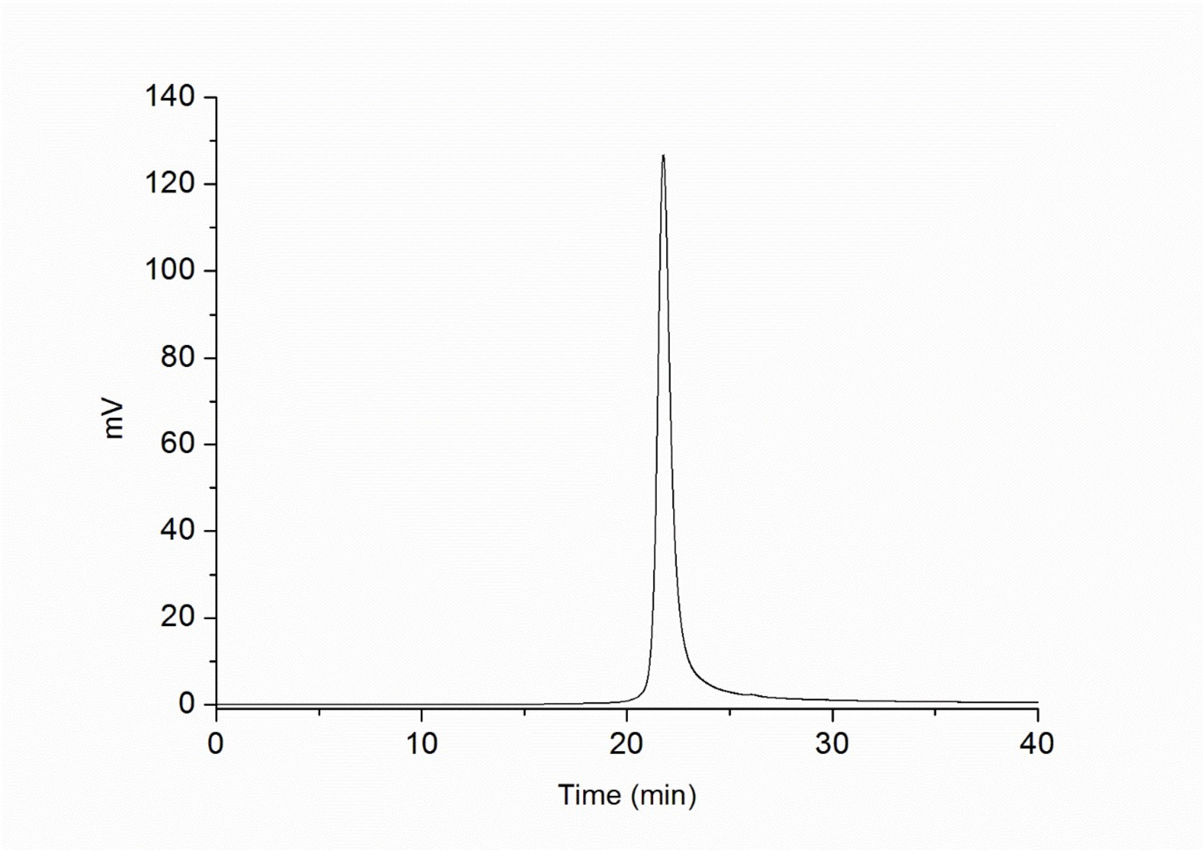


**eFigure 3**. **The profile of the TCan from size exclusion chromatography**.

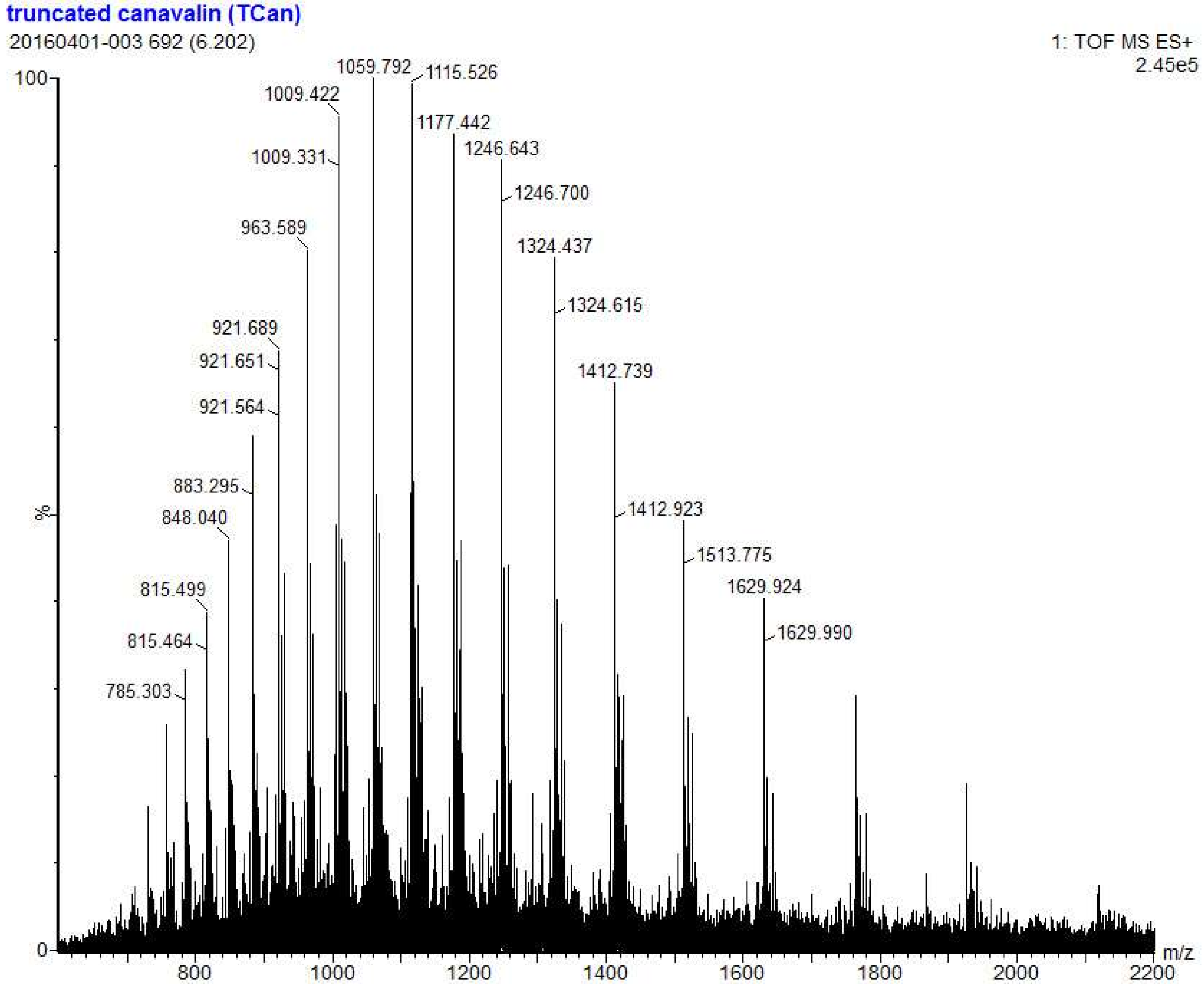


**eFigure 4**. **Multiple-charge electrospray mass spectra of TCan.** 10 μM of TCan with 0.1% formic acid in dielectric water by ESI-QTOF-mass spectrometry.

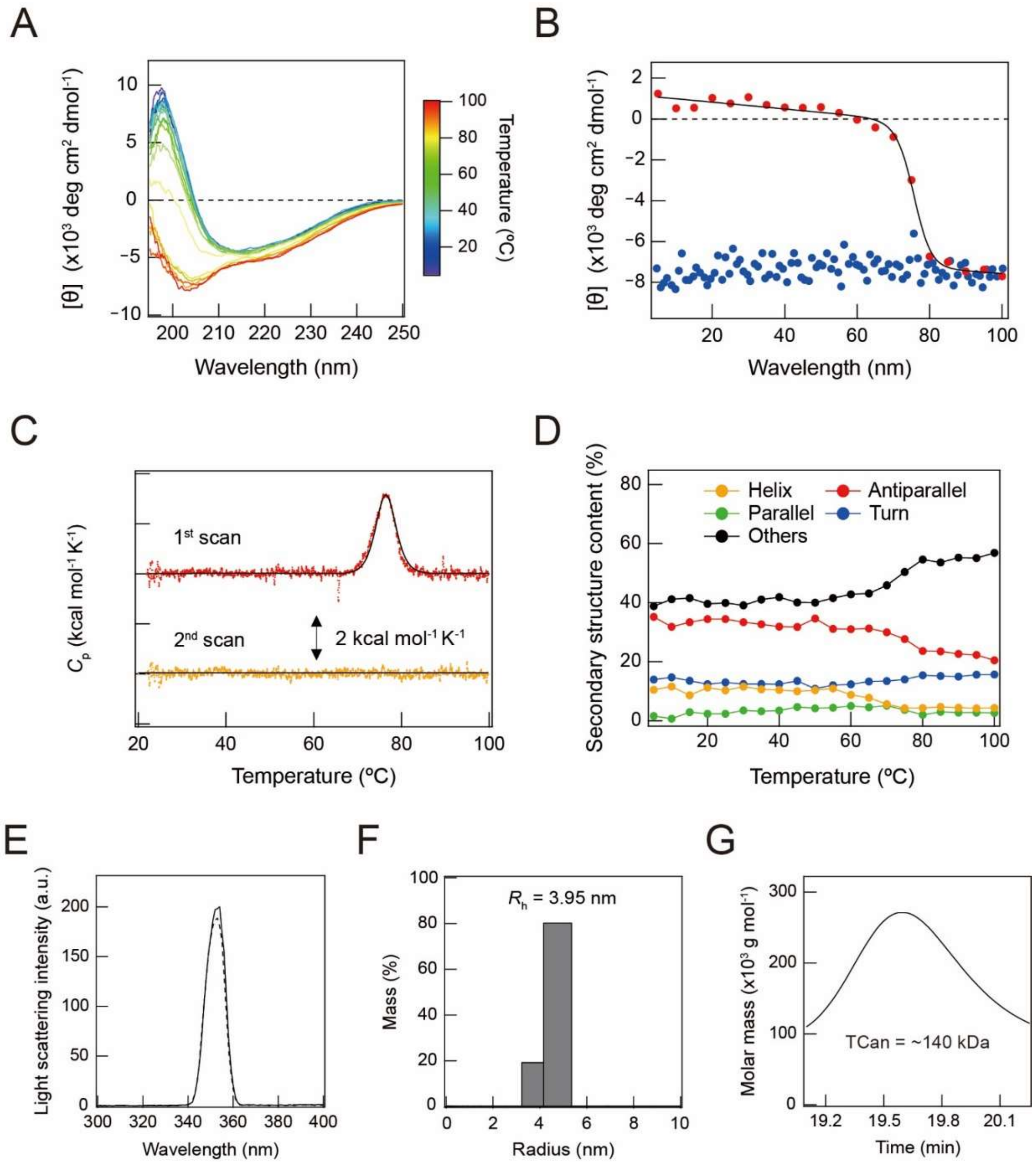


**eFigure 5. Molecular characterization of TCan using various biophysical approaches.** (A–D) Structure and stability of TCan examined by spectroscopy and calorimetry. (A) Far-UV circular dichroism (CD) spectra from 5 to 100 °C are shown with a color code on the right. (B) Heat scanning of CD intensities at 204 nm from 5 to 100 °C (red) and cooling down from 100 to 5 °C (blue). Fit curve is displayed with a continuous line (black). (C) The first (red) and second (yellow) differential scanning calorimetric thermograms of TCan. A continuous line indicates a fit curve. (D) Temperature-dependent changes in the content of the secondary structure of TCan: helical structure (yellow), antiparallel (red) and parallel (green) structure, turn structure (blue), and the other types of structures (black). (E–G) Characterization of oligomeric and colloidal states of TCan. (E) Light scattering spectra of TCan (continuous line) and 20 mM sodium phosphate buffer (dotted line) are shown. (F) Distribution of the hydrodynamic radius (Rh) of TCan obtained by dynamic light scattering before a heat scan. (G) Profile of multi-angle light scattering of TCan before a heat scan.

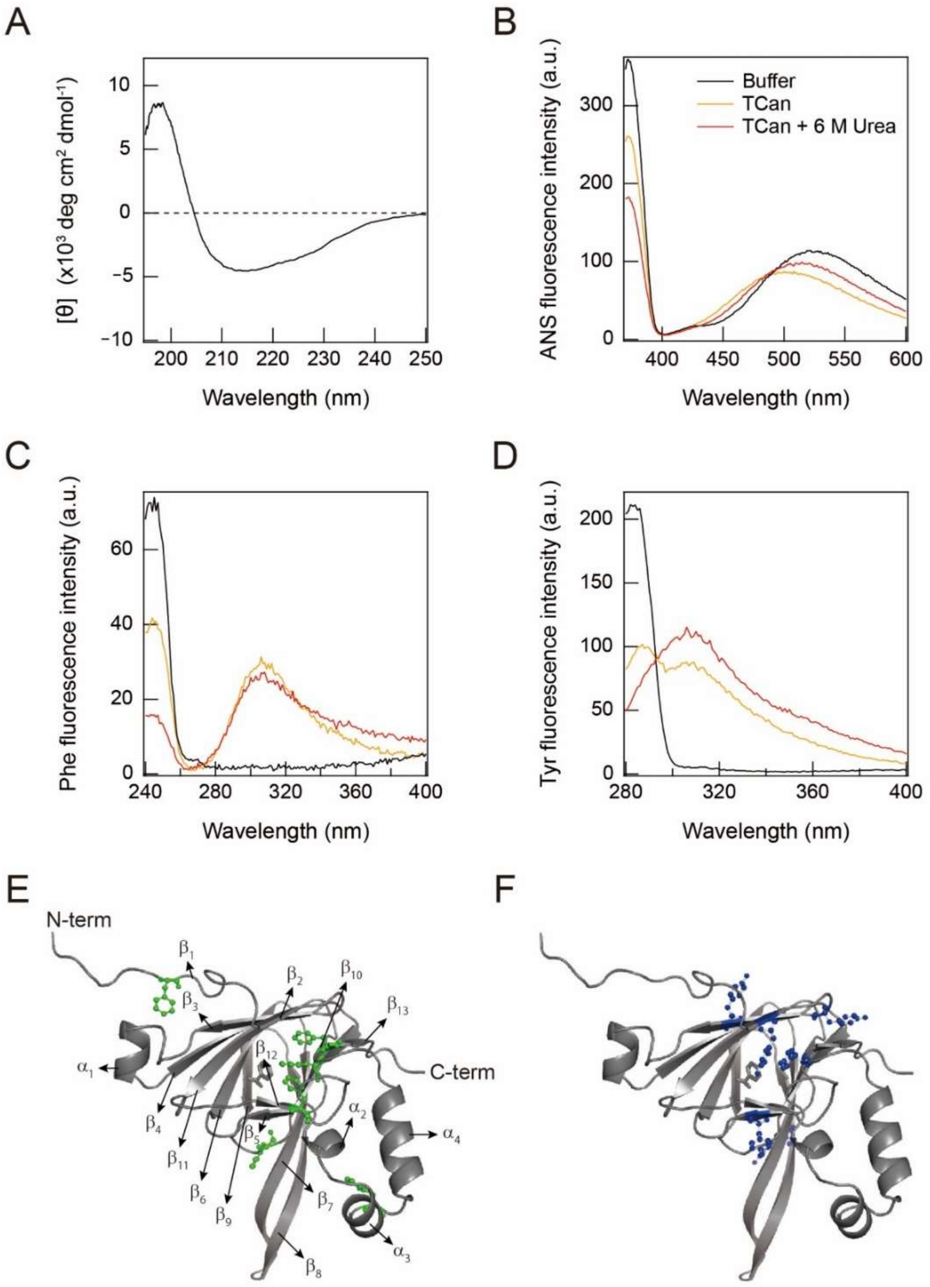


**eFigure 6. Structural characterization of TCan using spectroscopies.** (A) Far-UV circular dichroism spectrum at 25 °C. (B–D) Fluorescence emission spectra of ANS (B), Phe (C), and Tyr (D) of TCan in the absence (orange) and presence (red) of 6 M urea and 20 mM sodium phosphate buffer (black) are shown. (E and F) Three-dimensional structures of TCan generated from a crystal structure of canavalin (PDB ID: 6v7j)[11] are shown. Phe (green) (E) and Tyr (blue) (F) with a ball and stick model and secondary structures are indicated.

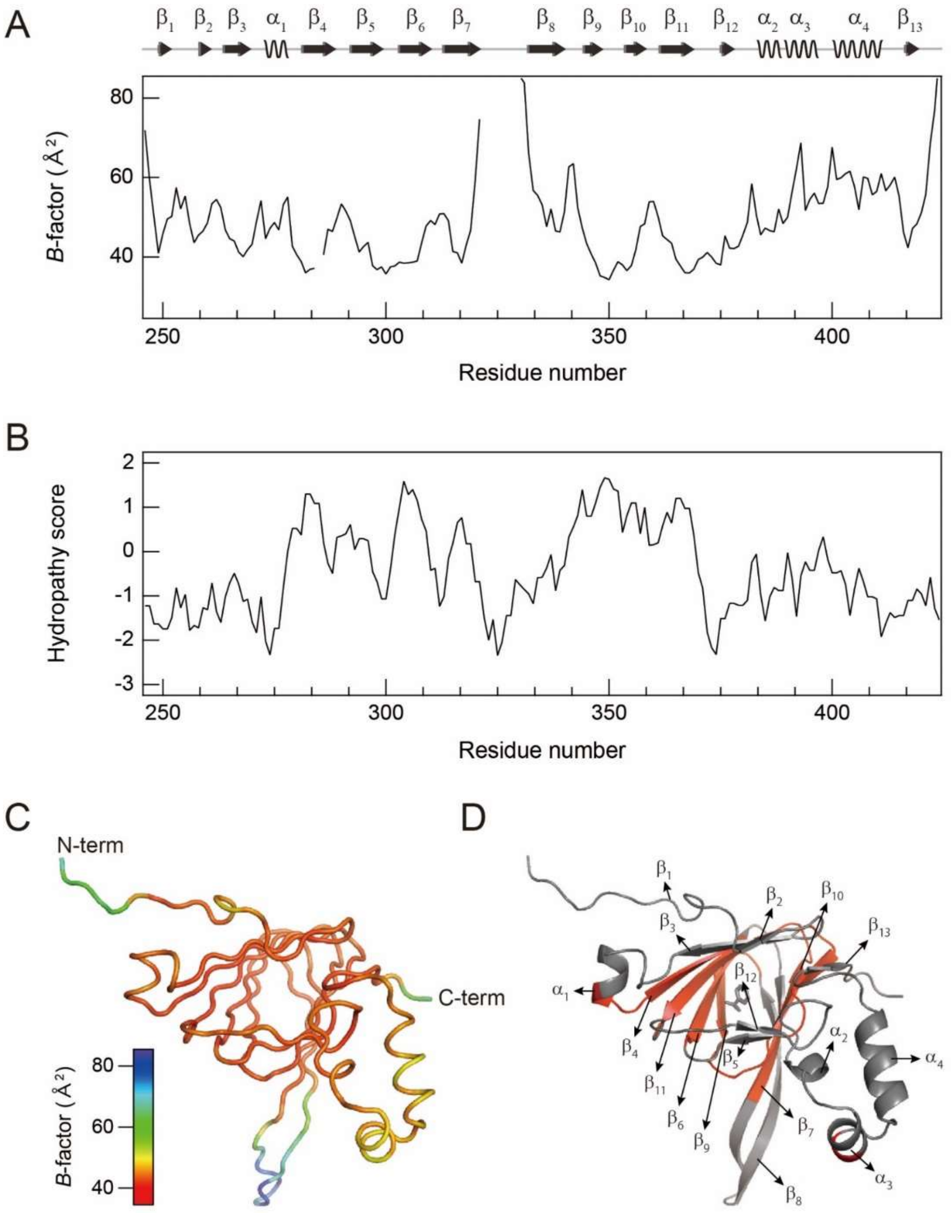


**eFigure 7. Intramolecular characterization of TCan.** (A and B) *B*-factor values (A) and hydropathy scores (B) are plotted against the residue number of TCan. *B*-factors were calculated by averaging *B*-factor values of the main-chain atoms (Cα, N, C', O) in the X-ray structure (PDB ID: 6v7j).[11] Hydropathy scores were estimated based on the scores reported by Kyte and Doolittle.[12] The secondary structure elements, α-helix (α) and β-structure (β), are schematically displayed at the top of A. (C) The structure of TCan is color-coded according to the values of *B*-factor as indicated in the scale bar. N- and C-termini are labeled as N-term and C-term, respectively. (D) Hydrophobic regions (in which hydropathy scores are positive) in TCan are colored red. Line and cartoon models of the crystal structure of TCan (PDB ID: 6v7j)[11] were used in C and D, respectively.

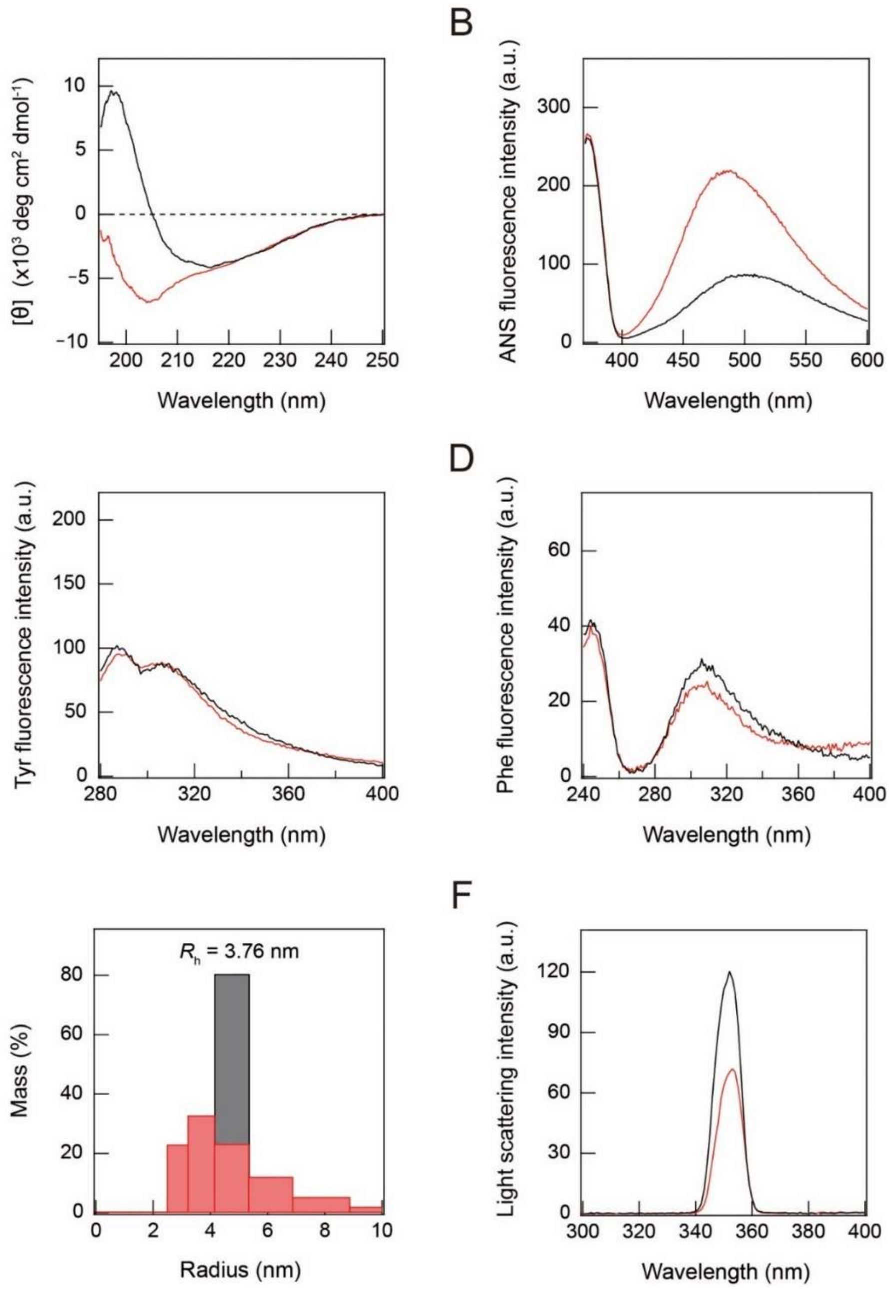


**eFigure 8. Irreversibility of the unfolding reaction of TCan after heat scanning.** (A) Far-UV CD spectra of TCan at 37 °C before (black) and after heat scanning (red). (B–D) Fluorescence emission spectra of ANS (B) as well as Phe (C) and Tyr (D) of TCan before (black) and after the heat treatment (red) are shown. (E and F) Distribution of hydrodynamic radius ($R_h$) obtained using dynamic light scattering (E) and light scattering spectra (F) before (black) and after the heat treatment (red) are shown.

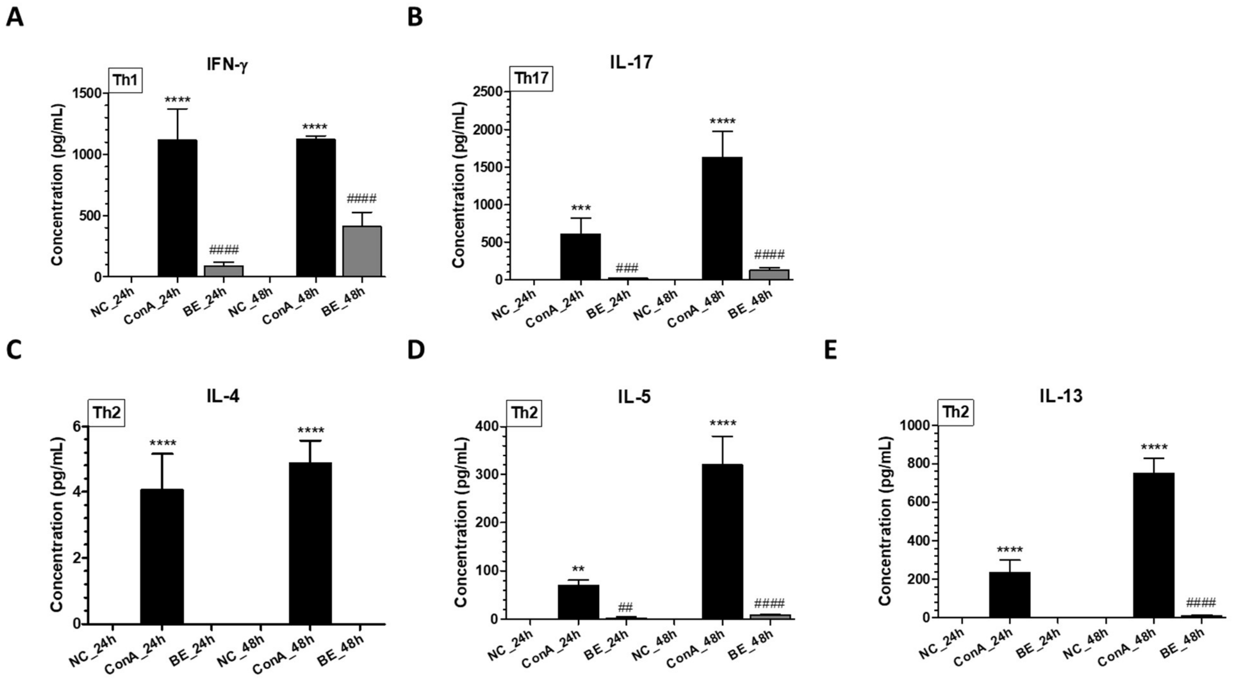


**eFigure 9. Changes in the expression levels of inflammatory cytokines**: (A) interferon gamma (IFN-γ), (B) interleukin (IL)-17, (C) IL-4, (D) IL-5, and (E) IL-13 in splenocytes of Con A- and BE-instilled C57BL/6 mice. Data represent mean ± SD (n=3 per group). **p < .01 and ****p < .0001 vs. naïve control (NC) group; ##p < .01 and ####p < .0001 vs. Con A group.

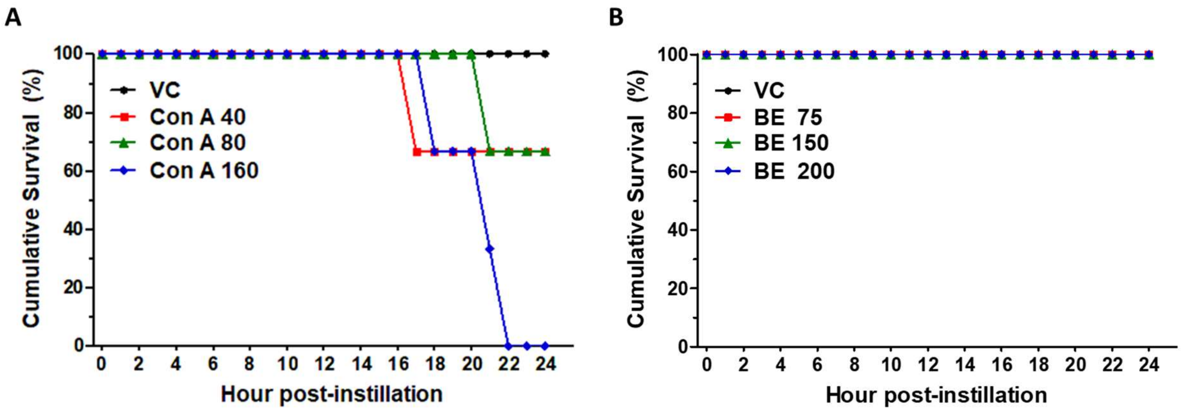


**eFigure 10**. **Effect of Con A and BE instillation on survival rate of C57BL/6 mice.** (A and B) Survival was recorded every hour up to 24 h after the instillation of Con A at a concentration of 40, 80 or 160 mg/kg body weight (BW), or BE at a concentration of 75, 160, and 200 mg/kg BW, and vehicle control (VC).

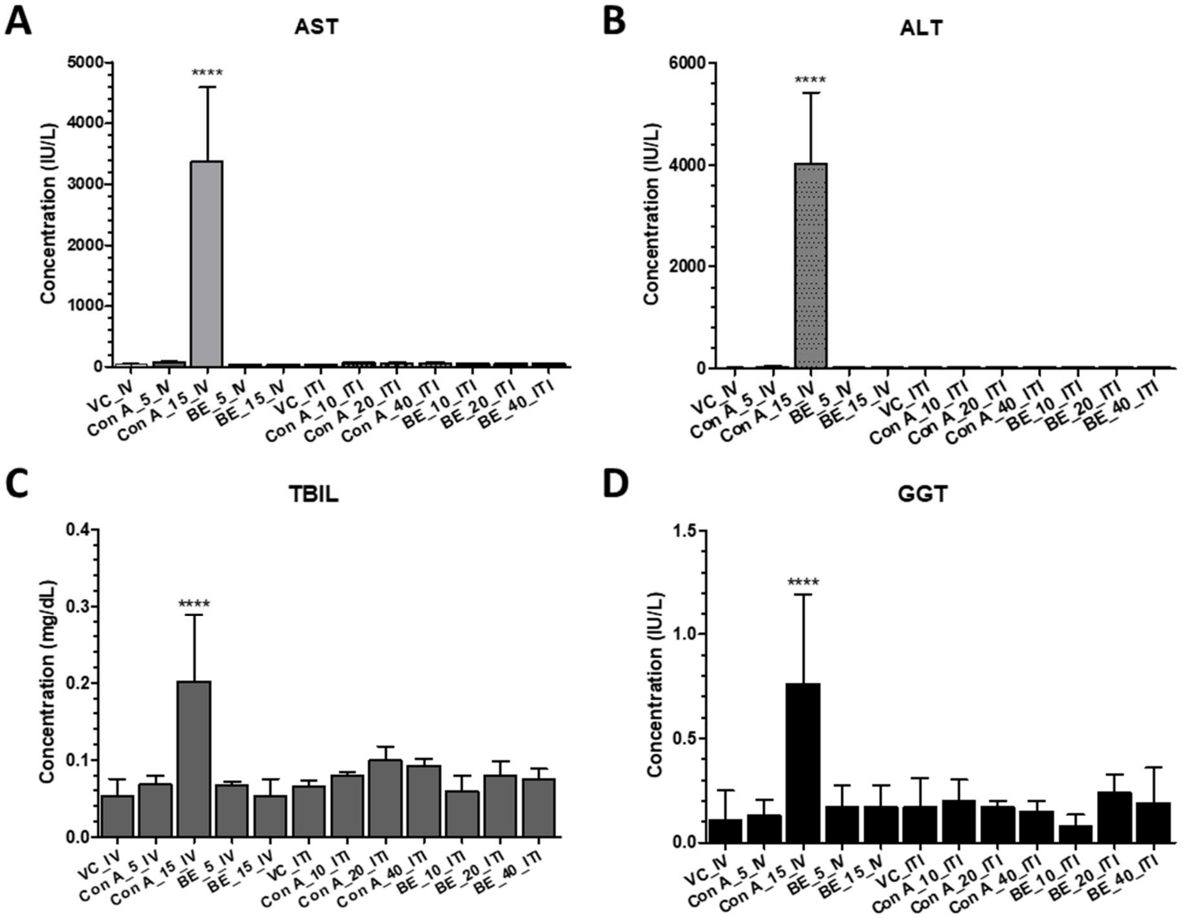


**eFigure 11. Changes in the levels of hepatic biomarkers (AST, ALT, TBIL, and GGT) in response to Con A and BE instillation in mice.** Biomarker concentrations were measured in the blood serum of Con A- and BE-administered mice (intravenously or intratracheal instillation; 5–15 or 10–40 mg/kg BW, respectively). Data represent mean ± SD (n=5 mice per group). $^{****}p < .0001$ vs. VC. AST, aspartate aminotransferase; ALT, alanine aminotransferase; TBIL, total bilirubin level; GGT, γ-glutamyl transferase.

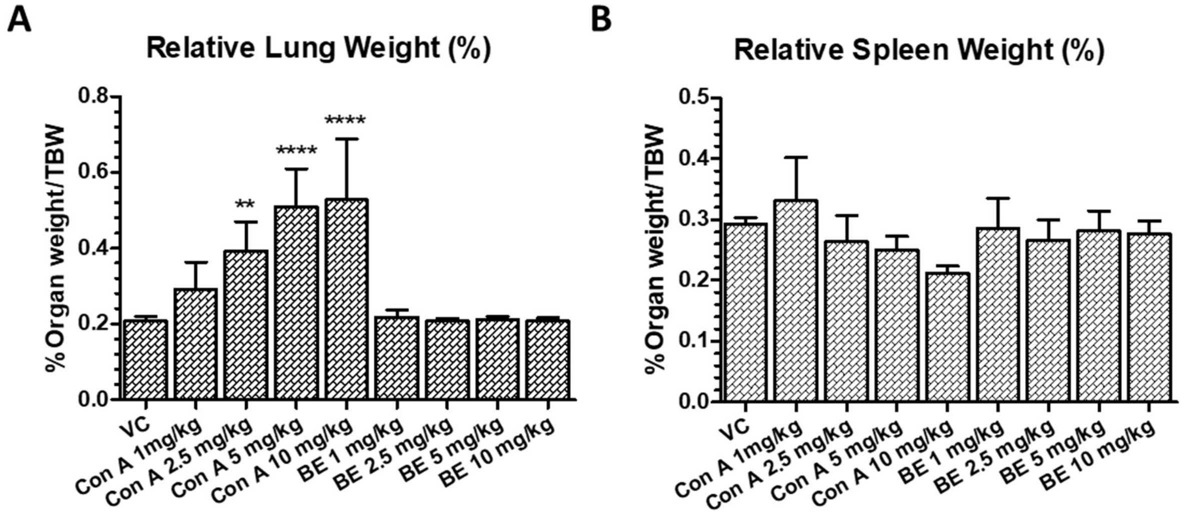


**eFigure 12. Changes in lung and spleen weight-to-body weight ratio after Con A and BE instillation in mice.** (A) Lung and (B) spleen weights at a dose of 1–10 mg/kg body weight (BW) were calculated with the following formula: relative organ weight = organ weight (g)/final body weight (g) × 100%. Data represent mean ± SD (n=5 per group). *p < .05, **p < .01, ***p < .001 and ****p < .0001 when compared to the corresponding vehicle control (VC), determined using Dunnett's t-test.

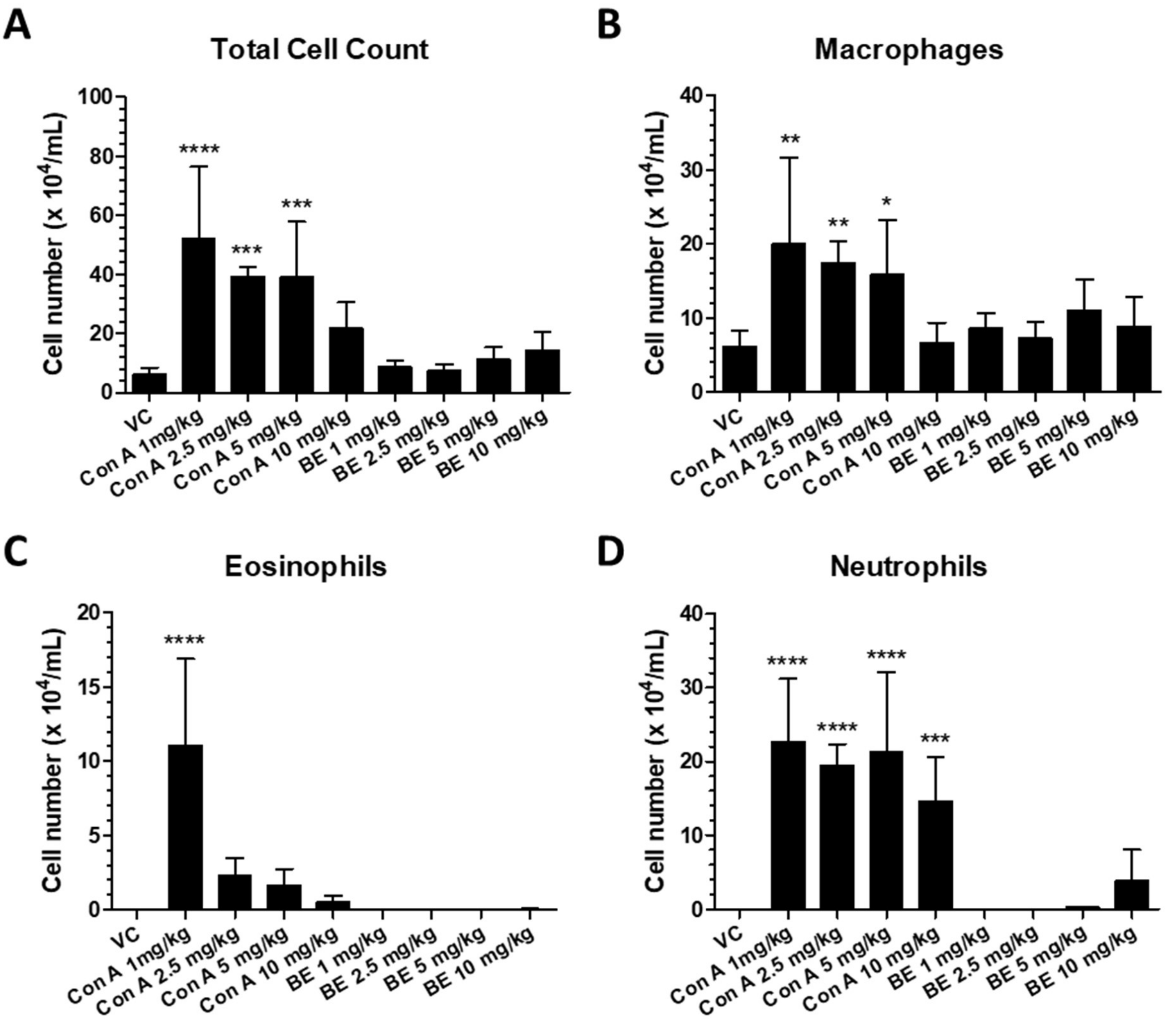


**eFigure 13**. **Changes in inflammatory cell numbers in bronchoalveolar lavage fluid (BALF) of mice following Con A and BE instillation.** (A) total cells, (B) macrophages, (C) eosinophils, and (D) neutrophils were counted in BALF. BALF was collected and analyzed 24 h after vehicle, Con A, or BE instillation. Data represent mean ± SD (n=5 per group). *p < .05, **p < .01, ***p < .001 and ****p < .0001 compared to the corresponding vehicle control (VC), determined using Dunnett's t-test.

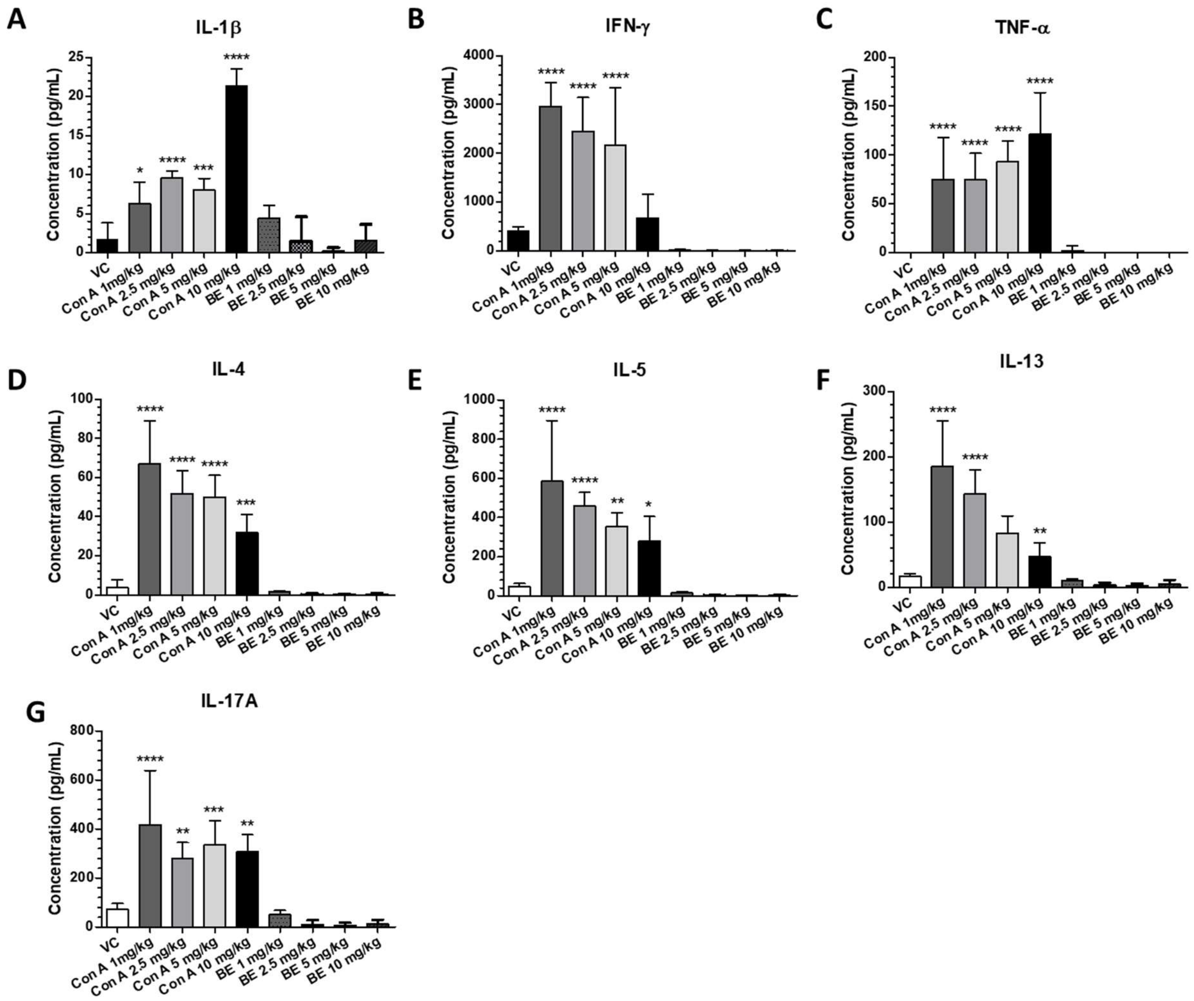


**eFigure 14**. **Changes in the expression of inflammatory cytokines.** (A) IL-1β, (B) IFN-γ, (C) TNF-α (D), IL-4 (E), IL-5 (F), IL-13, and (G) IL-17A in bronchoalveolar lavage fluid (BALF) after Con A or BE instillation at doses of 1–10 mg/kg BW in mice. Data represent mean ± SD (n=5 mice per group). *p < .05, **p < .01, ***p < .001 and ****p < .0001 when compared to the corresponding vehicle control (VC), determined using Dunnett's t-test. IL, interleukin, IFN-γ, interferon gamma, TNF-α, tumor necrosis factor alpha.

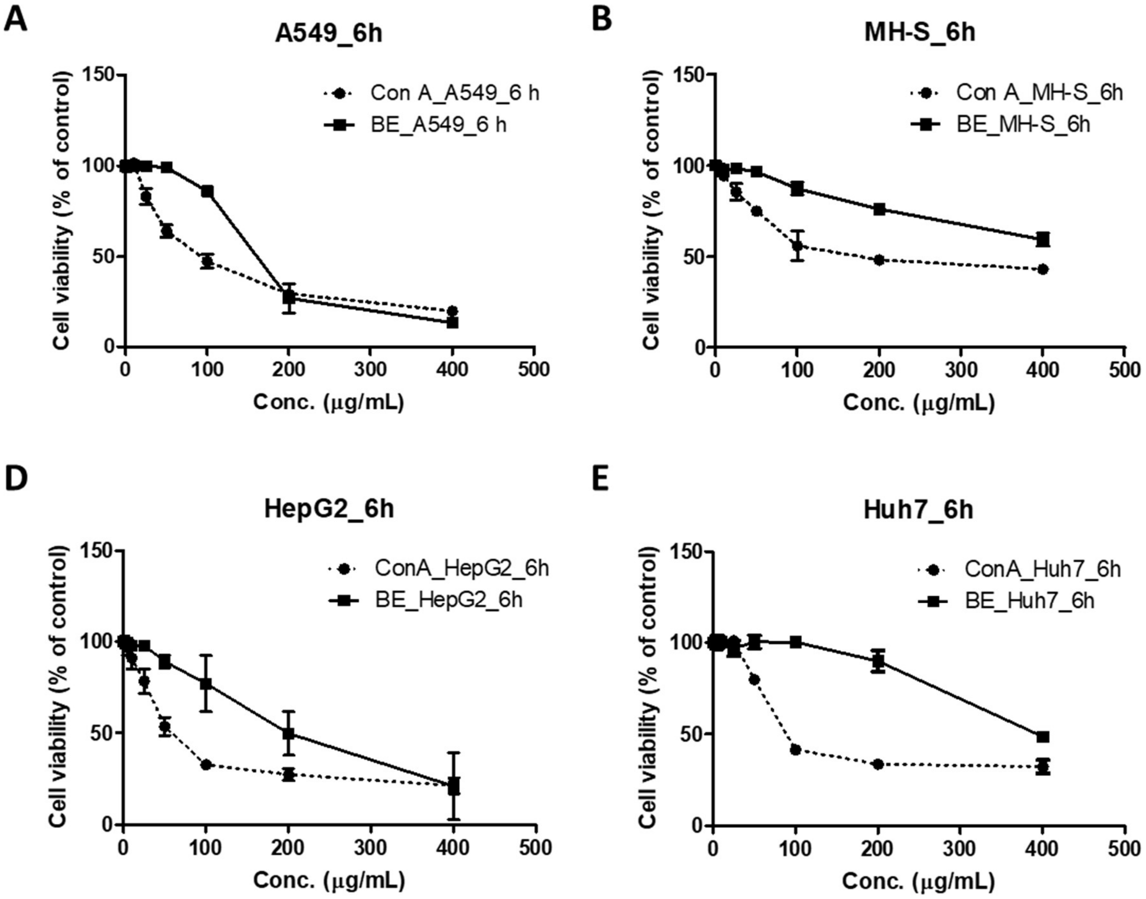


**eFigure 15**. ***In vitro* cytotoxicity of Con A and BE**. The toxicity of Con A and BE toward (A) A549, (B) MH-S, (C) HepG2, and (D) Huh7 cells at a concentration of 100–400 μg/mL. Cell viability was measured by counting cells using a NucleoCounter 6 h post infection and is expressed as a percentage of the viability of control cultures. Data represent mean ± SD (n=3 per group).

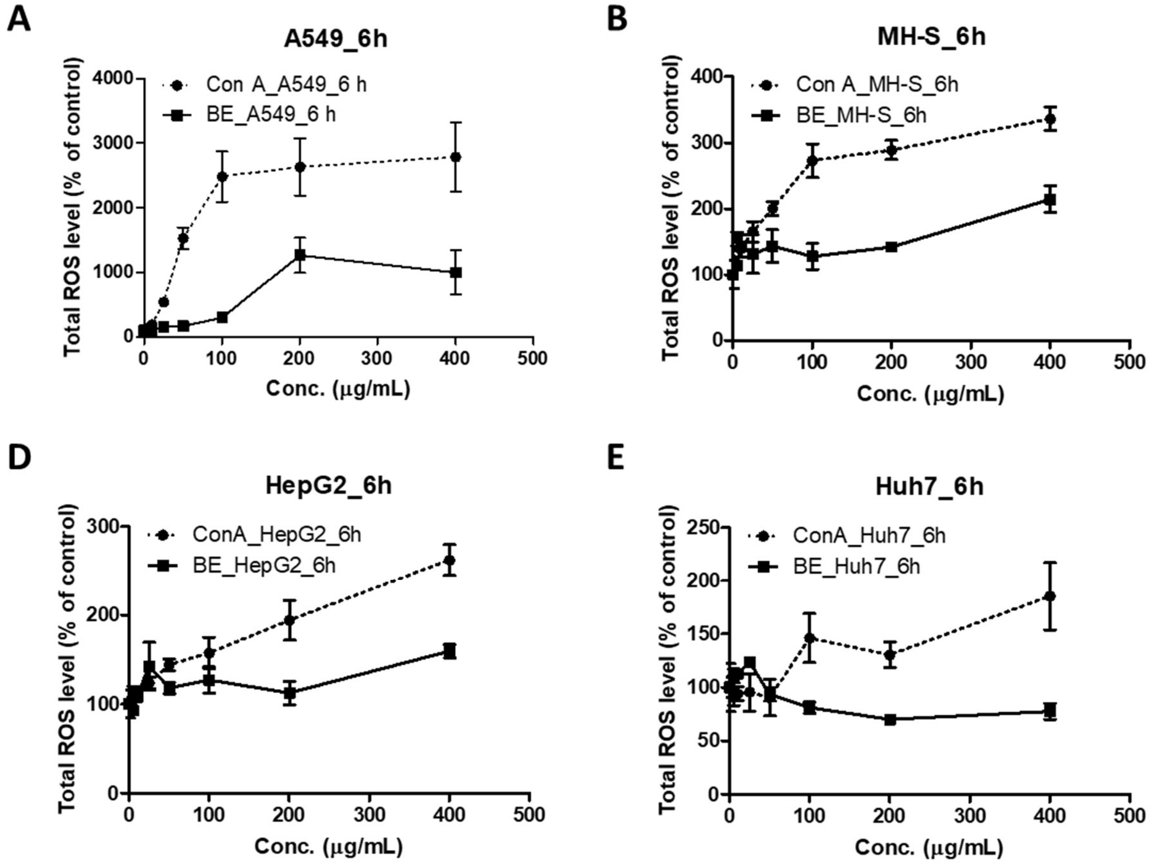


**eFigure 16**. **Reactive oxygen species (ROS) level in cells after Con A and BE treatment.** ROS generation in (A) A549, (B) MH-S, (C) HepG2, and (D) Huh7 cells after treatment with Con A and BE. ROS level was measured by DCFH-DA staining 6 h after treatment with Con A or BE. The ROS level in cells is expressed as a percentage of untreated controls. Data represent mean ± SD (n=3 per group).

#### Supplementary Tables

**eTable 1. Evaluation of the diagnostic performance of BG-RT-PCR and BG-Ag-RDTs for patients with COVID-19 throughout the infection duration (n=102)**

| Compared to NPS-RT-PCR | | COVID-19 patients | | | Asymptomatic cases | | |
|---|---|---|---|---|---|---|---|
| | | Positive | Negative | Total | Positive | Negative | Total |
| BG-RT-PCR | Positive | 83 | 0 | 83 | 23 | 0 | 23 |
| | Negative | 19 | 0 | 19 | 4 | 0 | 4 |
| | Total | 102 | 0 | 102 | 27 | 0 | 27 |
| Sensitivity (%) | | 81.4 (95% CI: 72.7–87.7) | | | 85.2 (95% CI: 67.5–94.1) | | |
| Specificity (%) | | *N.A.* | | | *N.A.* | | |
| PPA(%) | | 100 (95% CI: 95.6–100) | | | 100 (95% CI: 85.7–100) | | |
| NPA(%) | | 0 (95% CI: 0–16.8) | | | 0 (95% CI: 0–49.0) | | |
| **Compared to BG-RT-PCR** | | **Patients (Asymptomatic)[a]** | | | **Patients (Asymptomatic)[b]** | | |
| | | **Positive** | **Negative** | **Total** | **Positive** | **Negative** | **Total** |
| BG-Ag-RDTs | Positive | 54 (13) | 0 (0) | 54 (13) | 53 (13) | 0 (0) | 53 (13) |
| | Negative | 29 (10) | 19 (4) | 48 (14) | 30 (10) | 19 (4) | 49 (14) |
| | Total | 83 (23) | 19 (4) | 102 (27) | 83 (23) | 19 (4) | 102 (27) |
| Sensitivity (%) | | 65.1 (56.5) | | | 63.9 (56.6) | | |
| Specificity (%) | | 100 (100) | | | 100 (100) | | |
| PPA (%) | | 100 (100) | | | 100 (100) | | |
| NPA (%) | | 39.6 (28.6) | | | 38.8 (28.6) | | |
| **Compared to NPS-RT-PCR** | | | | | | | |
| BG-Ag-RDTs | Positive | 54 (13) | 0 (0) | 54 (13) | 53 (13) | 0 (0) | 53 (13) |
| | Negative | 48 (14) | 0 (0) | 48 (14) | 49 (14) | 0 (0) | 49 (14) |
| | Total | 102 (27) | 0 (0) | 102 (27) | 102 (27) | 0 (0) | 102 (27) |
| Sensitivity (%) | | 52.9 (48.1) | | | 52.0 (48.1) | | |
| Specificity (%) | | *N.A.* | | | *N.A.* | | |
| PPA (%) | | 100 (100) | | | 100 (100) | | |
| NPA (%) | | 0 (0) | | | 0 (0) | | |

*N.A.*: Not applicable; PPA, positive predictive agreement; NPA, negative predictive agreement; Two saliva-based Ag-RDTs were assessed: [a]Ag-RDTs is STANDARD™ Q COVID-19 Ag Saliva test, and [b]Ag-RDTs is Gmate® COVID-19 Ag Saliva.

**eTable 2. Results for the BG-RT-PCR and BG-Ag-RDTs from negative controls (n=100) and COVID-19 patients within 6 days of illness (n=45)**

| | | Participants (n=145) | | | Asymptomatic case (n=11) | | |
|---|---|---|---|---|---|---|---|
| | | Positive | Negative | Total | Positive | Negative | Total |
| **BG-RT-PCR compared to NPS-RT-PCR** | | | | | | | |
| | **Positive** | 45 | 0 | 45 | 11 | 0 | 11 |
| **BG-RT-PCR** | **Negative** | 0 | 100 | 100 | 0 | 0 | 0 |
| | **Total** | 45 | 100 | 145 | 11 | 0 | 11 |
| **BG-Ag-RDT compared to BG-RT-PCR** | | | | | | | |
| | **Positive** | 44 | 0 | 44 | 11 | 0 | 11 |
| **BG-Ag-RDT**[a] | **Negative** | 1 | 100 | 101 | 0 | 0 | 0 |
| | **Total** | 45 | 100 | 145 | 11 | 0 | 11 |
| **BG-Ag-RDT compared to NPS-RT-PCR** | | | | | | | |
| | **Positive** | 44 | 0 | 44 | 11 | 0 | 11 |
| **BG-Ag-RDT**[a] | **Negative** | 1 | 100 | 101 | 0 | 0 | 0 |
| | **Total** | 45 | 100 | 145 | 11 | 0 | 11 |

[a]Two saliva-based Ag-RDTs were assessed: 1) STANDARD™ Q COVID-19 Ag Saliva test, and 2) Gmate® COVID-19 Ag Saliva. There is no difference between the test results of two Ag-RDTs tested for negative controls and COVID-19 patients within 6 days of illness.

**eTable 3. Evaluation of the diagnostic performance of BG-RT-PCR and BG-Ag-RDTs for patients of SARS-CoV-2 variants within 6 days of illness**

| | | Alpha variant (n=6) | | | Delta variant (n=2) | | |
|---|---|---|---|---|---|---|---|
| | | **Positive** | **Negative** | **Total** | **Positive** | **Negative** | **Total** |
| **BG-RT-PCR compared to NPS-RT-PCR** | | | | | | | |
| **BG-RT-PCR** | **Positive** | 6 | 0 | 6 | 2 | 0 | 2 |
| | **Negative** | 0 | 0 | 0 | 0 | 0 | 0 |
| | **Total** | 6 | 0 | 6 | 2 | 0 | 2 |
| **BG-Ag-RDT compared to BG-RT-PCR** | | | | | | | |
| **BG-Ag-RDT[a]** | **Positive** | 6 | 0 | 6 | 2 | 0 | 2 |
| | **Negative** | 0 | 0 | 0 | 0 | 0 | 0 |
| | **Total** | 6 | 0 | 6 | 2 | 0 | 2 |
| **BG-Ag-RDT compared to NPS-RT-PCR** | | | | | | | |
| **BG-Ag-RDT[a]** | **Positive** | 6 | 0 | 6 | 2 | 0 | 2 |
| | **Negative** | 0 | 0 | 0 | 0 | 0 | 0 |
| | **Total** | 6 | 0 | 6 | 2 | 0 | 2 |

[a]Two saliva-based Ag-RDTs were assessed: 1) STANDARD™ Q COVID-19 Ag Saliva test, and 2) Gmate® COVID-19 Ag Saliva. There is no difference between the test results of two Ag-RDTs with the specimens tested positive for Alpha and Delta variants within 6 days of illness.

**Table S4. Sensitivity of BG-Ag-RDTs depending on Ct value of positive BG-RT-PCR samples**

| | | Within 6 days | Entire period |
|---|---|---|---|
| Ct ≤ 30 | Positive | 44 | 54 |
| | Negative | 1 | 18 |
| | Sensitivity (%) | 97.8<br>(95% CI, 88.4–99.6) | 75.0<br>(95% CI, 63.9–83.6) |
| Ct > 30 | Positive | 0 | 0 |
| | Negative | 0 | 11 |
| | Sensitivity (%) | *N.A.* | 0<br>(95% CI, 0–25.9) |

*N.A.*: Not applicable

**Table S5. Summary of thermodynamic parameters for the structural stability of TCan using circular dichroism spectroscopy**

| $^{CD}T_m$ (℃) | $^{CD}\Delta H$ (kcal/mol) | $\Delta C_p$ (kcal/mol K) |
|---|---|---|
| **75.4±0.4** | 100.2±20·0 | 2.2±0.3 |

**Table S6. Summary of thermodynamic parameters for the structural stability of TCan using differential scanning calorimetry**

| $^{DSC}T_m$ (℃) | $^{DSC}\Delta H_{cal}$ (kcal/mol) | $^{DSC}\Delta H_{VH}$ (kcal/mol) |
|---|---|---|
| **76.2±0.0 (76.21±0.01)** | 19.3±0.1 | 158.0±0.7 |
